# EXCESS DEATHS FROM ALL CAUSES AND BY COVID-19 IN BRAZIL IN 2020

**DOI:** 10.1101/2021.08.13.21261939

**Authors:** Alcione Miranda dos Santos, Bruno Feres de Souza, Carolina Abreu de Carvalho, Marcos Adriano Garcia Campos, Bruno Luciano Carneiro Alves de Oliveira, Eduardo Moraes Diniz, Maria dos Remédios Freitas Carvalho Branco, Rejane Christine de Sousa Queiroz, Vitória Abreu de Carvalho, Waleska Regina Machado Araújo, e Antônio Augusto Moura da Silva

## Abstract

**Objective:** To estimate the 2020 all-cause and COVID-19 excess mortality according to sex, age, race/color, and state, and to compare mortality rates by selected causes with that of the five previous years in Brazil.

**Methods:** Data from the Mortality Information System were used. Expected deaths for 2020 were estimated from 2015 to 2019 data using a negative binomial log-linear model.

**Results:** Excess deaths in Brazil in 2020 amounted to 13.7%, and the ratio of excess deaths to COVID-19 deaths was 0.90. Reductions in deaths from cardiovascular diseases (CVD), respiratory diseases, and external causes, and an increase in ill-defined causes were all noted. Excess deaths were also found to be heterogeneous, being higher in the Northern, Center-Western, and Northeastern states. In some states, the number of COVID-19 deaths was lower than that of excess deaths, whereas the opposite occurred in others. Moreover, excess deaths were higher in men, in those aged 20 to 59, and in black, yellow, or indigenous individuals. Meanwhile, excess mortality was lower in women, individuals aged 80 years or older, and in whites. Additionally, deaths among those aged 0 to 19 were 7.2% lower than expected, with reduction in mortality from respiratory diseases and external causes. There was also a drop in mortality due to external causes in men and in those aged 20 to 39 years. Furthermore, reductions in deaths from CVD and neoplasms were noted in some states and groups.

**Conclusion:** There is evidence of underreporting of COVID-19 deaths and of the possible impact of restrictive measures in the reduction of deaths from external causes and respiratory diseases. The impacts of COVID-19 on mortality were heterogeneous among the states and groups, revealing that regional, demographic, socioeconomic, and racial differences expose individuals in distinct ways to the risk of death from both COVID-19 and other causes.

## INTRODUCTION

Given the low RT-PCR (Reverse Transcription Polymerase Chain Reaction) for COVID-19 testing rate in Brazil, it is possible that some of the recorded COVID-19 deaths might have been due to other causes. Moreover, the restrictive measures adopted to control the pandemic may have caused variations in other causes of death. Excess deaths from all causes have been one of the most useful measures to assess the direct impact of COVID-19 on mortality. Additionally, it also assesses the indirect impact caused by reduced health services access in patients with other co-morbidities and the impact of restrictive measures adopted to fight the pandemic^1,2^.

Globally, Brazil is the second country with the highest number of COVID-19 deaths^3^. In the year 2020, 194,976 COVID-19 deaths were registered in the country, and by August 10, 2021, they totaled 564,773 deaths^4^. Despite the high number of recorded deaths, it is likely that there is still underreporting of deaths caused by COVID-19. Among the main causes of underreporting was the uncertainty of COVID-19 diagnosis in the first months of 2020, as it was a new disease and healthcare professionals had limited experience to certify these deaths^5,6^. Moreover, most COVID-19 deaths occurred among the elderly, who commonly present with multiple co-morbidities, consequently increasing the difficulty in identifying the basic cause of mortality^1^. Another factor that may have contributed to the underreporting of COVID-19 deaths in Brazil was the collapse of the healthcare network, as reported in some states^7^. Thus, it was likely that people who died outside health services would have a more flawed notification of the cause of death^8^.

Karlinsky and Kobak^9^ (2021) evaluated the excess mortality of 89 countries, observing that Peru had the highest annual excess mortality (146%). In this study, Brazil was in 7th place, with an excess mortality of 33% as of April 2021. It was also observed that the ratio between excess mortality and COVID-19 mortality differed among countries. Specifically, most of them presented with a ratio of above 1, suggesting underreporting of COVID-19 deaths, which was the case in Egypt (13.1), Russia (4.8), and Mexico (2.2). However, some countries, such as Belgium (0.6) and France (0.7), had ratios lower than 1, indicating that there were more COVID-19 deaths than excess deaths^9^. This probably occurred since the other causes of death, including deaths from influenza and external causes, decreased as a result of the distancing measures adopted to control the pandemic^9,10^.

In Brazil there are two mortality registration systems, both of which use the death declaration (DO in the Portuguese acronym) as the basic document. The Civil Registry (CR) is a registry-based system, whose information is compiled and disseminated by the IBGE (translated as the Brazilian Institute of Geography and Statistics), whereas the Mortality Information System (SIM in the Portuguese acronym) is based on the Ministry of Health. Underreporting of deaths has been reported to be low in both systems, with the SIM showing a lower rate than the CR. Notably, IBGE estimates showed that underreporting was 1.48% for the SIM and 4.00% for the CR in 2018, wherein underreporting in the SIM ranged from 0.29% in DF to 5.65% in AP^11^.

In Brazil, Silva et al.^12^ (2020) analyzed the excess mortality from March to May 2020, observing 39,146 more deaths than expected for this period. Moreover, the highest recorded excess mortality was found in the capitals of the North, Northeast, and Southeast regions. Werneck et al.^13^ (2021), on the other hand, analyzed the excess mortality in Brazil from April 2020 to March 2021, reporting 305,000 more deaths than expected. Additionally, the results of this study showed that excess deaths varied among Brazilian states, with the highest percentage in Amazonas (84%) and the lowest in Piauí (11%). Although both studies used data from the National Civil Registry Information Center, they only included natural causes, limiting the observation of the full impact of COVID-19 on mortality. This was because both studies did not capture the reductions that may have occurred in deaths from external causes, which was a result of reduced mobility during the pandemic^9,14^.

Given these findings, the objectives of this study were as follows: (1) to estimate the 2020 excess mortality from all causes and by COVID-19 in Brazil, according to sex, age, race/color, and state, based on the SIM data; and (2) to compare the mortality rates by selected causes in 2020 to the median rates of the years 2015 to 2019 according to state, sex, age, and race/color. To the best of our knowledge, there has been no published work in Brazil that used the SIM to calculate excess deaths, which included external causes and stratified excess mortality by sex, age, and race/color, or that has explored the variation of cause-specific mortality rates.

## METHODS

In this study, all individual records regarding non-fetal deaths based on place of residence from the years 2015 to 2020 available in the SIM were included, which was obtained from the website *opendatasus*.*saude*.*gov*.*br* and was accessed last August 4, 2021. Deaths considered for the year 2020 corresponded to all deaths that occurred in the period from December 29, 2019 to January 2, 2021 (epidemiological weeks 1 to 53). The SIM records for 2020 were still considered preliminary, and the last update was made on August 3, 2021.

Information on the date of death, basic cause of death, age (in complete years), sex, race/color, and the municipality code of the residence of the deceased were obtained. Deaths with no information (unknown) regarding sex (n=3,708), age (n=16,854), and race/color (n=261,955) were excluded in the specific analyses for these variables. The education variable was not included, because more than 20% of the values were missing.

Considering the International Classification of Diseases (ICD: 10) the causes were classified as the following: COVID-19 (code B34.2), other infectious diseases, neoplasms, cardiovascular diseases, respiratory diseases, ill-defined causes, external causes, and others. Deaths were then analyzed by epidemiological week and stratified according to sex (male and female), age groups (0-19, 20-39, 40-59, 60-79, and ≥ 80 years), race/color (white, black, brown and others), and state. Individuals of yellow and indigenous race/color were aggregated in others, due to the small number of deaths in each category.

Data regarding the population of Brazil were obtained from the IBGE, with estimates abstracted for each state, sex and age group in each year^15^. To estimate the Brazilian population by race/color in 2020, we used the median proportions of each category in the quarterly continuous national surveys taken via household sampling conducted over the past six years^16^ and multiplied them by the Brazilian population of each year.

### Statistical analysis

Initially, the identification of aberrations, which are significant changes in the series, including natural disasters and epidemics, was performed. For this, the procedure proposed by Farrington et al.^17^ (1996) was used, wherein the series of deaths from 2010 to 2014 was used as reference, and the series from 2015 to 2020 was used for monitoring. Although excess mortality from Chikungunya and influenza were detected in some states in 2016, these aberrations had little influence on the estimates of expected deaths for 2020 and were therefore disregarded. More details regarding this procedure are described in the supplementary material.

To obtain the number of expected deaths for 2020, the population was stratified by age group, sex, race/skin color, and state. For each group, a generalized linear model was adjusted to estimate the number of expected deaths in 2020 from the series of deaths recorded from 2015 to 2019. The number of deaths in epidemiological week *t* (*t* = 1,…,52), denoted by *yt*, assuming that *yt* followed a negative binomial distribution, with mean μ0._*t*_ and fixed dispersion parameter *α*, was estimated using the model described in Serfling^18^ (1963), in which *μ*0,_*t*_ was modeled by:

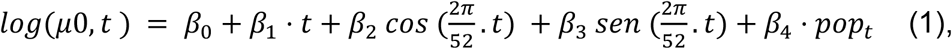

wherein *log* represents the natural logarithm, *β*_0_ the intercept, *β*_*i*_ (*i = 1*,…,4) the regressor parameters estimated by the model, and *pop*_*t*_ the population size in year *t*. For model fitting, the R surveillance package was used^19^. Furthermore, the number of deaths expected for week 52 was assumed for week 53.

Absolute excess deaths were obtained by subtracting the number of observed deaths from the number of expected deaths in each epidemiological week for each state, sex, age group, and race/color. Both weeks with excess and deficit of deaths were considered. The percentage of excess deaths was obtained by dividing the number of excess deaths by the number of expected deaths multiplied by 100. The ratio between excess deaths and the number of COVID-19 deaths was also calculated.

Afterwards, the 2020 mortality rates by selected causes were compared with the median of these rates for the years 2015 to 2019.

All analyses in this study were performed in the R software (version 4.1.0) and RStudio (version 1.4.1717). The scripts can be accessed at the following website: *https://github.com/aamouradasilva/excess_mortality.git*.

### Ethical aspects

Since the present study used public and unidentified secondary data, approval from the Ethics Committee on Research in Human Beings was waived (Resolution no. 510/2016 of the National Health Council).

## RESULTS

In 2020, 1,551,673 deaths were registered in the SIM, which was 13.7% more than what was expected. Among these deaths, the number of COVID-19 deaths (208,518) was higher than excess deaths (187,070), resulting in a ratio of excess deaths to COVID-19 deaths of 0.90 (Table 1). Notably, excess mortality began in March and April, peaked in May, decreased until September, and increased again from October onwards. Until the beginning of May, excess deaths from COVID-19 (representing the distance between the sum of expected and COVID-19 deaths and the number of expected deaths) coincided with excess deaths from all causes. However, starting from the end of May, deaths from COVID-19 exceeded excess deaths from all causes (Figure 1). On comparison of mortality rates by selected causes in 2020 with the median of these rates from 2015 to 2019, reductions in deaths caused by cardiovascular diseases, respiratory diseases, and external causes, as well as an increases in mortality from ill-defined causes, were all noted (Figure 2).

**Table 1.** Excess deaths due to all causes and deaths from COVID-19 by state in Brazil, 2020

| State | Expected deaths | Observed deaths | Excess deaths (n) | Excess deaths (%) | Deaths from COVID-19 (n) | Ratio of excess deaths to COVID-19 deaths |
| --- | --- | --- | --- | --- | --- | --- |
| RO | 8,380 | 10,295 | 1,915 | 22.9 | 1,907 | 1.00 |
| AC | 4,313 | 4,895 | 582 | 13.5 | 813 | 0.72 |
| AM | 18,574 | 24,997 | 6,423 | 34.6 | 5,478 | 1.17 |
| RR | 3,106 | 3,515 | 409 | 13.2 | 803 | 0.51 |
| PA | 42,048 | 51,982 | 9,934 | 23.6 | 7,796 | 1.27 |
| AP | 3,679 | 4,553 | 874 | 23.8 | 1,106 | 0.79 |
| TO | 8,263 | 9,221 | 958 | 11.6 | 1,245 | 0.77 |
| MA | 35,556 | 43,303 | 7,747 | 21.8 | 5,124 | 1.51 |
| PI | 21,514 | 23,353 | 1,839 | 8.5 | 2,796 | 0.66 |
| CE | 58,570 | 69,201 | 10,631 | 18.2 | 11,828 | 0.90 |
| RN | 22,078 | 24,480 | 2,402 | 10.9 | 3,077 | 0.78 |
| PB | 27,379 | 30,852 | 3,473 | 12.7 | 3,424 | 1.01 |
| PE | 63,602 | 77,159 | 13,557 | 21.3 | 10,624 | 1.28 |
| AL | 20,050 | 24,159 | 4,109 | 20.5 | 3,492 | 1.18 |
| SE | 13,251 | 15,919 | 2,668 | 20.1 | 2,169 | 1.23 |
| BA | 93,854 | 104,420 | 10,566 | 11.3 | 10,524 | 1.00 |
| MG | 141,695 | 150,962 | 9,267 | 6.5 | 13,418 | 0.69 |
| ES | 24,852 | 28,953 | 4,101 | 16.5 | 4,703 | 0.87 |
| RJ | 146,253 | 170,522 | 24,269 | 16.6 | 30,324 | 0.80 |
| SP | 308,807 | 351,814 | 43,007 | 13.9 | 48,178 | 0.89 |
| PR | 75,237 | 81,771 | 6,534 | 8.7 | 8,466 | 0.77 |
| SC | 43,261 | 45,246 | 1,985 | 4.6 | 4,857 | 0.41 |
| RS | 90,237 | 93,181 | 2,944 | 3.3 | 8,791 | 0.33 |
| MS | 17,135 | 19,169 | 2,034 | 11.9 | 2,219 | 0.92 |
| MT | 18,779 | 23,356 | 4,577 | 24.4 | 4,292 | 1.07 |
| GO | 41,265 | 48,045 | 6,780 | 16.4 | 7,766 | 0.87 |
| DF | 12,816 | 16,350 | 3,534 | 27.6 | 3,298 | 1.07 |
| Brazil | 1,364,603 | 1,551,673 | 187,070 | 13.7 | 208,518 | 0.90 |

**Figure 1.**
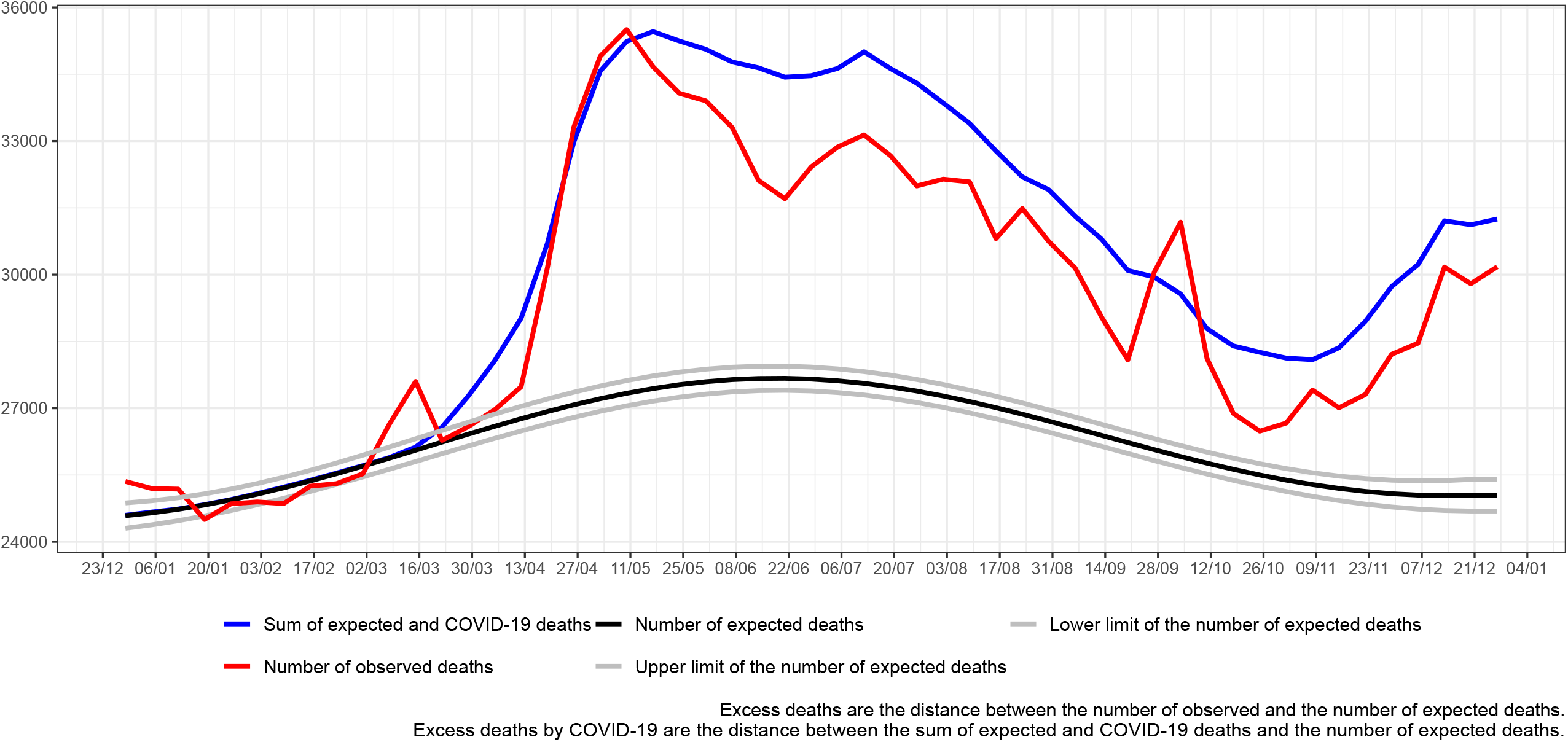
Excess deaths due to all causes and to COVID-19 by epidemiological week, Brazil, 2020

**Figure 2.**
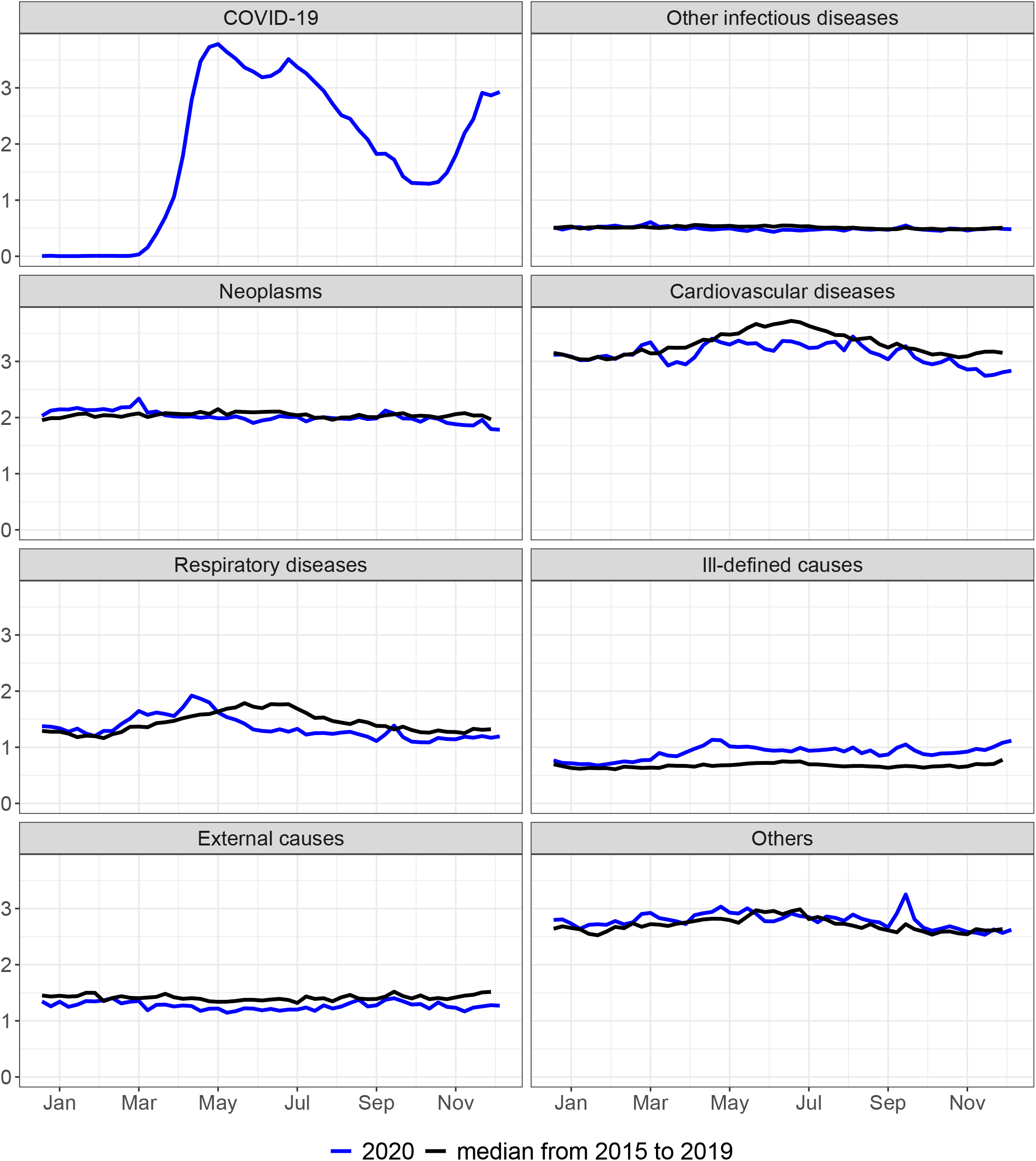
Mortality rate (per 100.000) by epidemiological week according to selected causes, Brazil, 2015 to 2020

The Brazilian states presented variations in the proportional excess deaths, with the biggest percentages occurring in Northern states, especially AM (34.6%), AP (23.8%), and PA (23.6%), and Center-Western states, particularly DF (27.6%) and MT (24.4%). On the other hand, the percentage of excess deaths was smaller in Southern states, with prominence in RS (3.3%), MG (6.5%), and also in PI (8.5%). In most states, the ratio between excess deaths and COVID-19 deaths was lower than 1, especially in RS (0.33), SC (0.41), RR (0.51), and PI (0.66). However, in other states, this ratio was higher than 1, as seen in MA (1.51), PE (1.28), and PA (1.27) (Table 1). Excess all-cause deaths and excess COVID-19 deaths by epidemiological week and state are presented by geographical region in Supplementary Figures (SF) 1 to 5. In AM, PA (SF 1), MA, and PE (SF 2), COVID-19 deaths were below excess deaths during the epidemic peak. In other states, such as MG, SP (SF 3), SC, and RS (SF 4), the opposite was true.

The comparison of mortality rates in 2020 by selected causes with the median of these rates from 2015 to 2019 by epidemiological week and state are found in SF 8 to 34. In AM (SF 10), PA (SF 12), and MA (SF 15), the peak in COVID-19 deaths coincided with the peaks in deaths from cardiovascular, respiratory, ill-defined, and other causes. In other states, such as MG (SF 24), RJ (SF 26), and RS (SF 30), reductions in deaths from cardiovascular diseases, respiratory diseases, and external causes were evident in 2020 in comparison to the five previous years. Moreover, in other states, it was possible to observe reductions in deaths from cardiovascular and respiratory diseases, such as SP (SF 27), and PR (SF 28). Furthermore, certain states, such as PE (SF 20), BA (SF 23), RJ (SF 26), SP (SF 27), RS (SF 30), MS (SF 31), MT (SF 34), and DF (SF 35) showed an increase in mortality from ill-defined causes.

The percentage of excess deaths was higher in males (15.7%) than in females (11.3%). For both sexes, the ratio between excess deaths from all causes and COVID-19 deaths was lower than 1, which was more markedly observed in females (SF 6). Additionally, reductions in mortality rates from cardiovascular and respiratory diseases in 2020 as compared to the five previous years, as well as an increase in mortality rate due to ill-defined causes, were observed in both sexes. In males, there was also a reduction in deaths from external causes (SF 35 and 36).

The 40-59 age group had the highest proportional excess deaths (19.3%), followed by the groups aged 20-39 years (17.6%) and 60-79 years (17.3%). Meanwhile, the lowest percentage of excess deaths was observed in the age group ≥ 80 years (10.1%). In the 0-19 age group, the number of observed deaths was lower than expected, resulting to a mortality deficit of −7.2% (Table 2), which occurred from March onwards and coincided with the time of the first COVID-19 deaths. In the 20-39 age group, the number of COVID-19 deaths was below excess deaths during the entire period, whereas in the 40-59 age group, COVID-19 deaths coincided with excess deaths. In the last two age groups, 60-79 and ≥ 80 years, the number of COVID-19 deaths exceeded excess deaths (Figure 3). Additionally, mortality rates due to respiratory, external causes, and other causes in 2020 were well below the rates from 2015 to 2019 among those under 20 years of age (SF 37). In the 20-39 age group, there was a sharp drop in mortality from external causes and a slight increase in mortality from ill-defined causes (SF 38). In the 40-59 age group, there was a fall in deaths from neoplasms, cardiovascular diseases, and external causes, and an increase of mortality from ill-defined causes (SF 39). Lastly, in the age groups ≥ 60 years old, reductions in deaths from neoplasms, cardiovascular diseases, and respiratory diseases, as well as an increase in mortality from ill-defined causes, were all noted (SF 40 and 41).

**Table 2.**
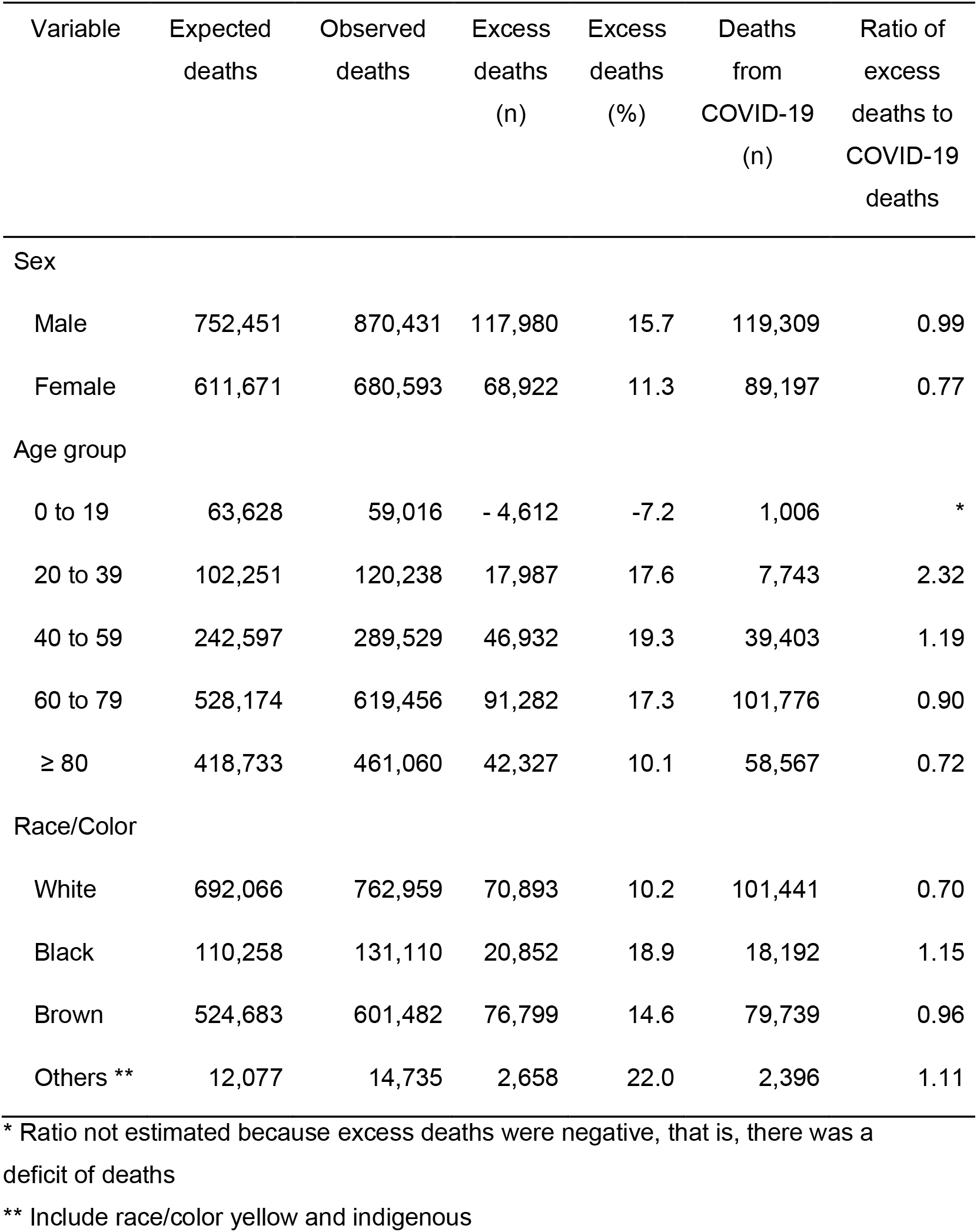
Excess deaths due to all causes and deaths from COVID-19 by sex, age group and race/skin color in Brazil, 2020

| Variable | Expected deaths | Observed deaths | Excess deaths (n) | Excess deaths (%) | Deaths from COVID-19 (n) | Ratio of excess deaths to COVID-19 deaths |
| --- | --- | --- | --- | --- | --- | --- |
| Sex |  |  |  |  |  |  |
| Male | 752,451 | 870,431 | 117,980 | 15.7 | 119,309 | 0.99 |
| Female | 611,671 | 680,593 | 68,922 | 11.3 | 89,197 | 0.77 |
| Age group |  |  |  |  |  |  |
| 0 to 19 | 63,628 | 59,016 | - 4,612 | -7.2 | 1,006 | * |
| 20 to 39 | 102,251 | 120,238 | 17,987 | 17.6 | 7,743 | 2.32 |
| 40 to 59 | 242,597 | 289,529 | 46,932 | 19.3 | 39,403 | 1.19 |
| 60 to 79 | 528,174 | 619,456 | 91,282 | 17.3 | 101,776 | 0.90 |
| ≥ 80 | 418,733 | 461,060 | 42,327 | 10.1 | 58,567 | 0.72 |
| Race/Color |  |  |  |  |  |  |
| White | 692,066 | 762,959 | 70,893 | 10.2 | 101,441 | 0.70 |
| Black | 110,258 | 131,110 | 20,852 | 18.9 | 18,192 | 1.15 |
| Brown | 524,683 | 601,482 | 76,799 | 14.6 | 79,739 | 0.96 |
| Others ** | 12,077 | 14,735 | 2,658 | 22.0 | 2,396 | 1.11 |

\* Ratio not estimated because excess deaths were negative, that is, there was a deficit of deaths
\*\* Include race/color yellow and indigenous

**Figure 3.**
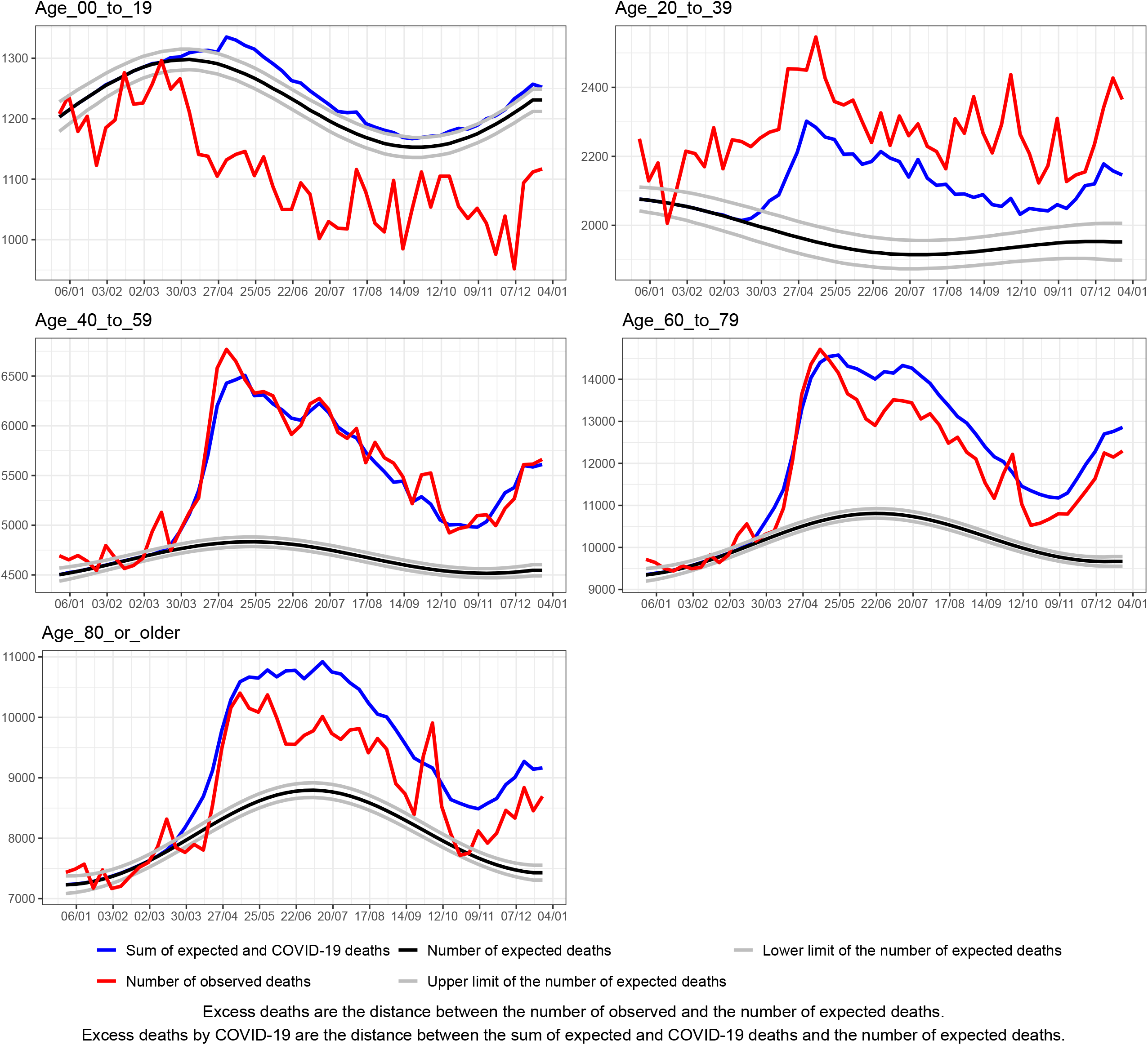
Excess deaths by all causes and by COVID=19 by epidemiological week according to age, Brazil, 2020

Regarding race/color, the highest proportional excess mortality occurred in other races/colors (yellow and indigenous, 22.0%) and in blacks (18.9%). Brown individuals had an intermediate percentage (14.6%) of excess deaths, whereas whites had the lowest proportional excess deaths (10.2%) (Table 2). Among brown subjects and more intensely among whites, the number of COVID-19 deaths surpassed excess deaths, while in blacks and other races/colors, these numbers were almost coincident (SF 7). Notably, in whites, there was a drop in deaths from neoplasms, cardiovascular diseases, respiratory diseases, and external causes, as well as an increase in deaths from ill-defined causes (SF 42). Furthermore, in the black and brown race groups, an increase in deaths from ill-defined causes and other causes were observed, additionally showing a slight decrease in mortality due to external causes (SF 43 and 44).

## DISCUSSION

The estimates presented in this study indicated a 13.6% excess mortality from all causes in Brazil in 2020. The number of COVID-19 deaths was higher than excess deaths, resulting in a ratio of excess deaths to COVID-19 deaths of 0.90, which suggested that there was a reduction in some causes of death. This was asserted by the findings of decreases in mortality rates from cardiovascular diseases, respiratory diseases, and external causes, as compared to the previous five years.

Data from our study suggest the occurrence of underreporting of COVID-19 deaths, as described in other articles^6,7,20,21^. In several Brazilian states, there was an increase in mortality rate from ill-defined causes. Particularly, in MA, PA, and AM, the peak of COVID-19 mortality coincided with the peaks of mortality from cardiovascular, respiratory, and ill-defined diseases. Moreover, in some states, the number of COVID-19 deaths was lower than excess deaths, resulting in a ratio of excess deaths to COVID-19 deaths greater than 1.

Patterns of excess deaths were heterogeneous among the Brazilian states. Excess deaths were notably higher in Northern, Center-Western, and Northeastern states, especially in AM, DF, and MT, indicating a more intense pandemic impact in these states in 2020. Meanwhile, the percentage of excess deaths was lower in Southern and Southeastern states, especially in RS, SC, and MG. These states tended to have ratios of excess deaths to COVID-19 deaths lower than 1, suggesting a decrease in deaths from other causes. This was quite evident in RS, which had a deficit of deaths in the second quarter of the year, suggesting that non-pharmacological measures were probably responsible for the observed reduction in deaths from external causes and other respiratory diseases, such as influenza, and for the delay in the peak of COVID-19^22^. Furthermore, the lower excess deaths in the Southern and Southeastern states may reflect the greater purchasing power of the population in these areas, which would greatly enhance their social distancing practices, including remote home work^23^.

Higher excess deaths were reported among males, those in the age group 20-59 years and of black, yellow, or indigenous race/color, suggesting that in Brazil, these groups were more vulnerable and had lesser chances of protecting themselves from contamination. For indigenous communities, this finding may be due to historical inequalities in access to health services and sanitary conditions, as well as a higher prevalence of malnutrition, infections, and chronic diseases^24^. For black and brown individuals, the higher excess deaths may have resulted from an increased exposure to COVID-19 associated with greater use of public transportation and insertion in informal and essential jobs requiring physical presence by these groups^25,26^.

A lower percentage of excess deaths was reported among women, those aged 80 years or more and in whites, suggesting that they were able to protect themselves more from COVID-19, with a greater adherence to social isolation and other non-pharmacological measures^27^. The ratio between excess deaths and COVID-19 deaths was less than 1 in these groups, wherein mortality rates from cardiovascular and respiratory diseases were reduced. Lower excess deaths in women can be explained by their lower insertion in the labor market, greater health care, greater adherence to the use of masks, and having fewer comorbidities associated with the aggravation of COVID-19 than men^28^. Regarding the lower percentage in whites, this was possibly attributed to the fact that they are wealthy and more likely to be employed in higher skilled occupations with a greater possibility of remote work^27^. Similarly, the elderly, many of whom are retired, were more likely to remain in social isolation and thus safer from COVID-19^29^.

There was a deficit of deaths in those under 20 years of age, which was explained by a lower COVID-19 mortality^30^ and large reduction in deaths from respiratory diseases and external causes in this age group. This reduction in deaths from these causes was also observed in other countries in 2020 among individuals below 15 years of age^31^. This deficit has been attributed to a combination of factors. For one, children and adolescents had to spend more time at home with the suspension of face-to-face learning^32^. Another reason was that the restricted mobility caused by the quarantine measures likely led to a reduction in unintentional (drowning and car accidents) and intentional injuries (homicides)^14,33,34^, as well as deaths from influenza^10^.

Interestingly, mortality from external causes in 2020 was lower in those younger than 20 years of age and in males. This was most likely due to the effect of social distancing in Brazil, which reduced firearm and stabbing injuries during the pandemic^35^, in addition to a drop of approximately 22% in deaths related to traffic accidents in São Paulo^36^.

In Southern and Southeastern states, such as MG, RS, RJ, SP, and PR, a reduction in the mortality rates for cardiovascular diseases and neoplasms were observed in those ≥ 40 years of age and in whites. Reasons for this reduction have yet to be studied, but it may be related to the substitution of causes of death^37^. Additionally, individuals at a higher risk of death from cardiovascular diseases and neoplasms may have died earlier from COVID-19, since these patients would present with a worse prognosis upon contracting COVID-19^38^.

Despite the findings of this study, certain limitations were noted. For one, SIM data for the year 2020 were preliminary and may not represent all deaths that occurred in the period, as it is subject to corrections, especially in the underlying cause of death. However, these changes in data are expected to be minimal in the future since most of 2020 deaths have already been recorded.

Nevertheless, we highlight some strengths in this study. First, the use of the SIM database made it possible to process detailed information on the basic cause of death and perform analysis by place of residence. Another strong point was the inclusion of deaths from external causes, which allowed the identification of an accentuated drop in violent deaths among men, those under 19 years of age, and those between 20 and 39 years of age.

In conclusion, the impacts of COVID-19 on mortality were heterogeneous among Brazilian states and in sex, age, and race/color groups, as well as modified the mortality structure in 2020 relative to the previous five years. These different impacts of COVID-19 on mortality in certain states and groups revealed geographic, demographic, socioeconomic, and racial inequalities that expose individuals in different ways to the risk of infection and mortality from COVID-19 or other causes. Furthermore, the underreporting of COVID-19 deaths and the increase of mortality from ill-defined causes reflect the shortcomings of the death surveillance system, reinforcing the need for greater funding and strengthening of this system in Brazil.

## Supporting information

Supplementary Material - additional description of methods and supplementary figures

## Data Availability

The R scripts for all analyses performed in this study can be accessed at the author's github.
Records regarding non-fetal deaths from the years 2015 to 2020 were obtained from the national Mortality Information System (SIM) available at the website opendatasus.saude.gov.br
Demographic/population data were obtained from the Brazilian Institute of Geographics and Statistics (IBGE).

https://github.com/aamouradasilva/excess_mortality.git

https://www.ibge.gov.br/

https://www.opendatasus.saude.gov.br

## Acknowledgments

We thank FAPEMA (Research and Scientific and Technological Development Support Foundation of Maranhão) for the article publication funding program. We thank CAPES (Coordination for the Improvement of Higher Education Personnel) [finance code No: 001].

We thank CNPq (National Council for Scientific and Technological Development) for the research productivity grants: Process: 306592/2018-5.

