## Supplementary Material - additional description of methods and supplementary figures for "EXCESS DEATHS FROM ALL CAUSES AND BY COVID-19 IN BRAZIL IN 2020"

10/08/2021

#### Supplementary Material

##### Methods

The Farrington algorithm is designed to detect outliers in univariate time series. To accomplish this, it considers an overdispersed Poisson generalized linear model (Quasi-Poisson) to determine a predictive distribution for the observed values of the series in a non-surge scenario. Considering the predictive distribution, a specific quantile is obtained, which will be used as a threshold to determine whether the observed value at time $t$ is an outlier. If the observed value is above this threshold, then an alarm is triggered. [1]

For execution of the algorithm, the time series of the number of deaths in the period from 2010 to 2020 for each Brazilian state was partitioned into two parts. The first partition of the series was composed with the data from week 1 of 2010 (i.e., January 3, 2010) to the last week of 2014 (December 28, 2014), corresponding to 260 epidemiological weeks. These data were used for fitting the quasi-Poisson model. The data from the first week of 2015 (January 4, 2015) until week 52 of 2020 (December 13, 2020) were considered for monitoring, totaling 312 weeks. Thus, using only values from the period 2010 to 2014 for model fitting, prediction intervals are obtained for the observed values for 2015 to 2020.

For detection of outliers in the series from 2015 to 2020, the quantile 0.995 of the predictive distribution of the observed values of the series was adopted as the threshold. The Farrington algorithm is implemented in the surveillance package of the R program, and the function farrington should be used for its execution. [2]

Supplementary figures 46, 47, and 48 present the detected alarms for the death series for some brazilian states where specific patterns of alarms were observed in some weeks of 2016 - RN, PE, AL, PB, RJ, SP, PR and RS states. For comparison purposes, we fitted the model specified in equation (1), described in the methods section of the main article, with the series from 2015 to 2019 with the weeks that showed anomalies in the year 2016 and without the inclusion of these weeks. Although the differences were statistically significant, the relative differences between the estimates of the number of expected deaths for the epidemiological weeks of the year 2020 calculated based on the 2015-2019 series with and without those anomalies were small (<0.7%). Thus, we chose to consider the complete series of the year 2016 for the estimation of expected deaths.

### Supplementary Tables

**Supplementary Table 1. Number of expected deaths and its lower and upper 95% confidence limits by state, Brazil, 2020**

| state | expected | lower__limit__expected | upper__limit__expected |
|---|---|---|---|
| RO | 8380 | 8118 | 8656 |
| AC | 4313 | 4122 | 4518 |
| AM | 18574 | 18097 | 19052 |
| RR | 3106 | 2944 | 3277 |
| PA | 42048 | 41224 | 42877 |
| AP | 3679 | 3507 | 3863 |
| TO | 8263 | 7990 | 8537 |
| MA | 35556 | 34794 | 36323 |
| PI | 21514 | 20838 | 22210 |
| CE | 58570 | 57326 | 59839 |
| RN | 22078 | 21500 | 22682 |
| PB | 27379 | 26717 | 28054 |
| PE | 63602 | 61886 | 65358 |
| AL | 20050 | 19531 | 20581 |
| SE | 13251 | 12913 | 13596 |
| BA | 93854 | 92469 | 95266 |
| MG | 141695 | 139184 | 144252 |
| ES | 24852 | 24312 | 25396 |
| RJ | 146253 | 143401 | 149165 |
| SP | 308807 | 303618 | 314077 |
| PR | 75237 | 73778 | 76723 |
| SC | 43261 | 42427 | 44112 |
| RS | 90237 | 88112 | 92417 |
| MS | 17135 | 16710 | 17572 |
| MT | 18779 | 18300 | 19267 |
| GO | 41265 | 40350 | 42199 |
| DF | 12816 | 12478 | 13170 |
| Brazil | 1364603 | 1350846 | 1378510 |

**Supplementary Table 2. Number of expected deaths and its lower and upper 95% confidence limits by sex, age and race, Brazil, 2020**

| Category | expected | lower_limit_expected | upper_limit_expected |
|---|---|---|---|
| Male | 752451 | 744860 | 760123 |
| Female | 611671 | 604904 | 618505 |
| Age_00_to_19 | 63628 | 62720 | 64539 |
| Age_20_to_39 | 102251 | 100011 | 104535 |
| Age_40_to_59 | 242597 | 240052 | 245164 |
| Age_60_to_79 | 528174 | 522349 | 534062 |
| Age_80_or_older | 418733 | 412727 | 424832 |
| White | 692066 | 683479 | 700760 |
| Black | 110258 | 108855 | 111680 |
| Brown | 524683 | 519541 | 529879 |
| Others | 12077 | 11787 | 12373 |

continue...

### Supplementary Figures

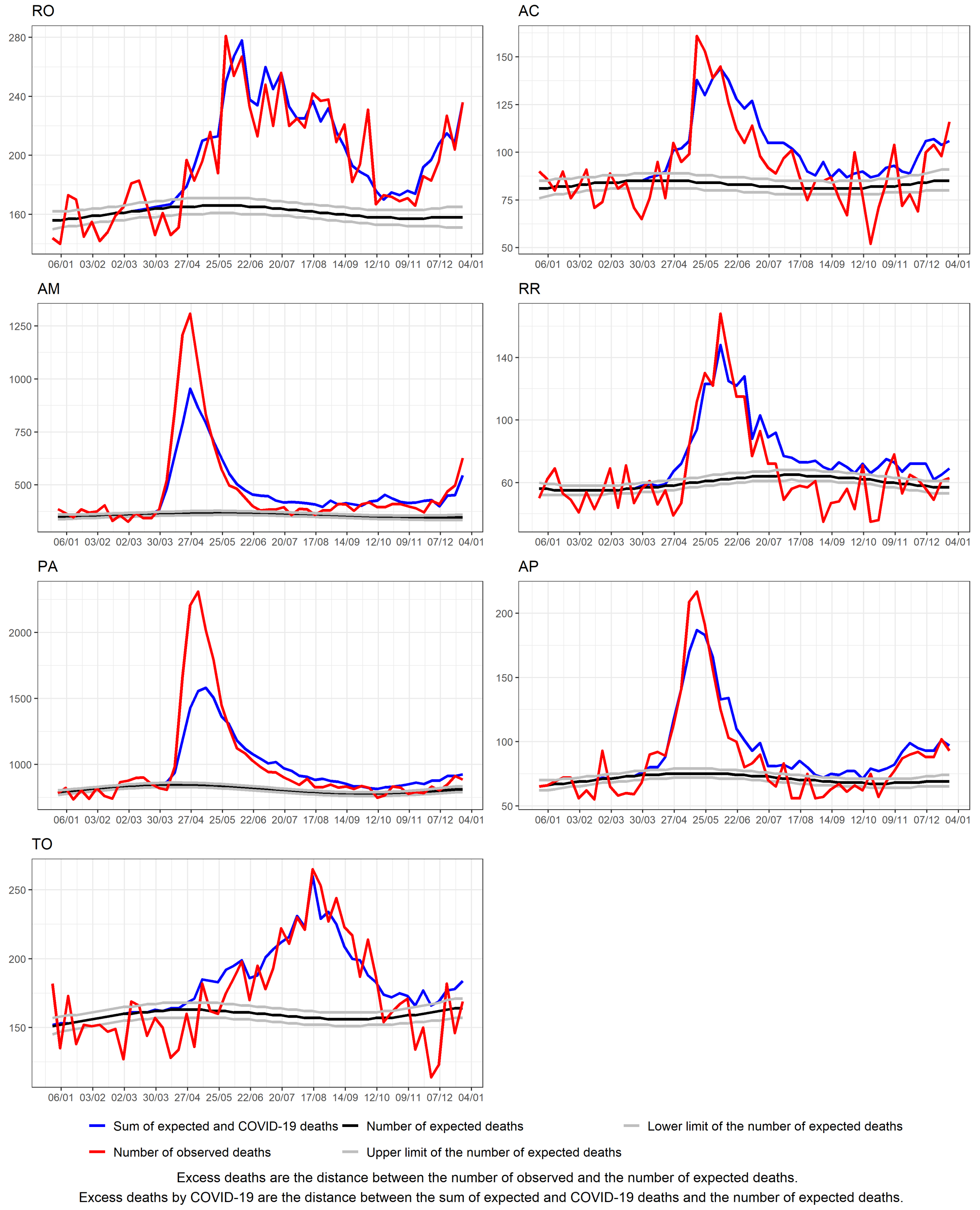


Supplementary figure 1. Excess deaths by all causes and by COVID=19 by epidemiological week, North Region, Brazil, 2020

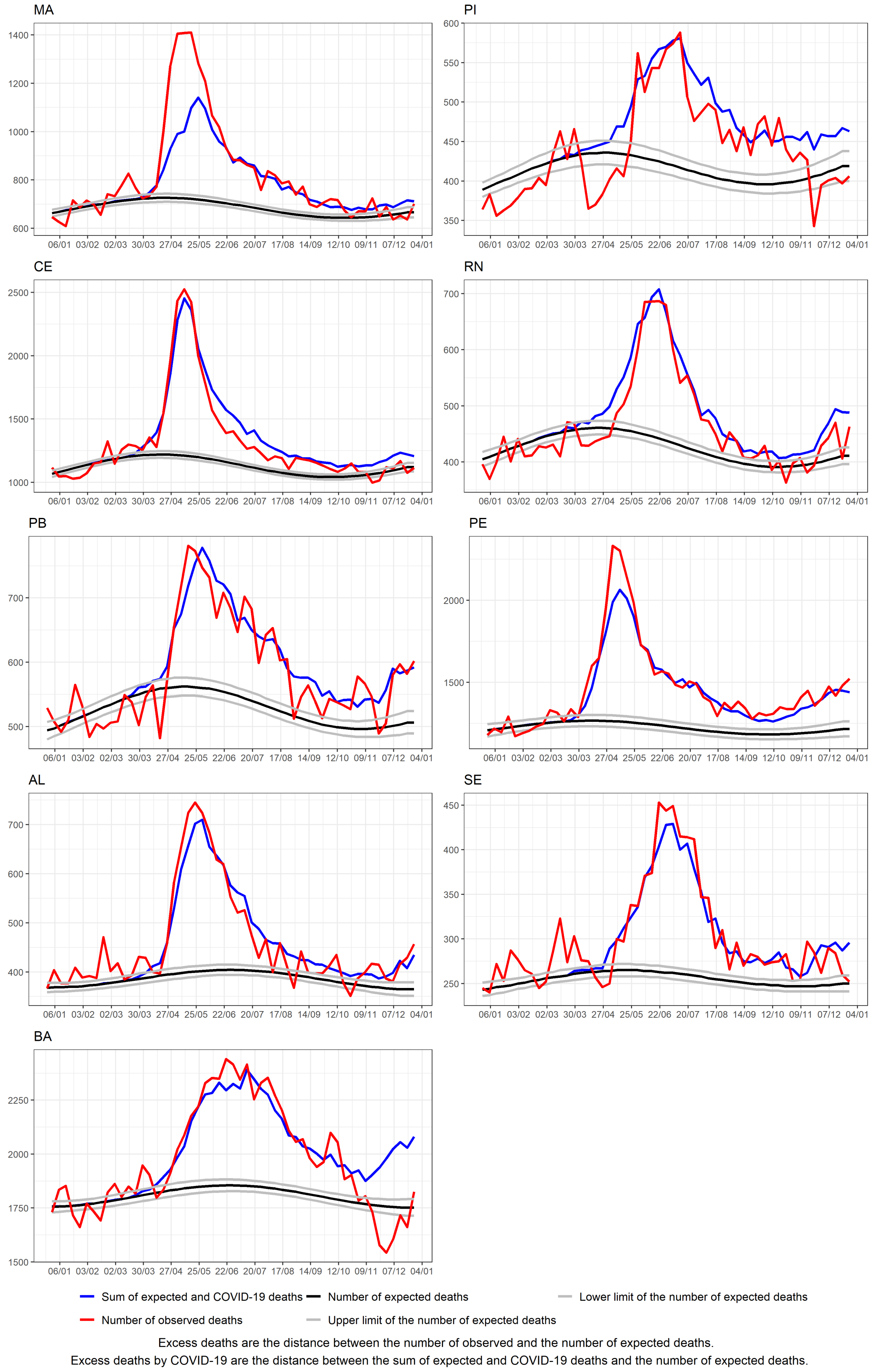


Supplementary figure 2. Excess deaths by all causes and by COVID=19 by epidemiological week, Northeast Region, Brazil, 2020

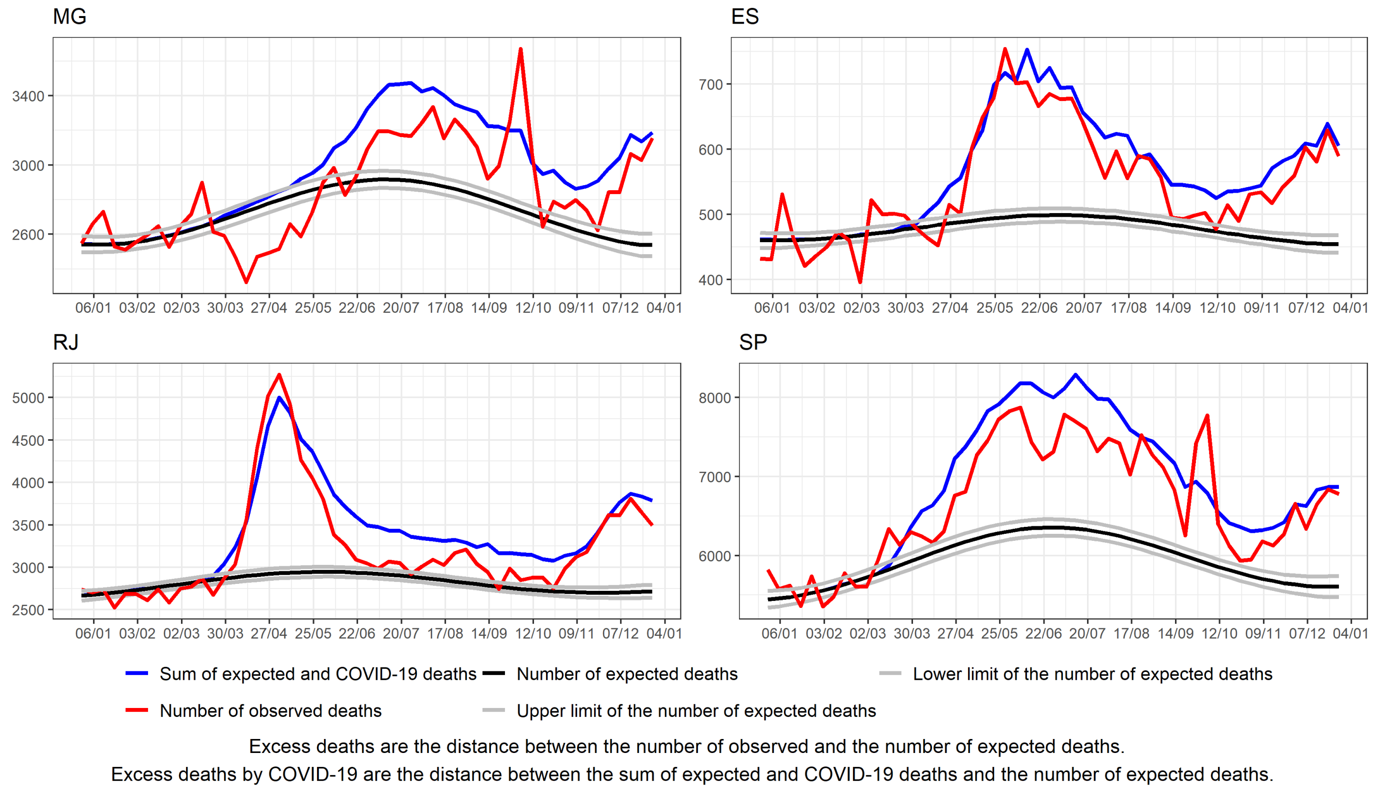


Supplementary figure 3. Excess deaths by all causes and by COVID=19 by epidemiological week, Southeast Region, Brazil, 2020

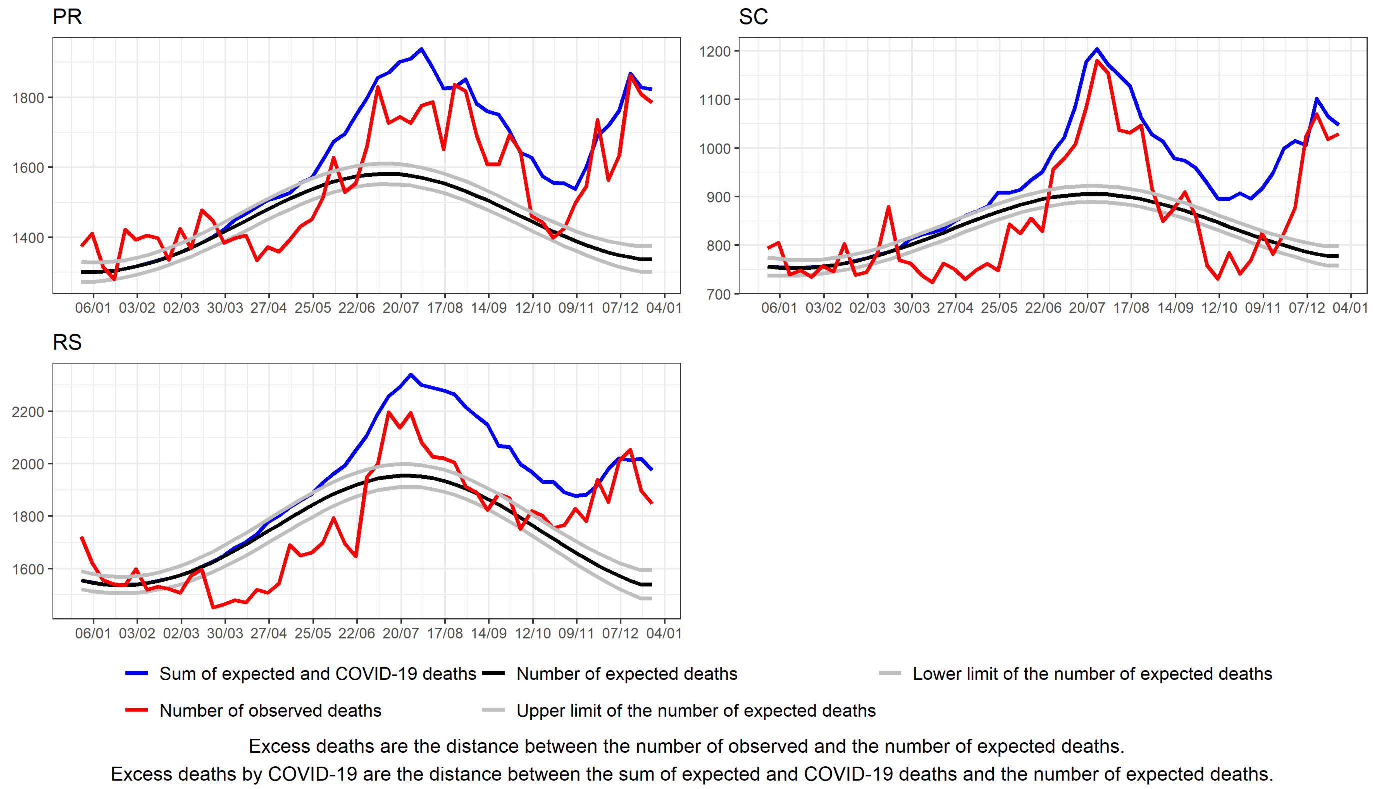


Supplementary figure 4. Excess deaths by all causes and by COVID=19 by epidemiological week, South Region, Brazil, 2020

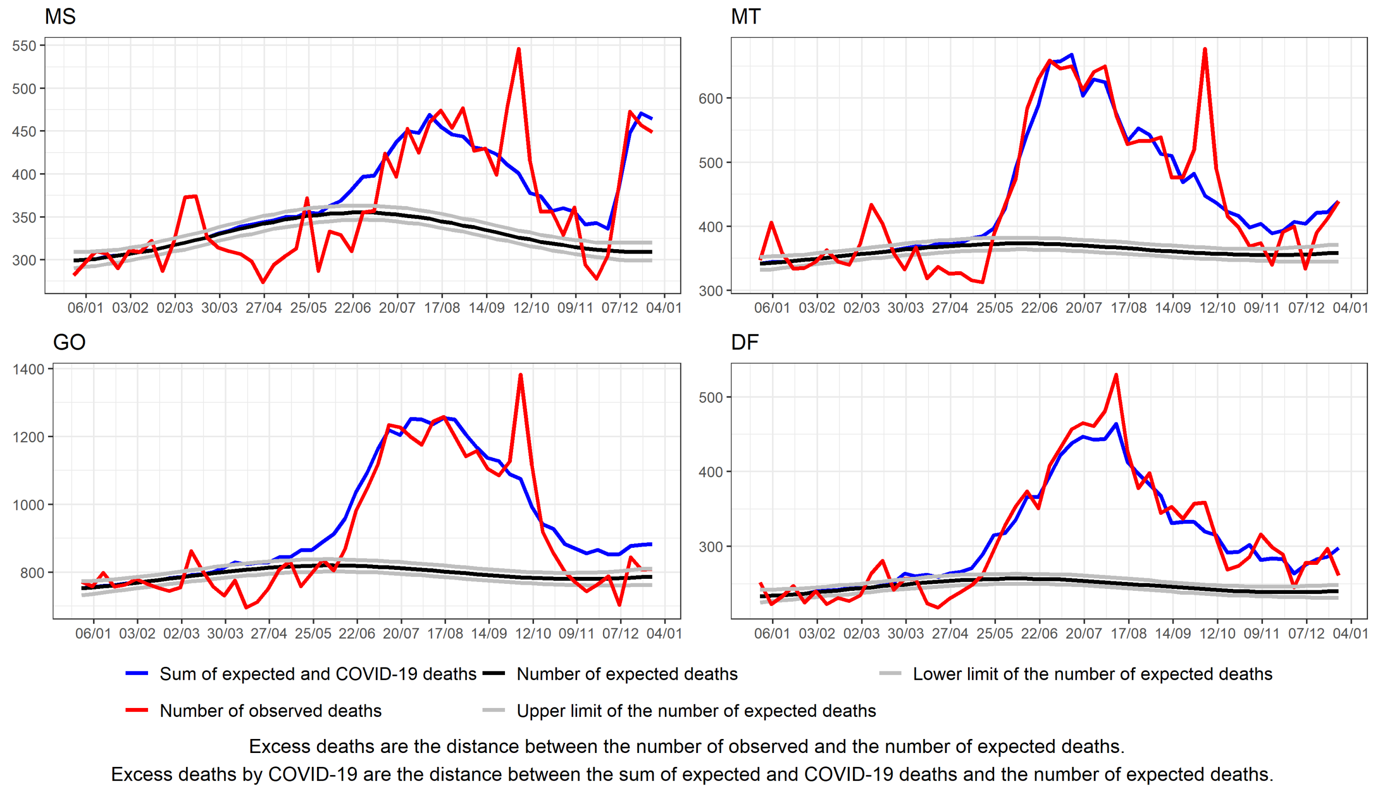


Supplementary figure 5. Excess deaths by all causes and by COVID=19 by epidemiological week, Center-West Region, Brazil, 2020

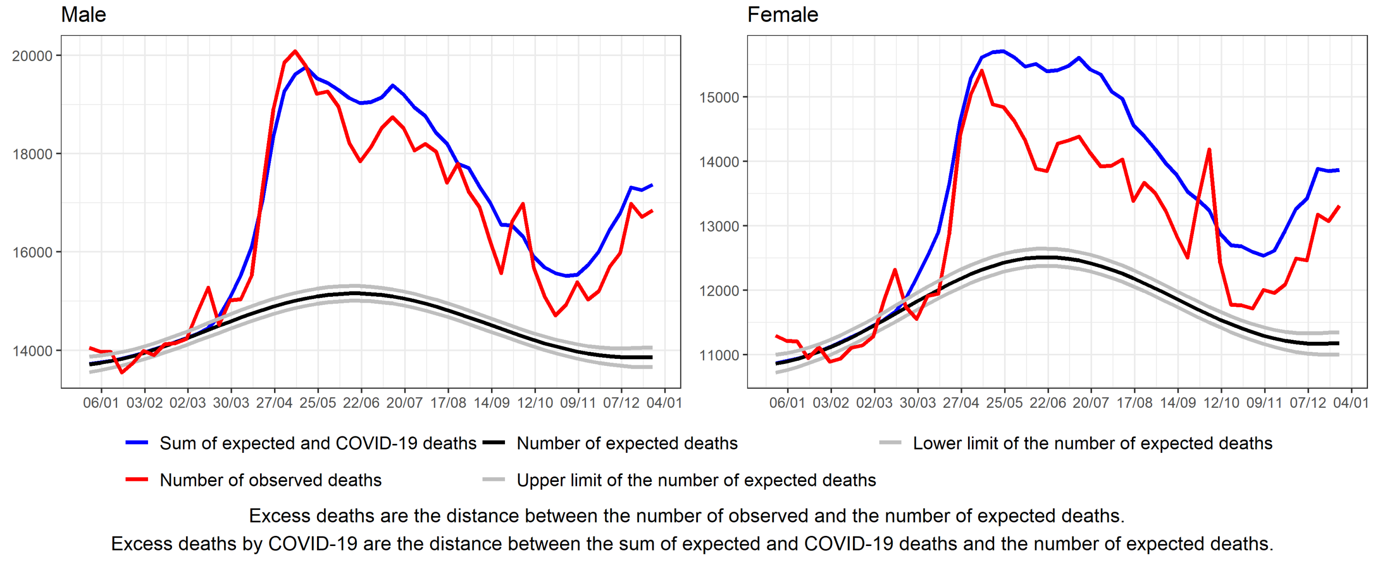


Supplementary figure 6. Excess deaths by all causes and by COVID=19 by epidemiological week according to sex, Brazil, 2020

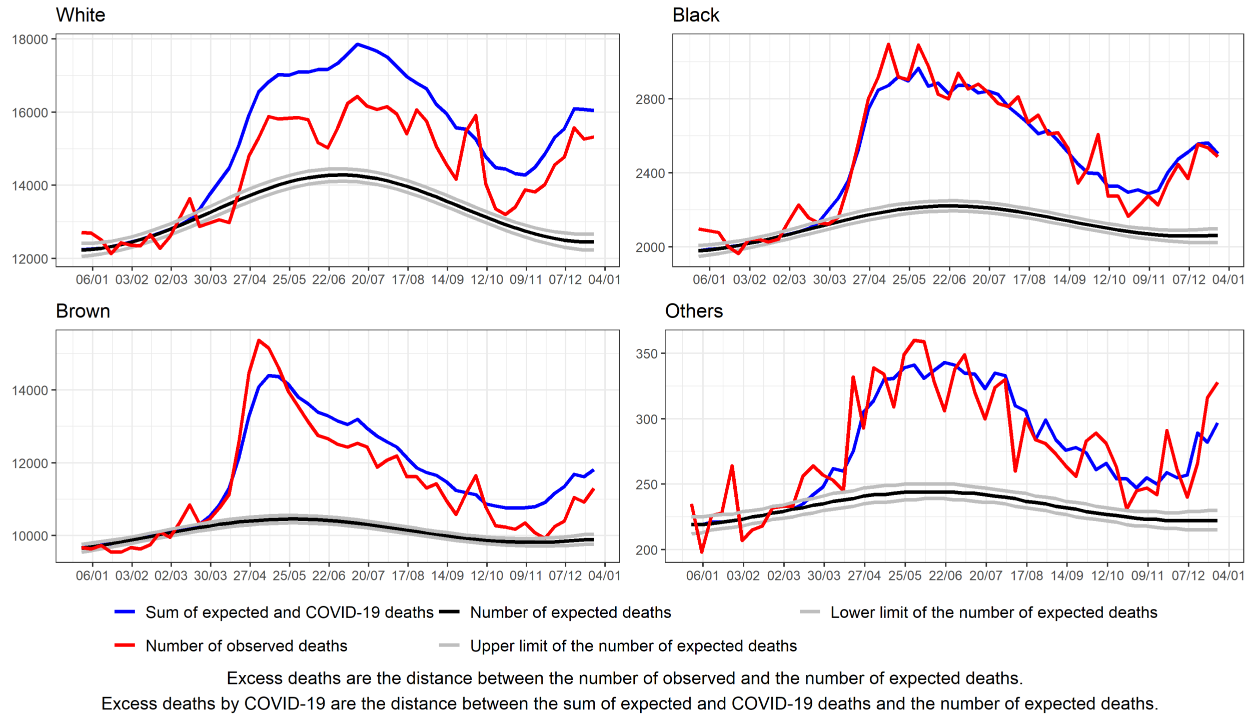


Supplementary figure 7. Excess deaths by all causes and by COVID=19 by epidemiological week according to race, Brazil, 2020

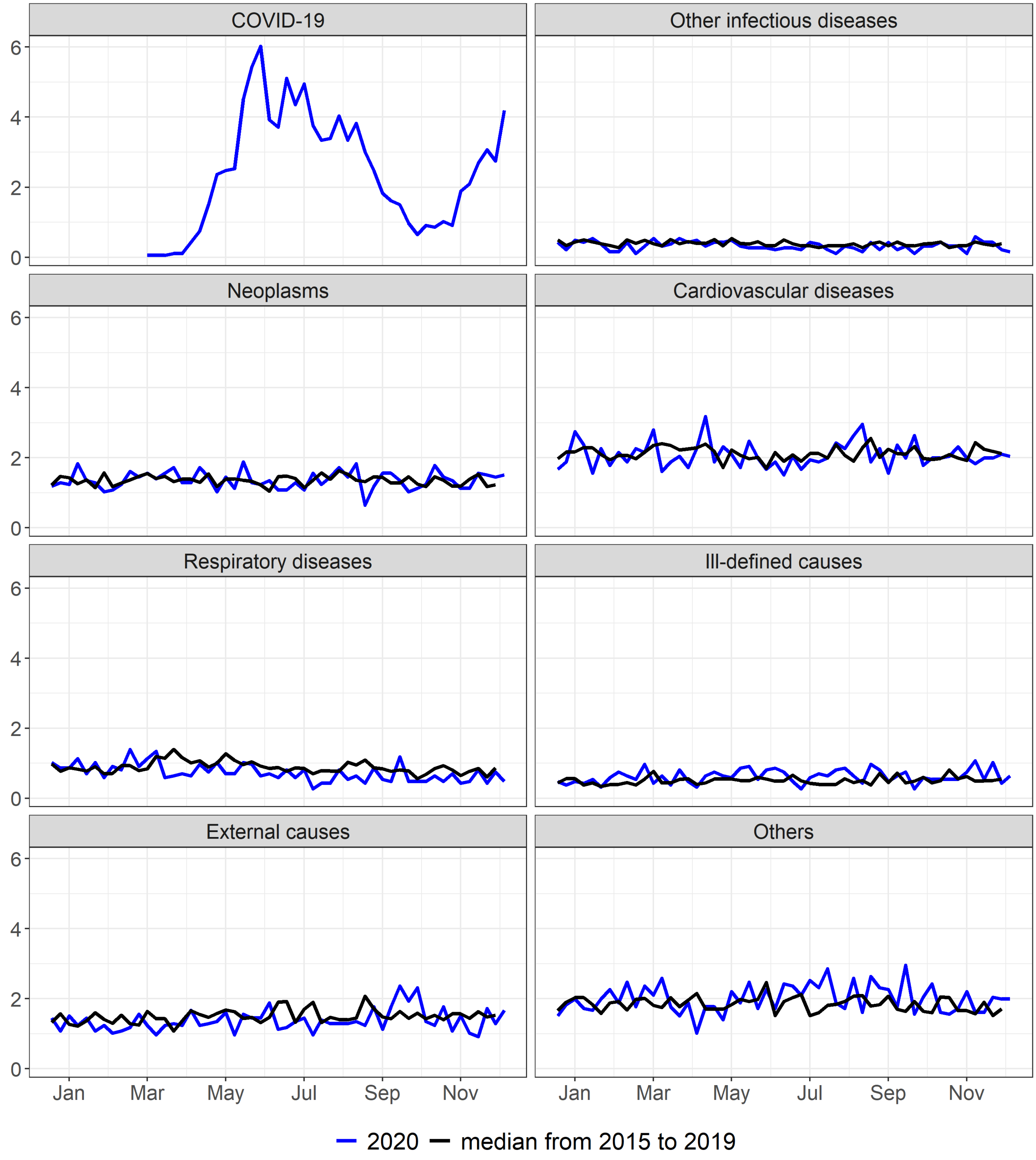


Supplementary figure 8. Mortality rate (per 100.000) by epidemiological week according to selected causes, RO, 2015 to 2020

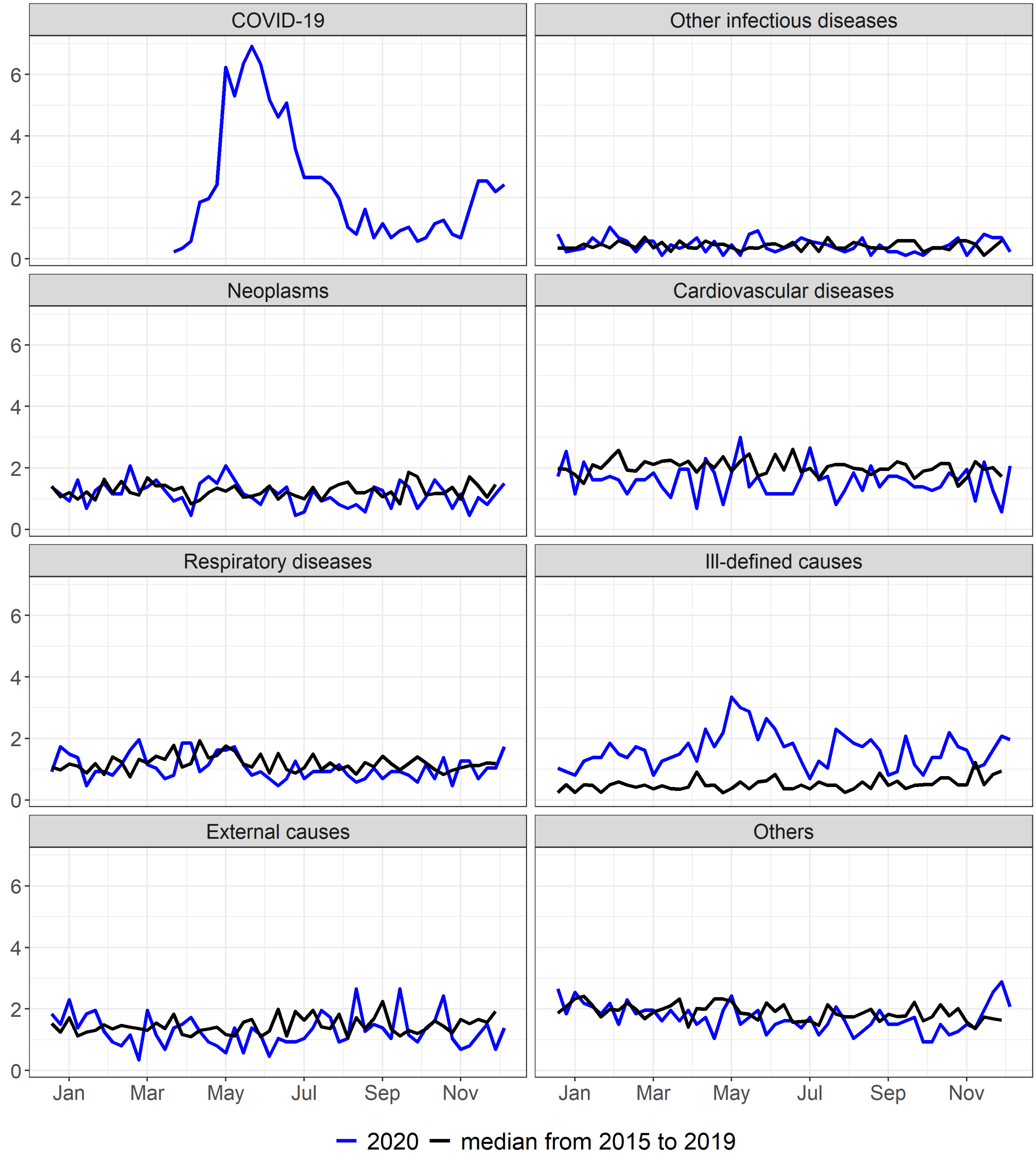


Supplementary figure 9. Mortality rate (per 100.000) by epidemiological week according to selected causes, AC, 2015 to 2020

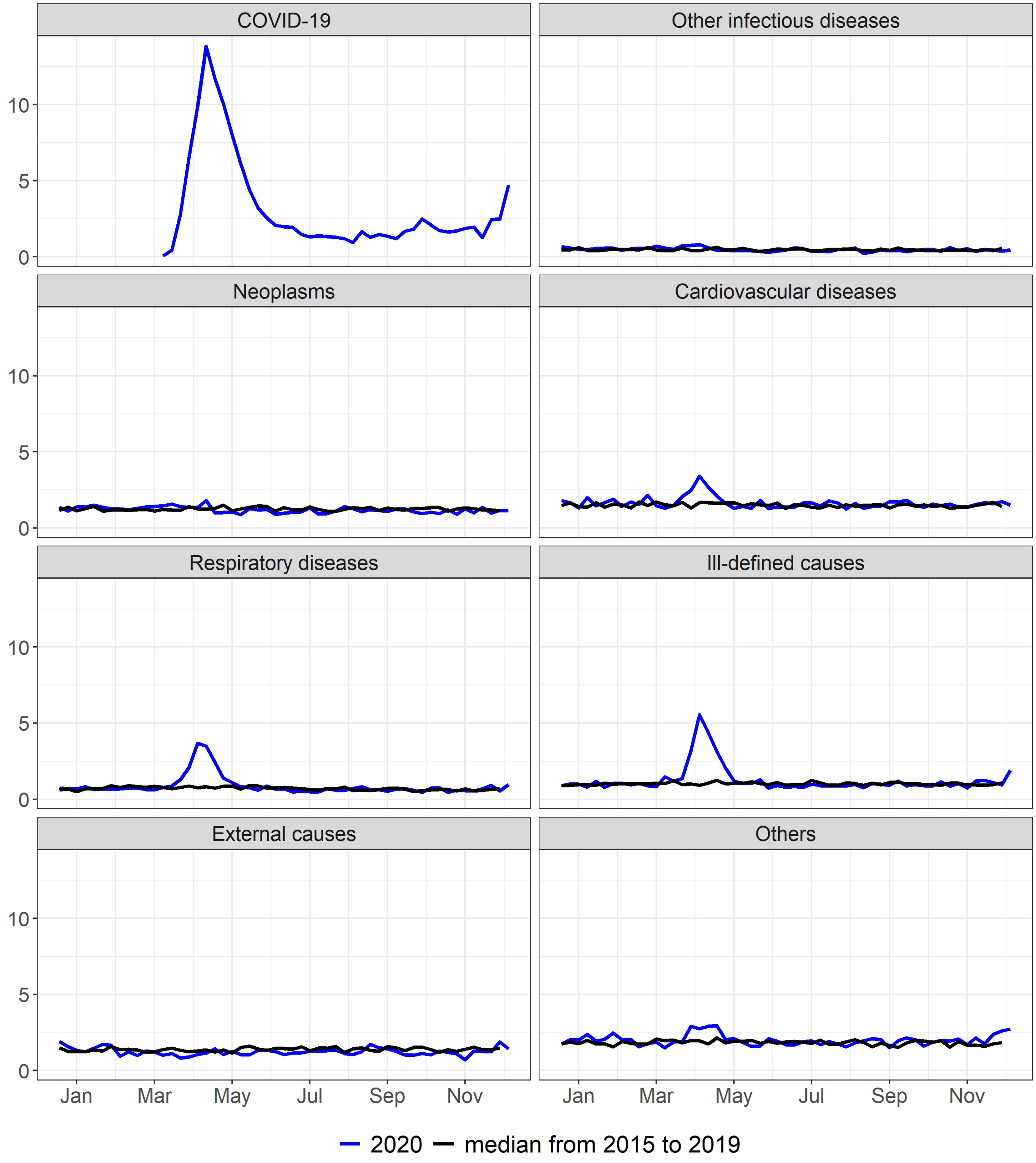


Supplementary figure 10. Mortality rate (per 100.000) by epidemiological week according to selected causes, AM, 2015 to 2020

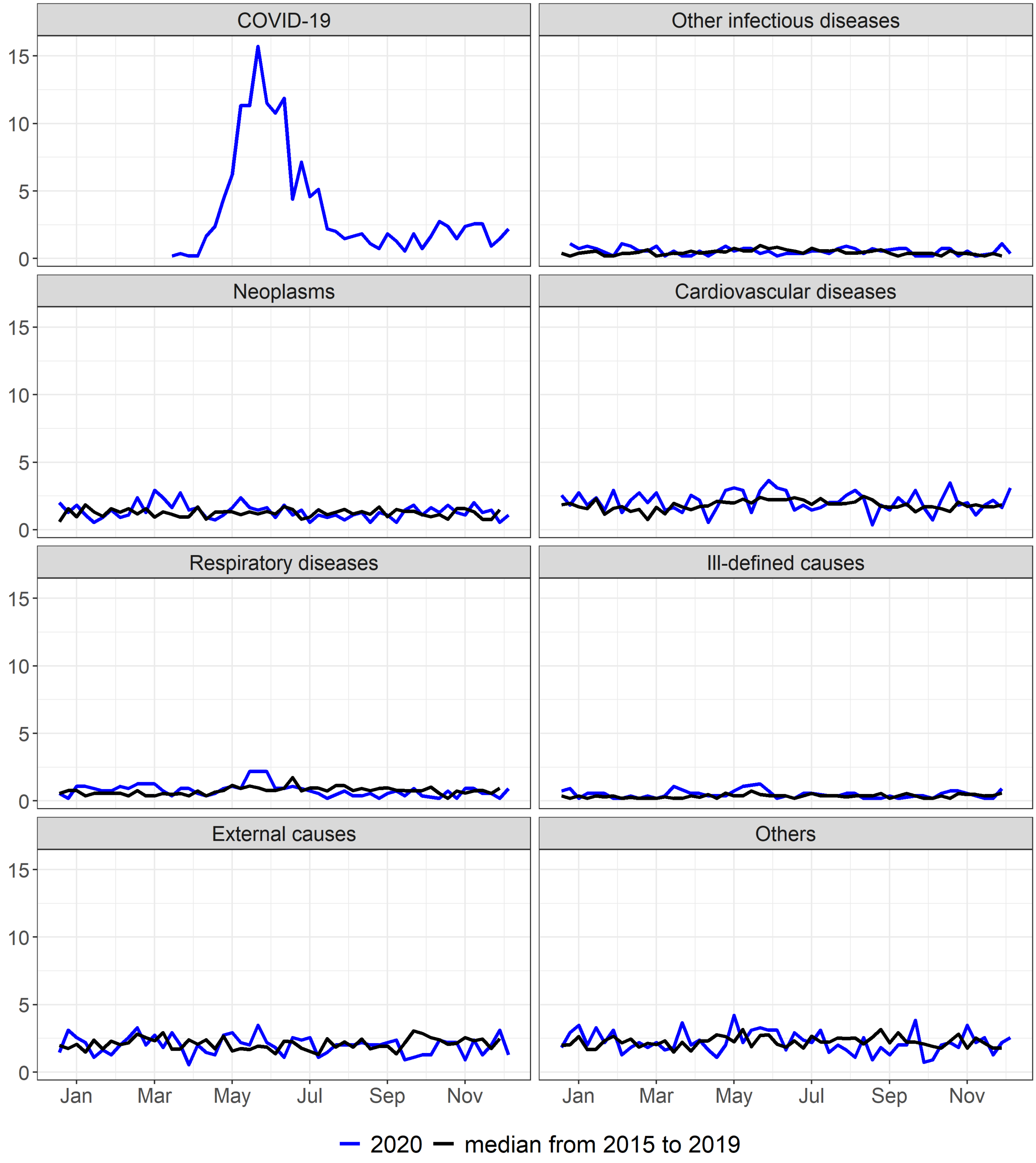


Supplementary figure 11. Mortality rate (per 100.000) by epidemiological week according to selected causes, RR, 2015 to 2020

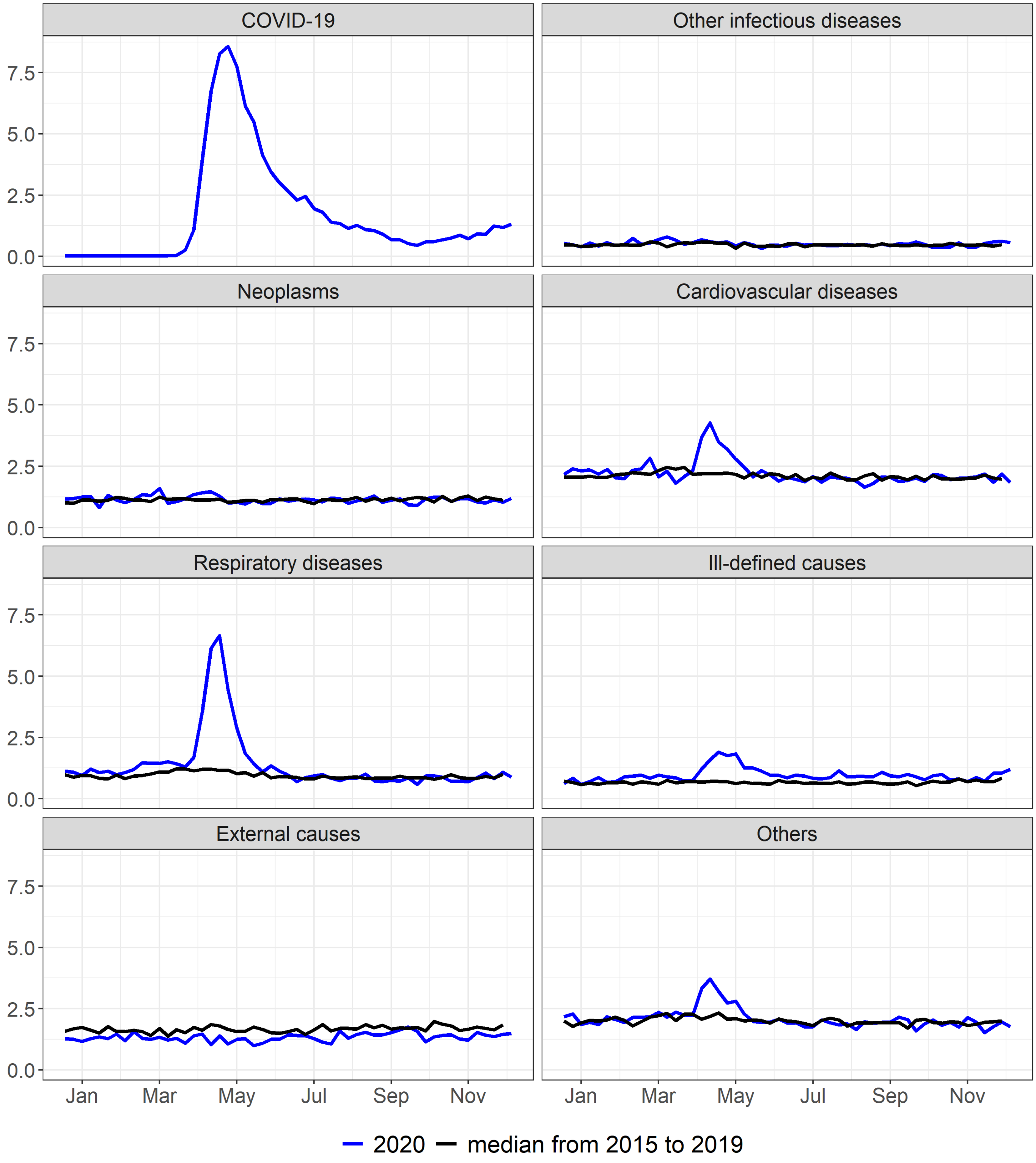


Supplementary figure 12. Mortality rate (per 100.000) by epidemiological week according to selected causes, PA, 2015 to 2020

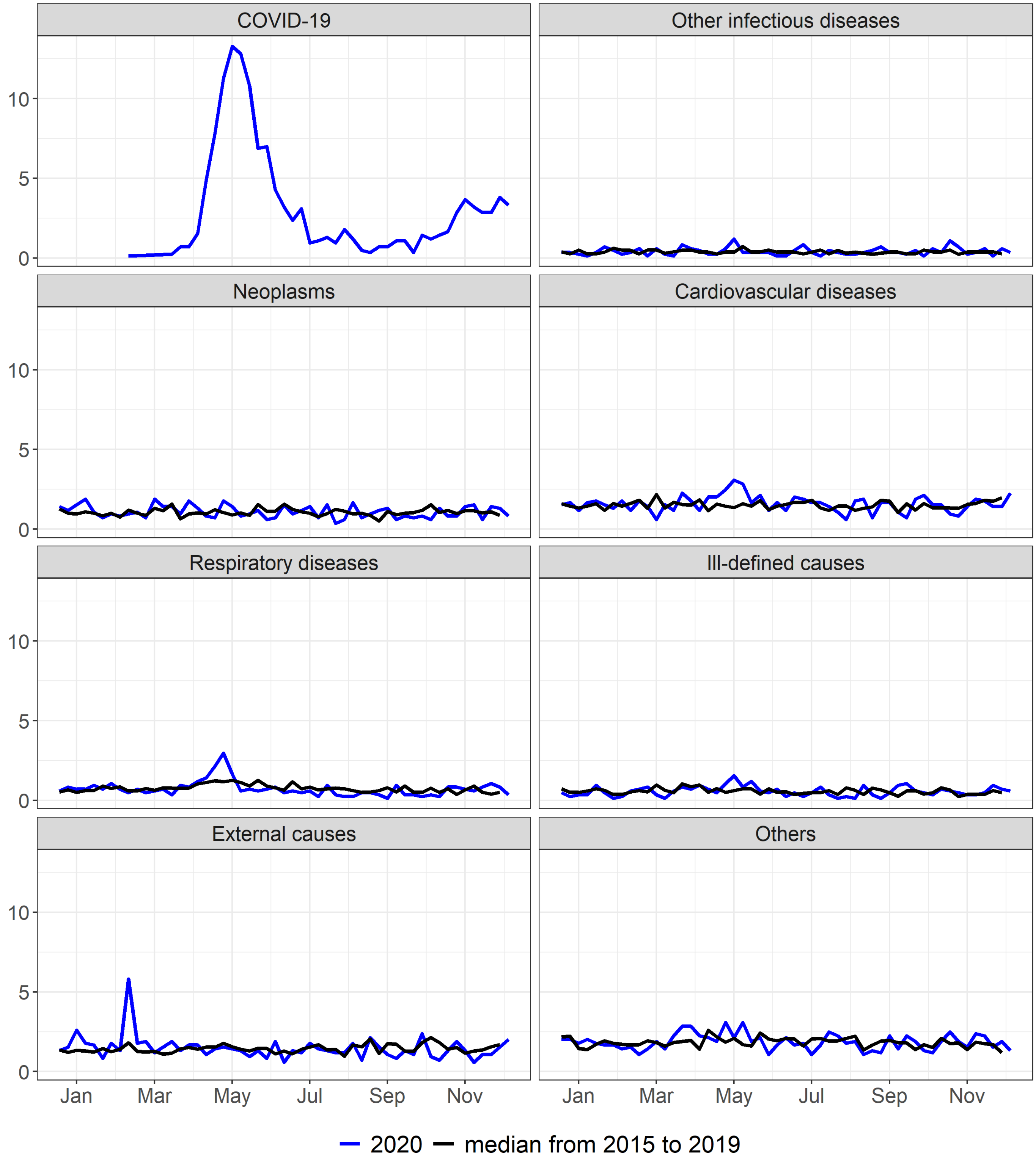


Supplementary figure 13. Mortality rate (per 100.000) by epidemiological week according to selected causes, AP, 2015 to 2020

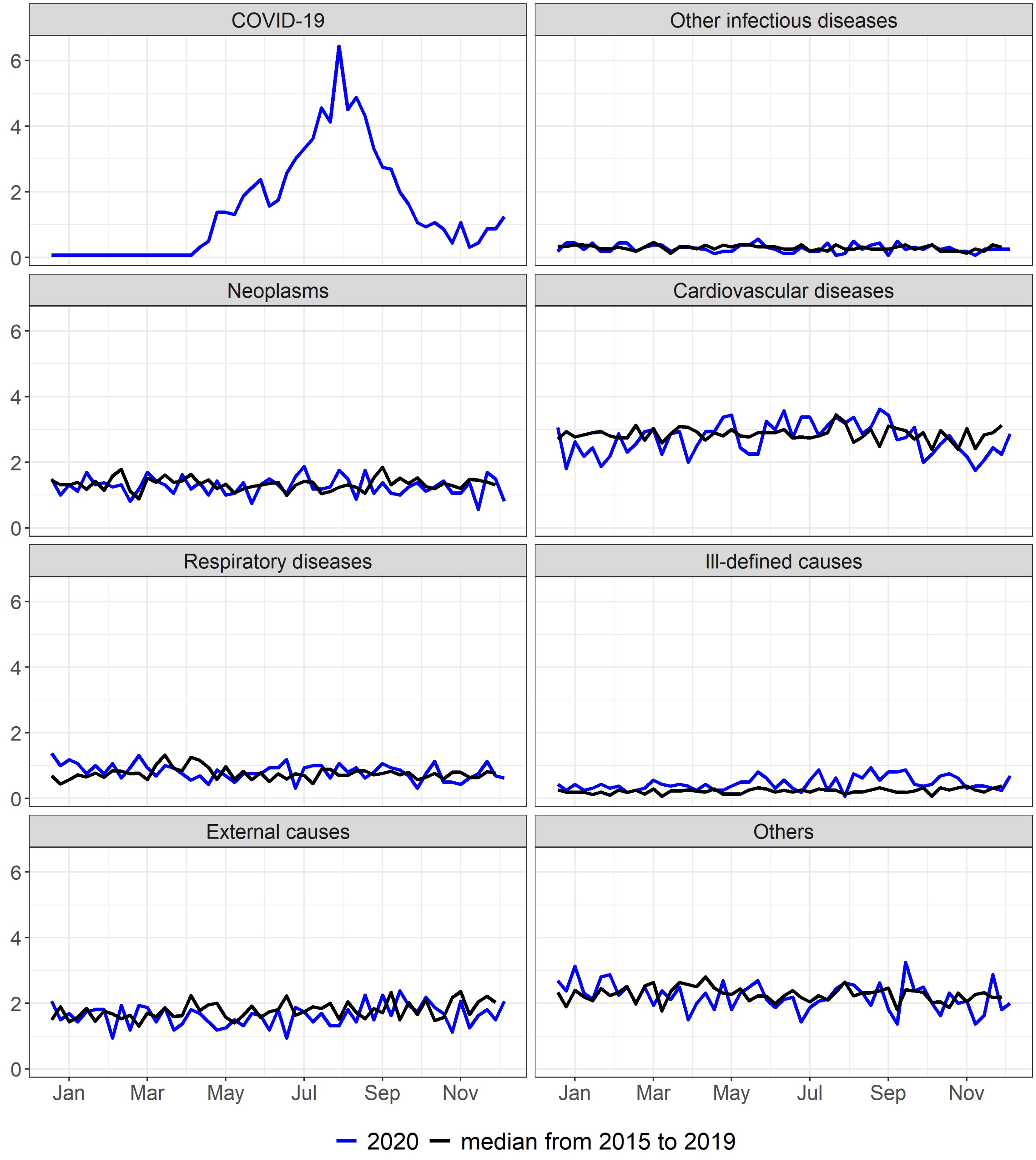


Supplementary figure 14. Mortality rate (per 100.000) by epidemiological week according to selected causes, TO, 2015 to 2020

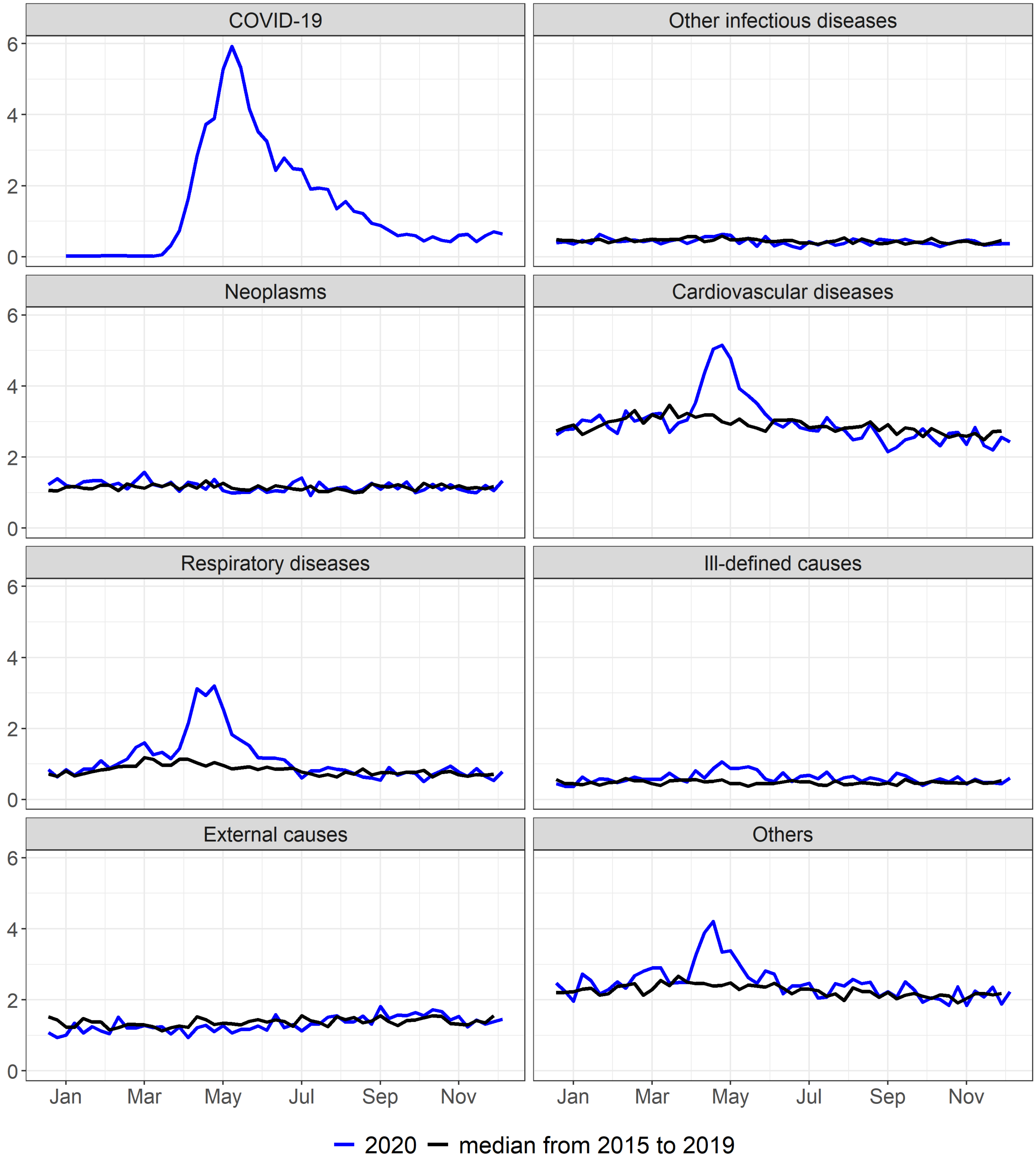


Supplementary figure 15. Mortality rate (per 100.000) by epidemiological week according to selected causes, MA, 2015 to 2020

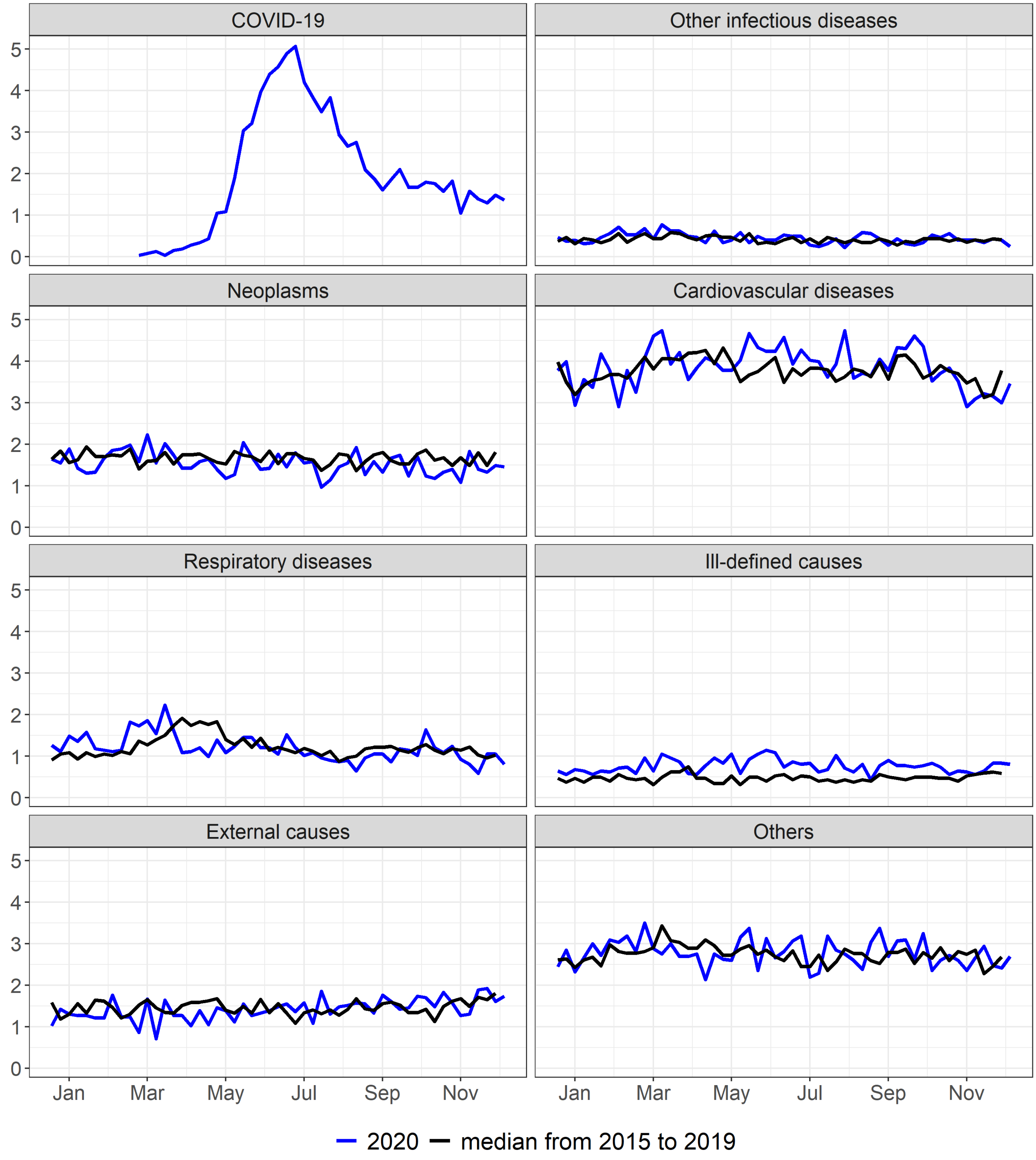


Supplementary figure 16. Mortality rate (per 100.000) by epidemiological week according to selected causes, PI, 2015 to 2020

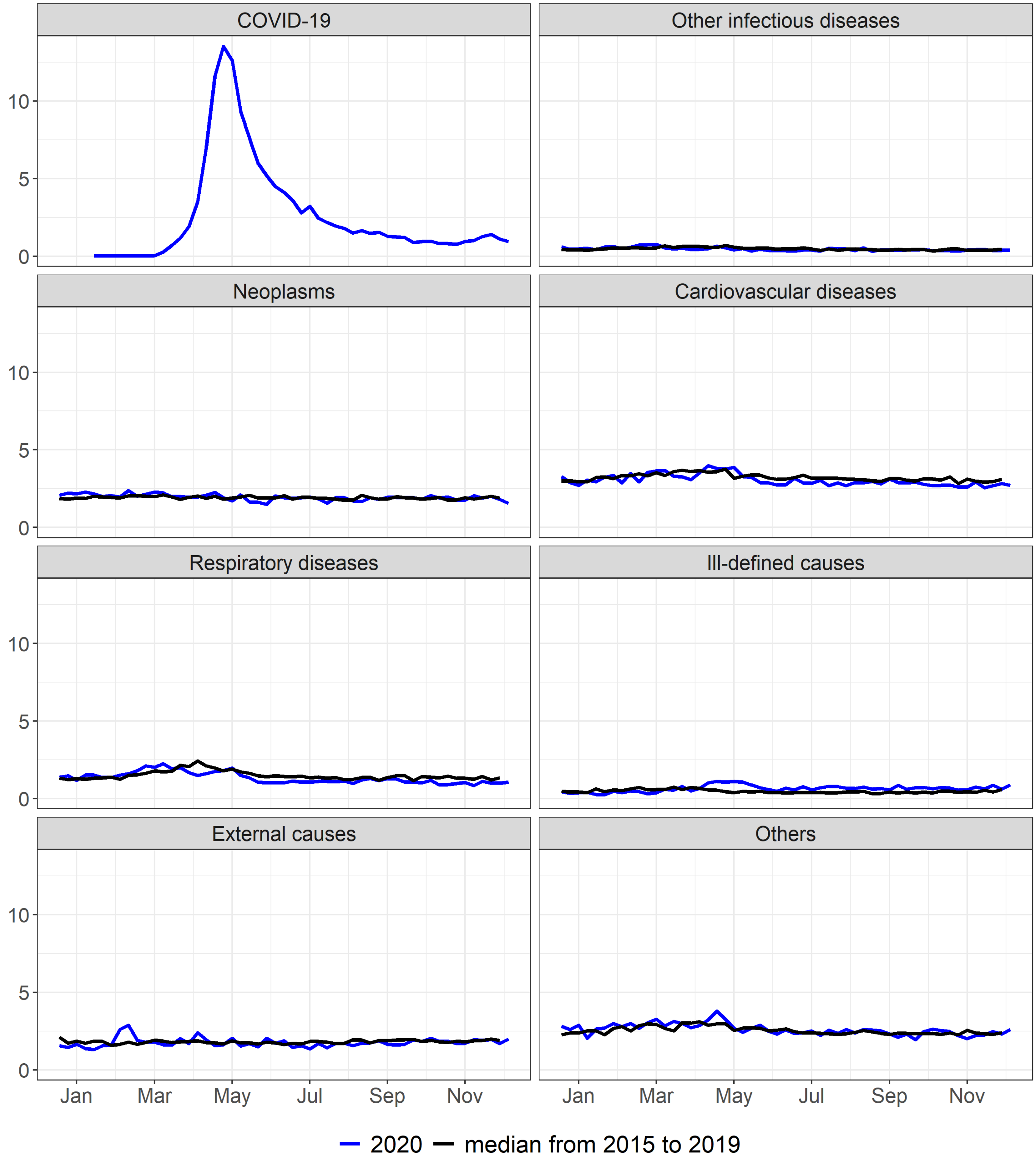


Supplementary figure 17. Mortality rate (per 100.000) by epidemiological week according to selected causes, CE, 2015 to 2020

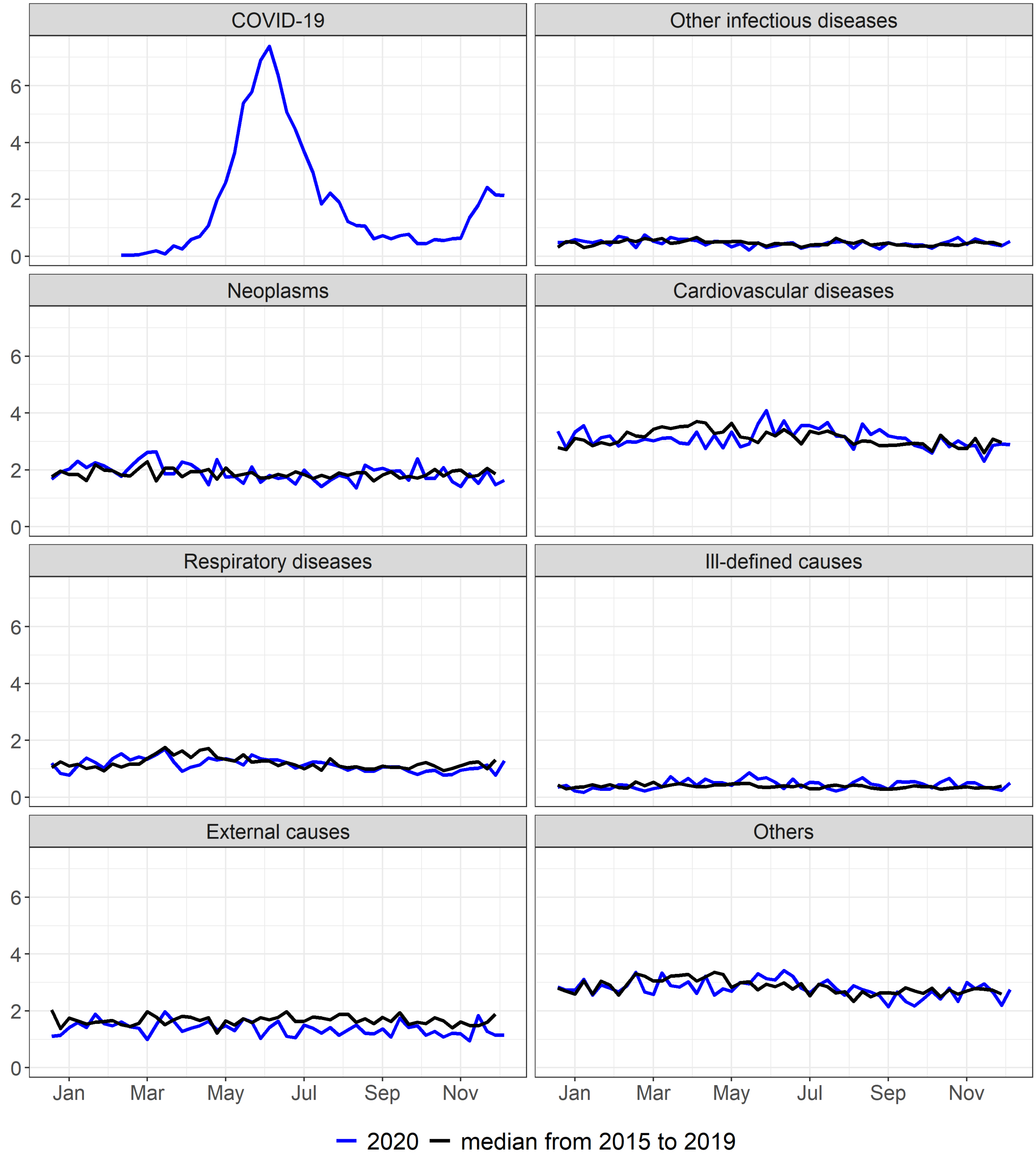


Supplementary figure 18. Mortality rate (per 100.000) by epidemiological week according to selected causes, RN, 2015 to 2020

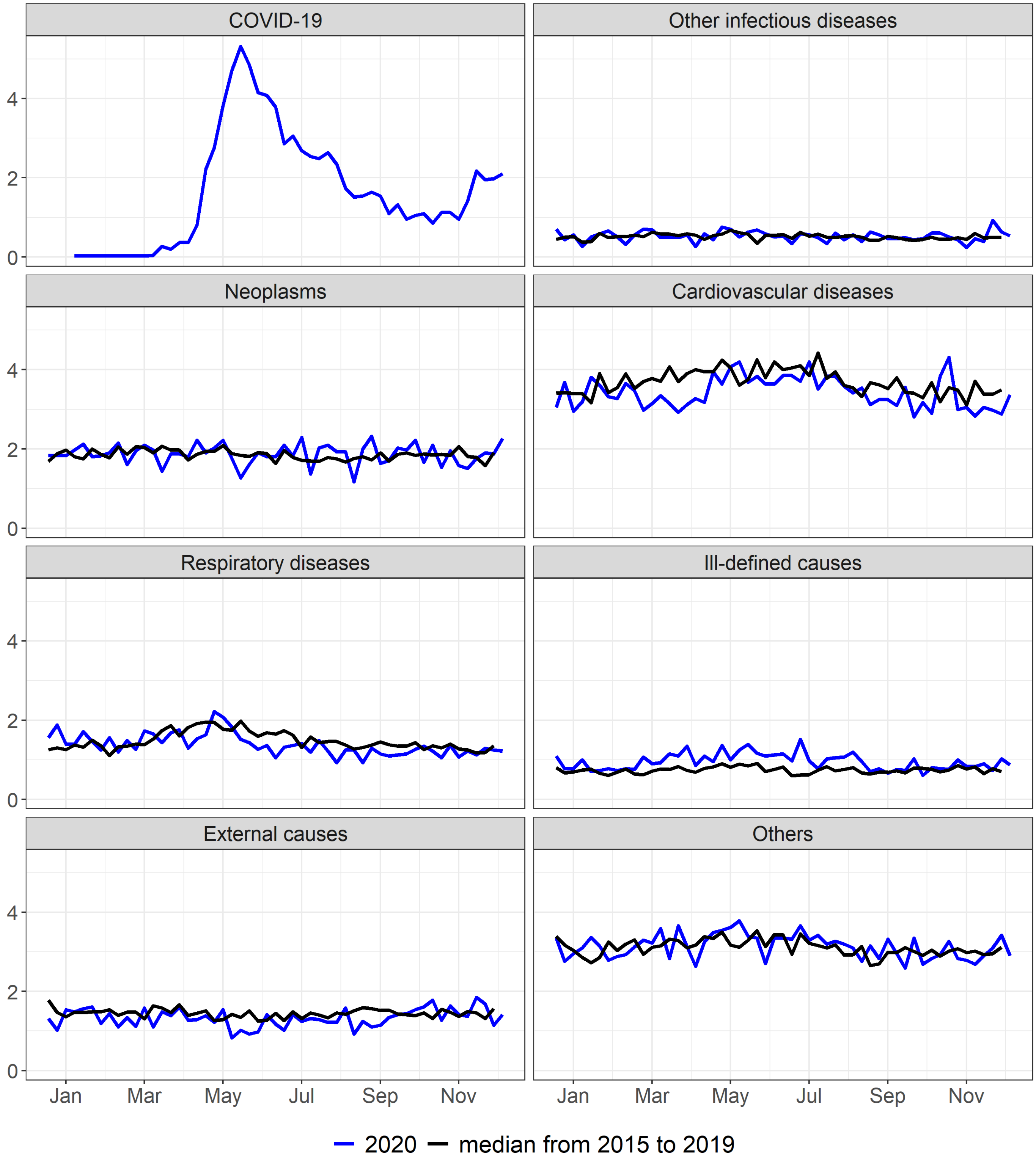


Supplementary figure 19. Mortality rate (per 100.000) by epidemiological week according to selected causes, PB, 2015 to 2020

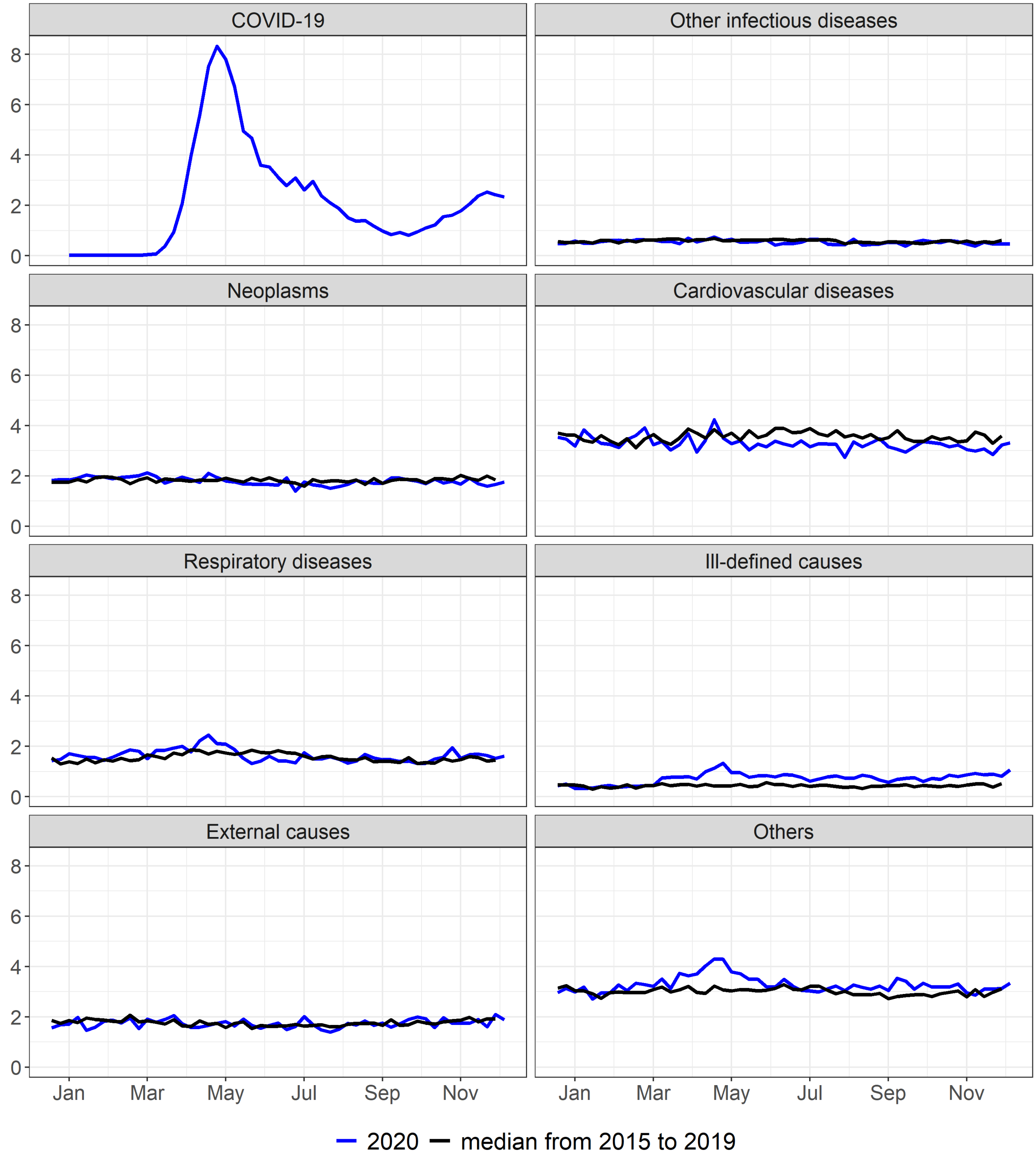


Supplementary figure 20. Mortality rate (per 100.000) by epidemiological week according to selected causes, PE, 2015 to 2020

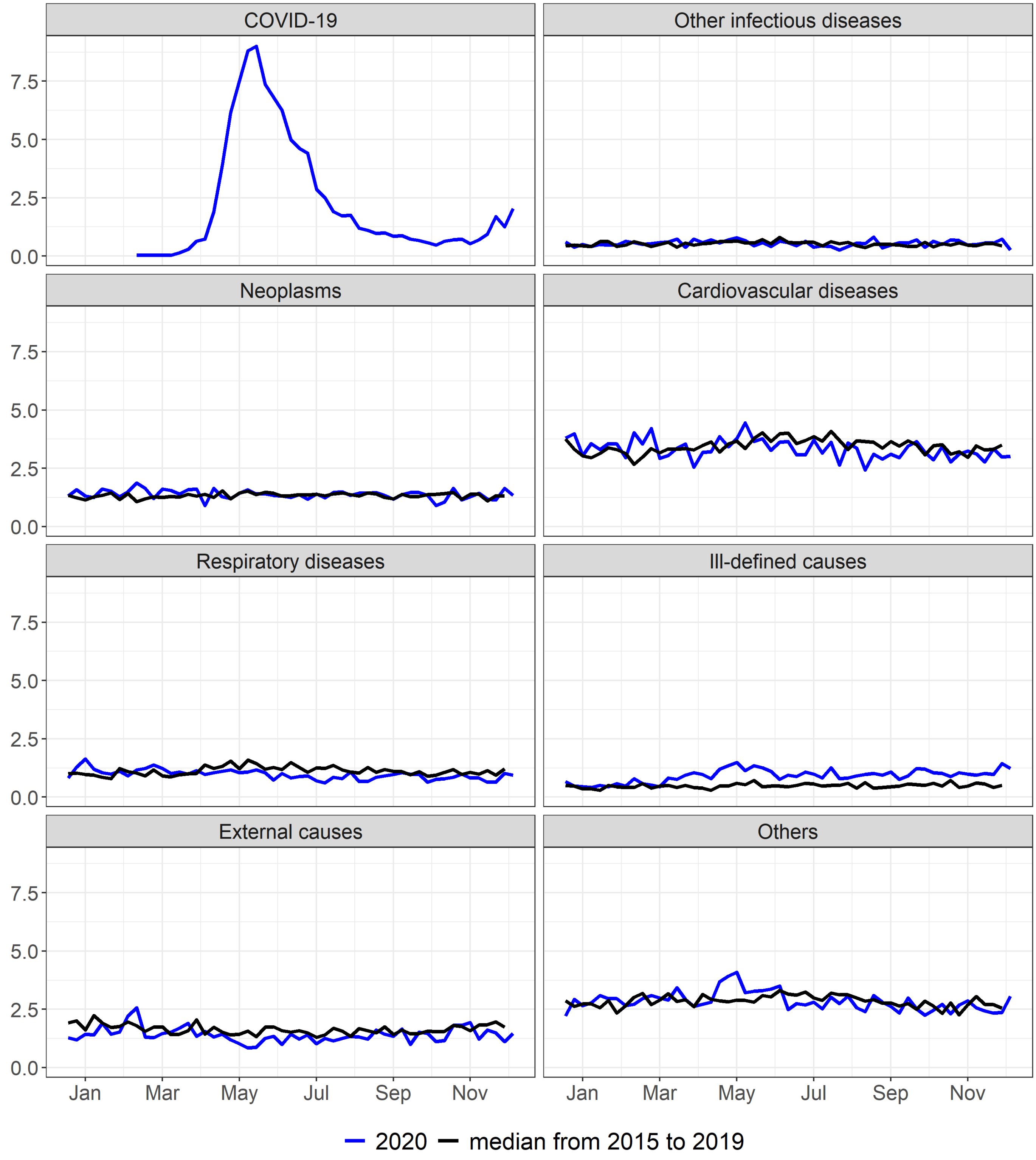


Supplementary figure 21. Mortality rate (per 100.000) by epidemiological week according to selected causes, AL, 2015 to 2020

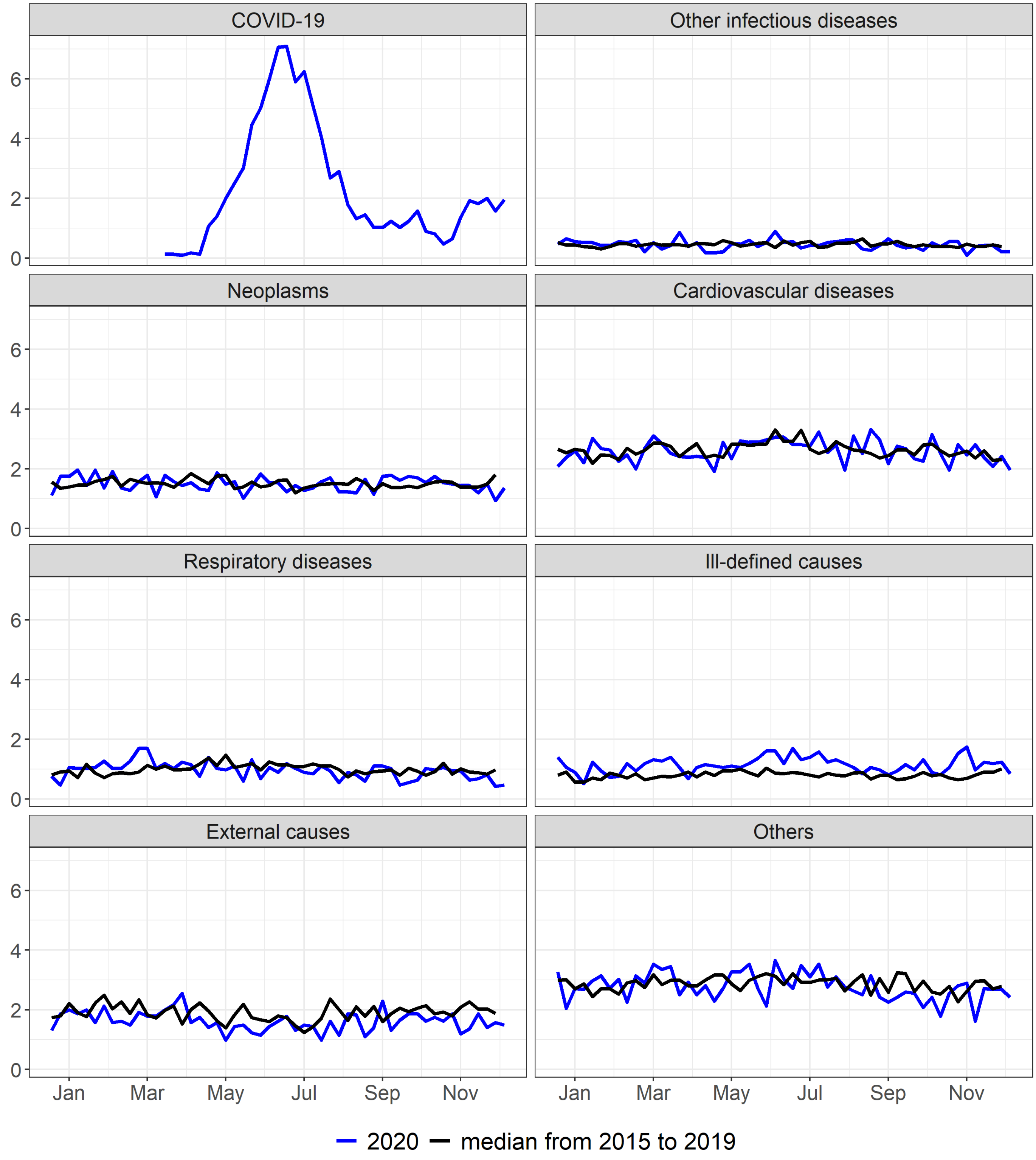


Supplementary figure 22. Mortality rate (per 100.000) by epidemiological week according to selected causes, SE, 2015 to 2020

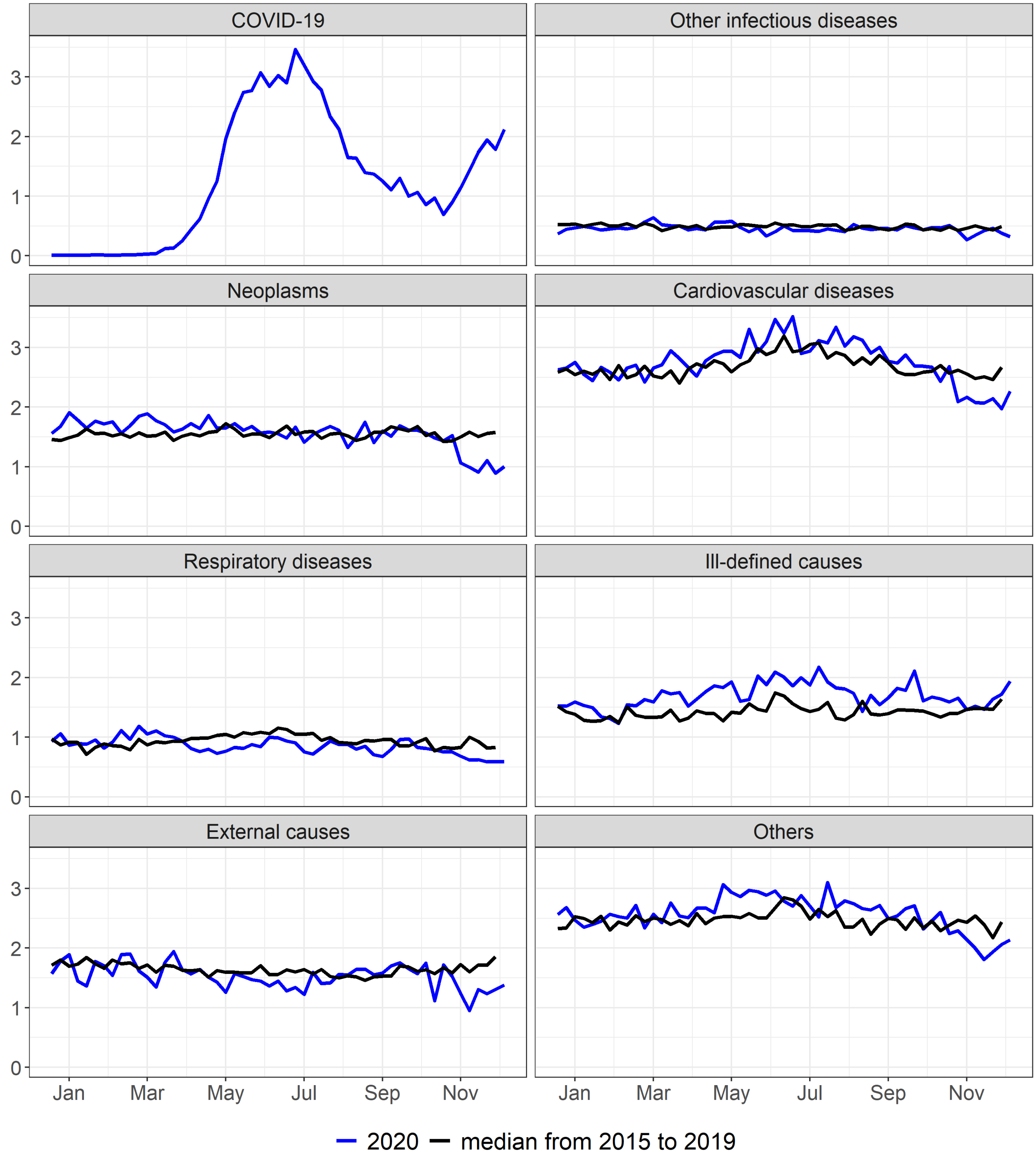


Supplementary figure 23. Mortality rate (per 100.000) by epidemiological week according to selected causes, BA, 2015 to 2020

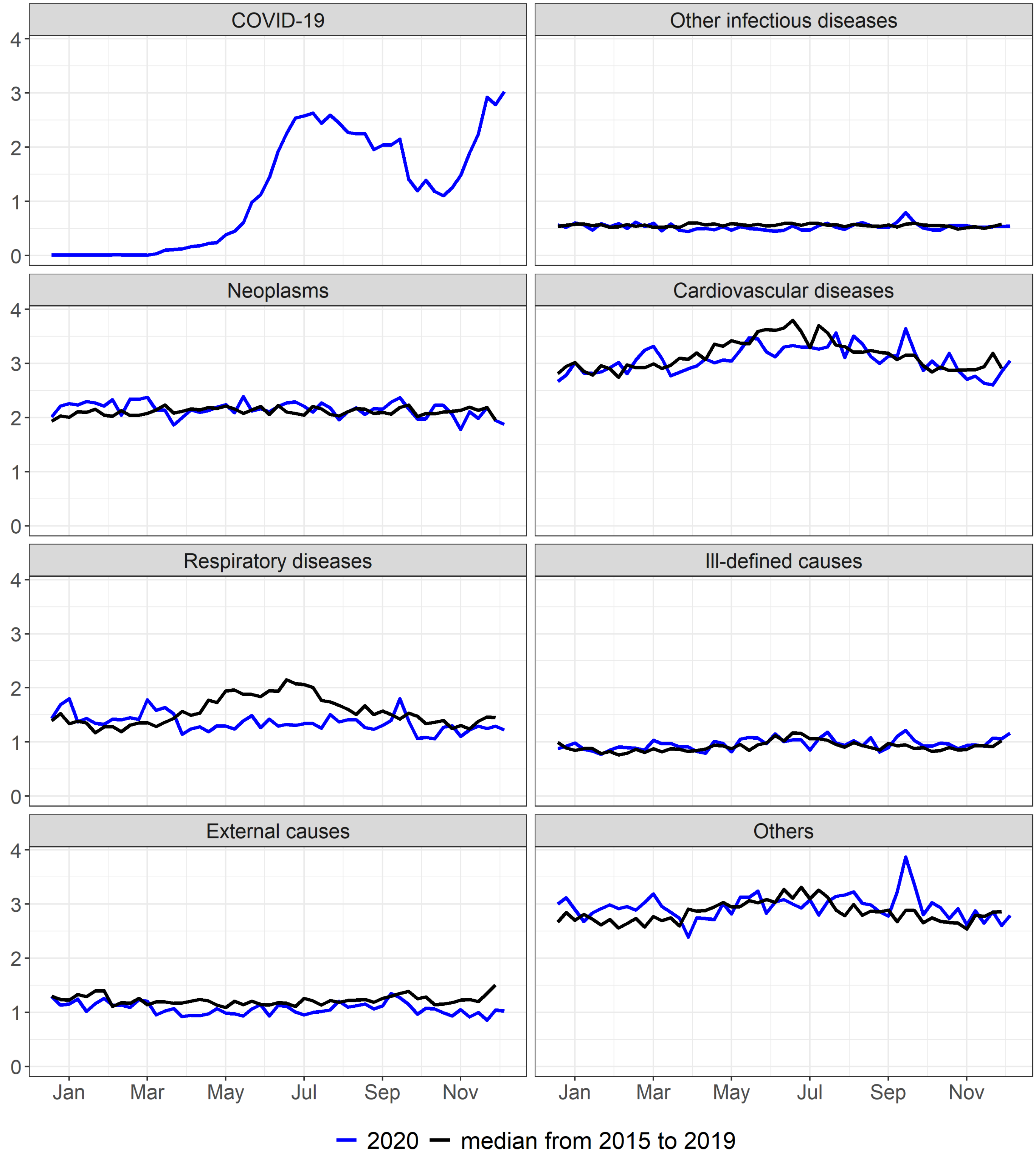


Supplementary figure 24. Mortality rate (per 100.000) by epidemiological week according to selected causes, MG, 2015 to 2020

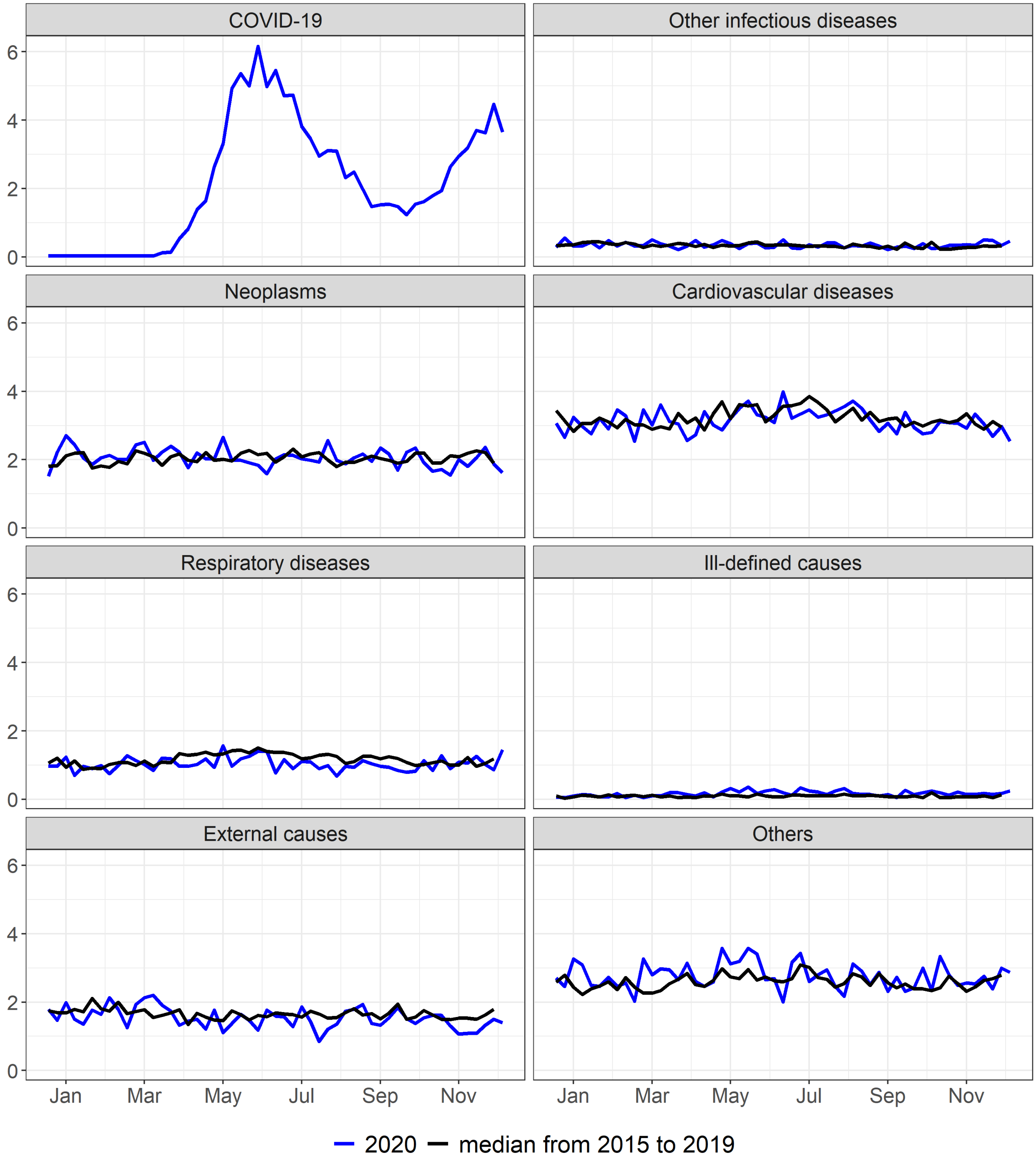


Supplementary figure 25. Mortality rate (per 100.000) by epidemiological week according to selected causes, ES, 2015 to 2020

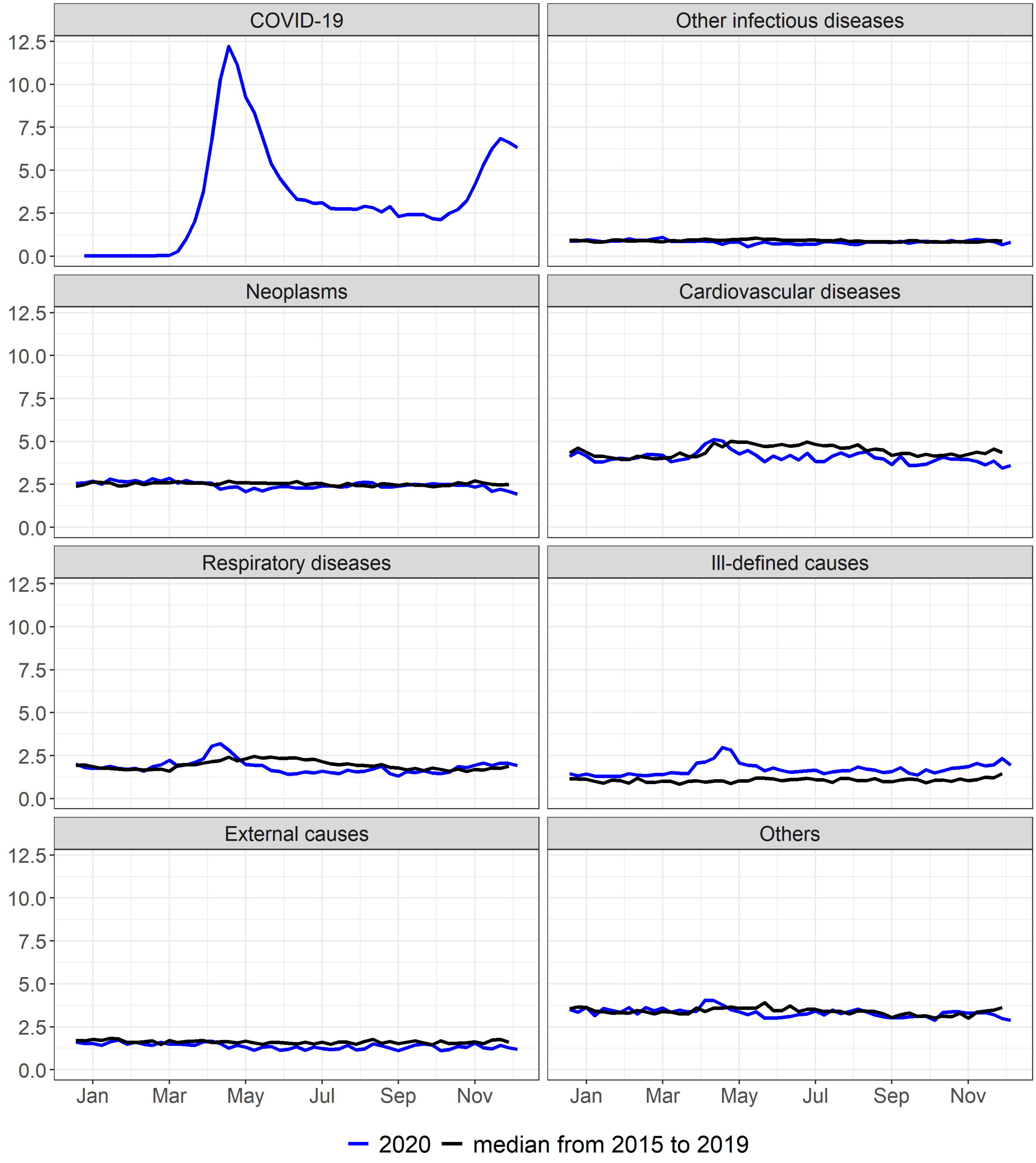


Supplementary figure 26. Mortality rate (per 100.000) by epidemiological week according to selected causes, RJ, 2015 to 2020

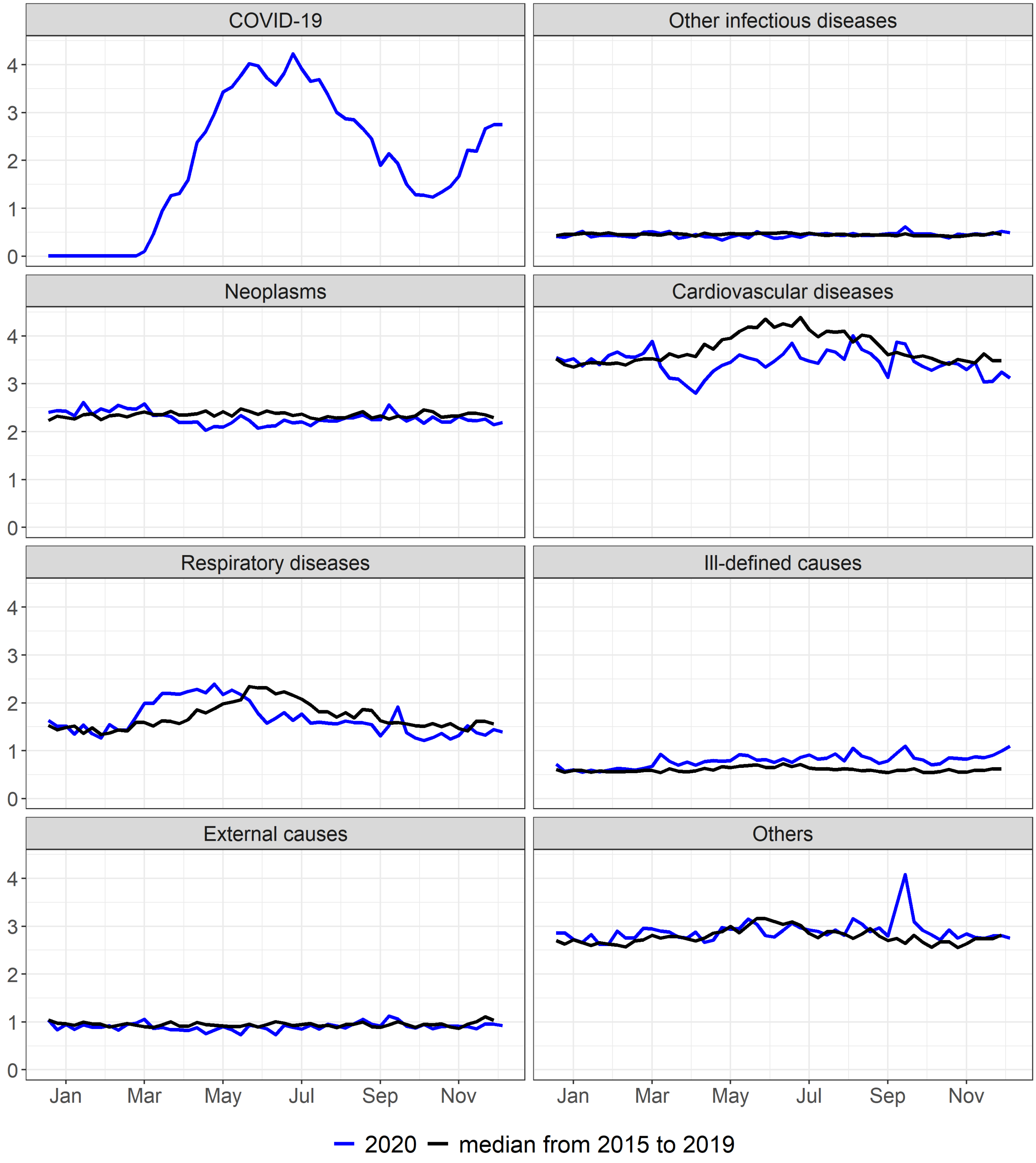


Supplementary figure 27. Mortality rate (per 100.000) by epidemiological week according to selected causes, SP, 2015 to 2020

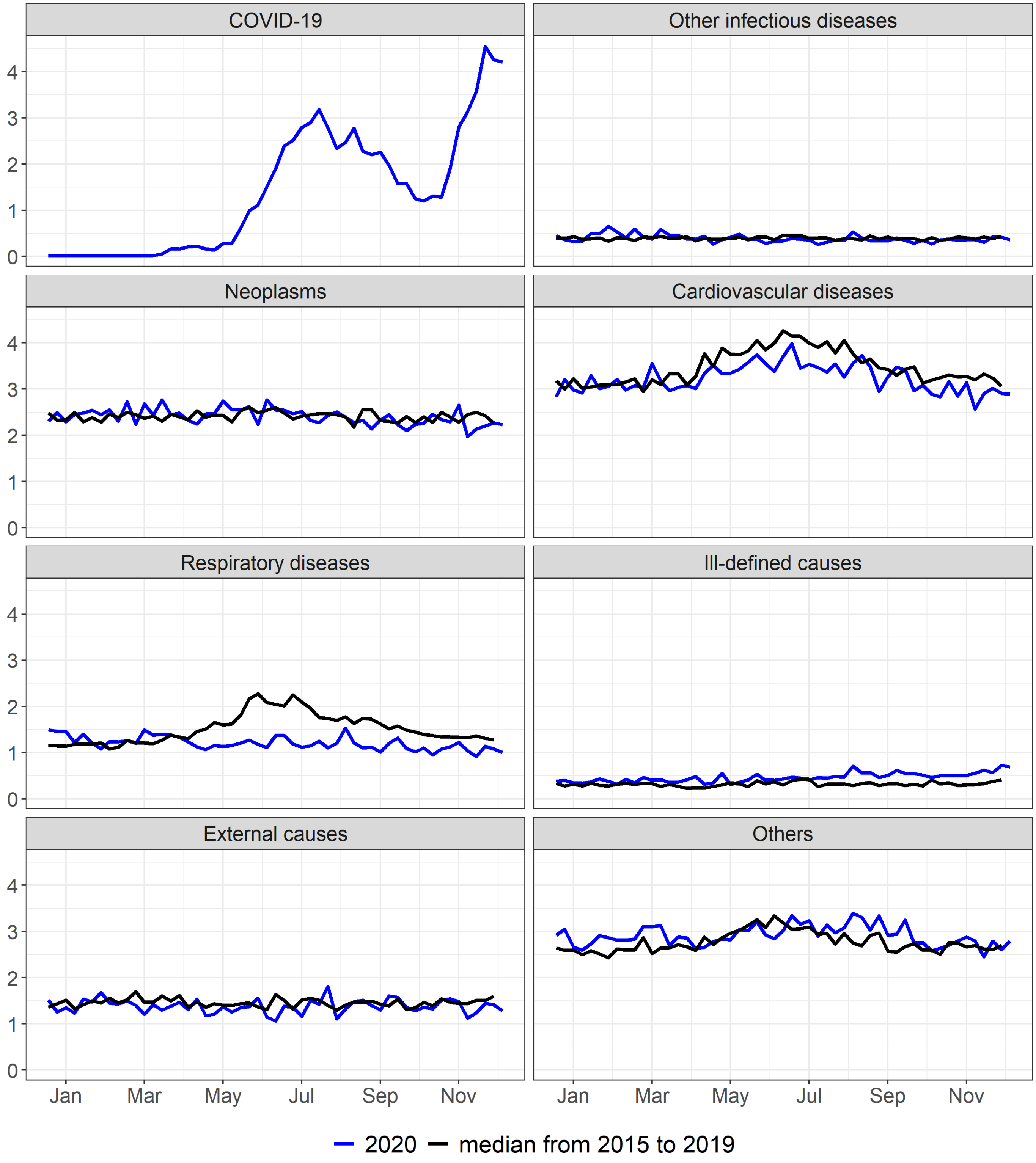


Supplementary figure 28. Mortality rate (per 100.000) by epidemiological week according to selected causes, PR, 2015 to 2020

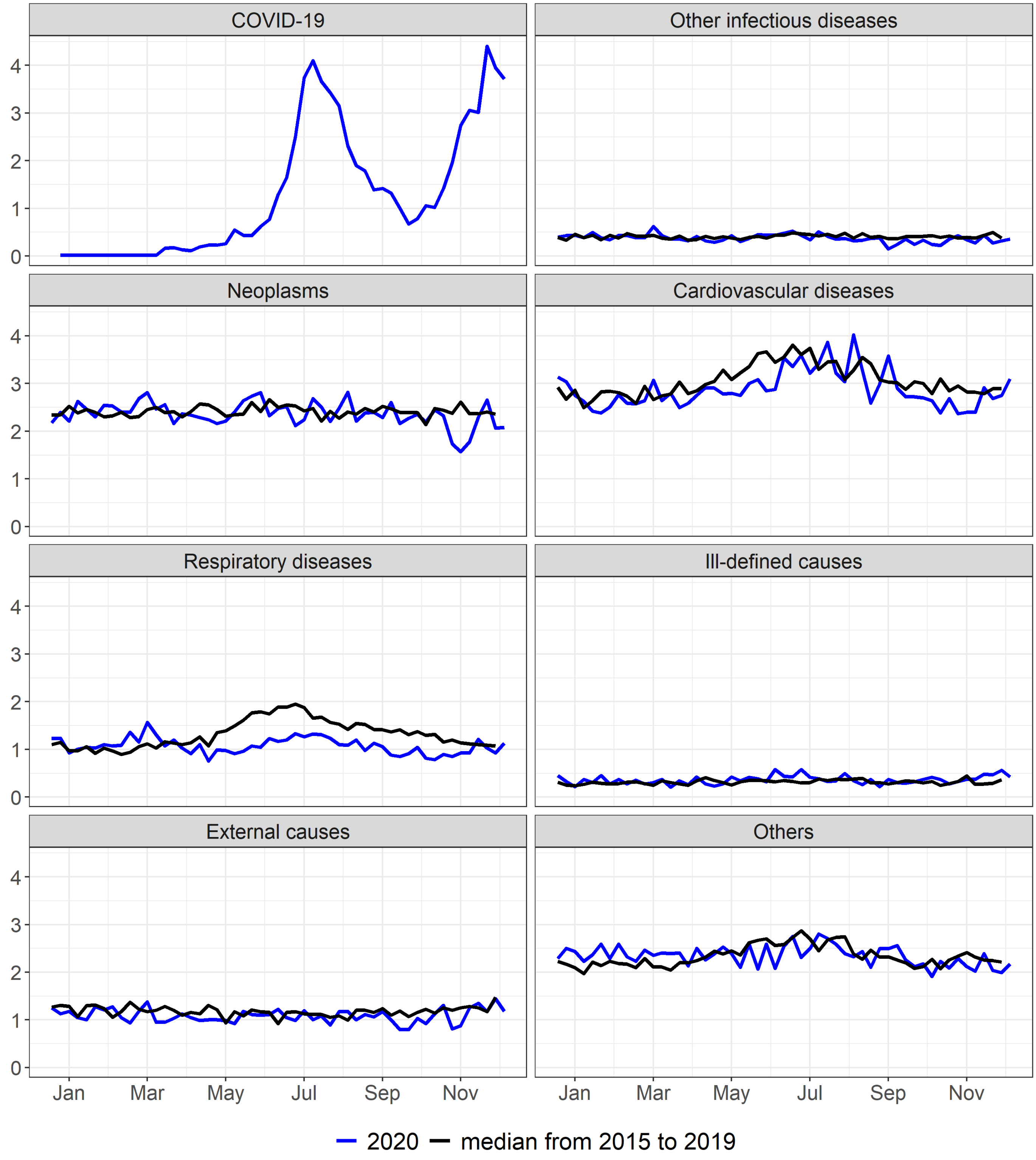


Supplementary figure 29. Mortality rate (per 100.000) by epidemiological week according to selected causes, SC, 2015 to 2020

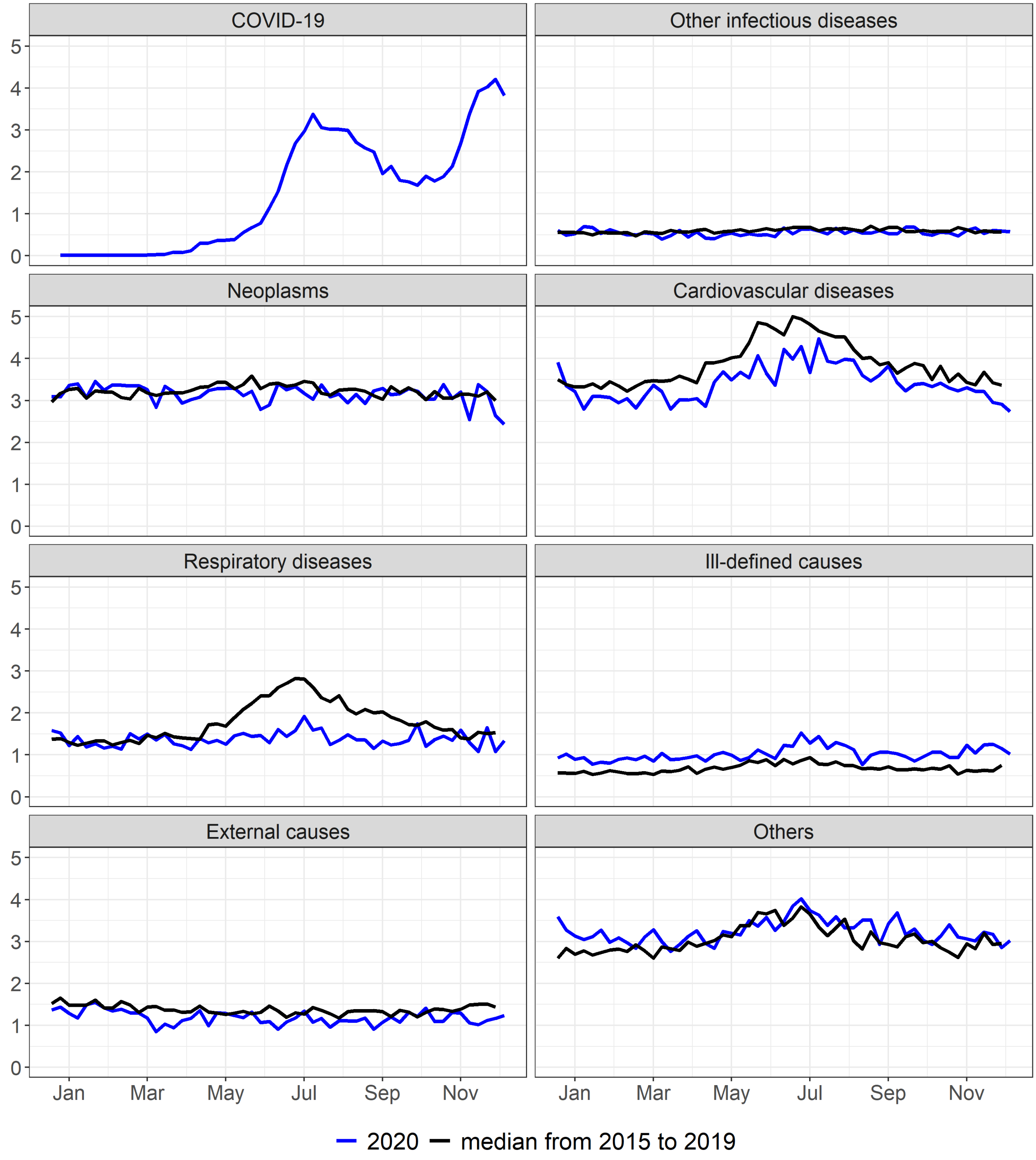


Supplementary figure 30. Mortality rate (per 100.000) by epidemiological week according to selected causes, RS, 2015 to 2020

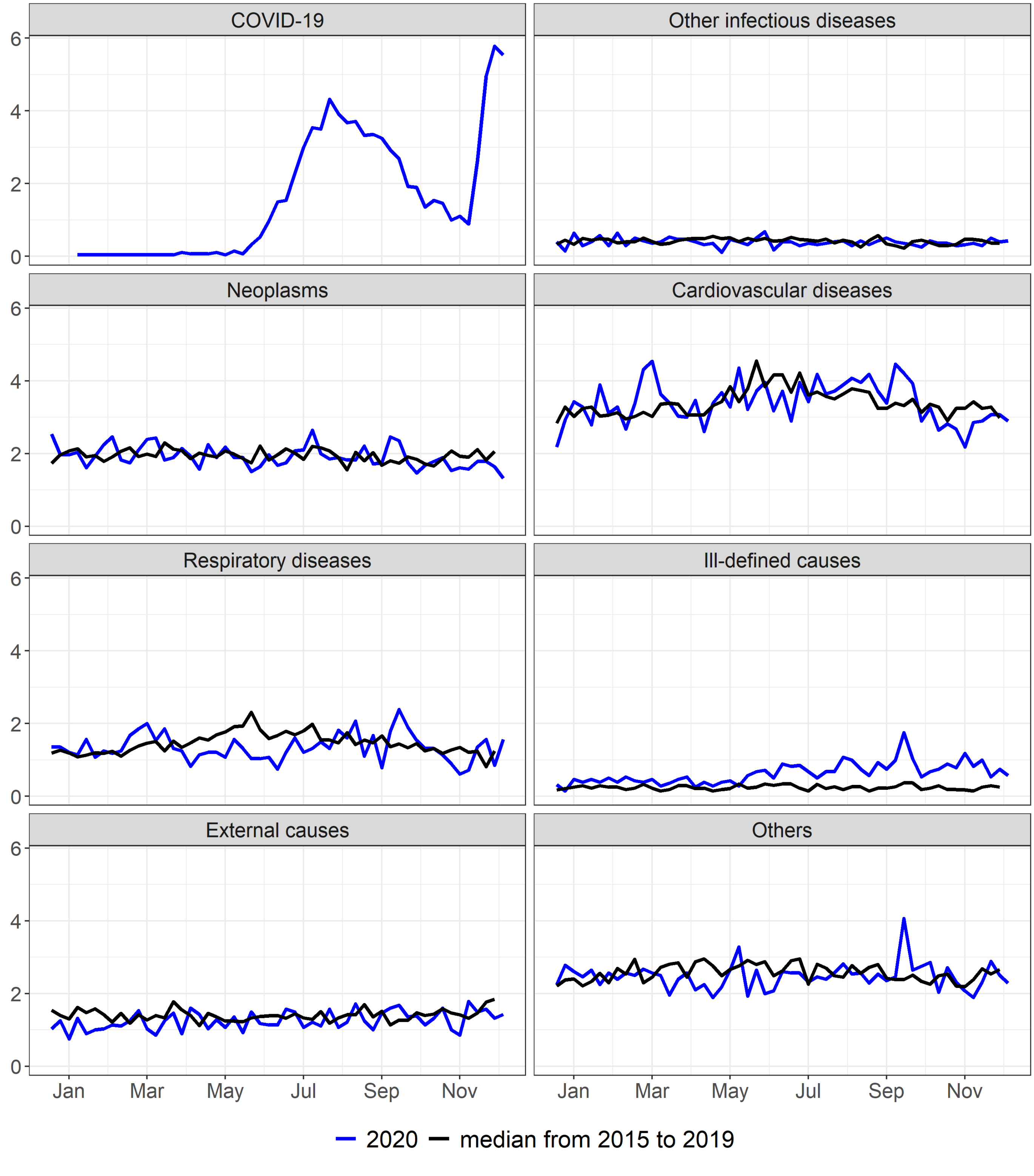


Supplementary figure 31. Mortality rate (per 100.000) by epidemiological week according to selected causes, MS, 2015 to 2020

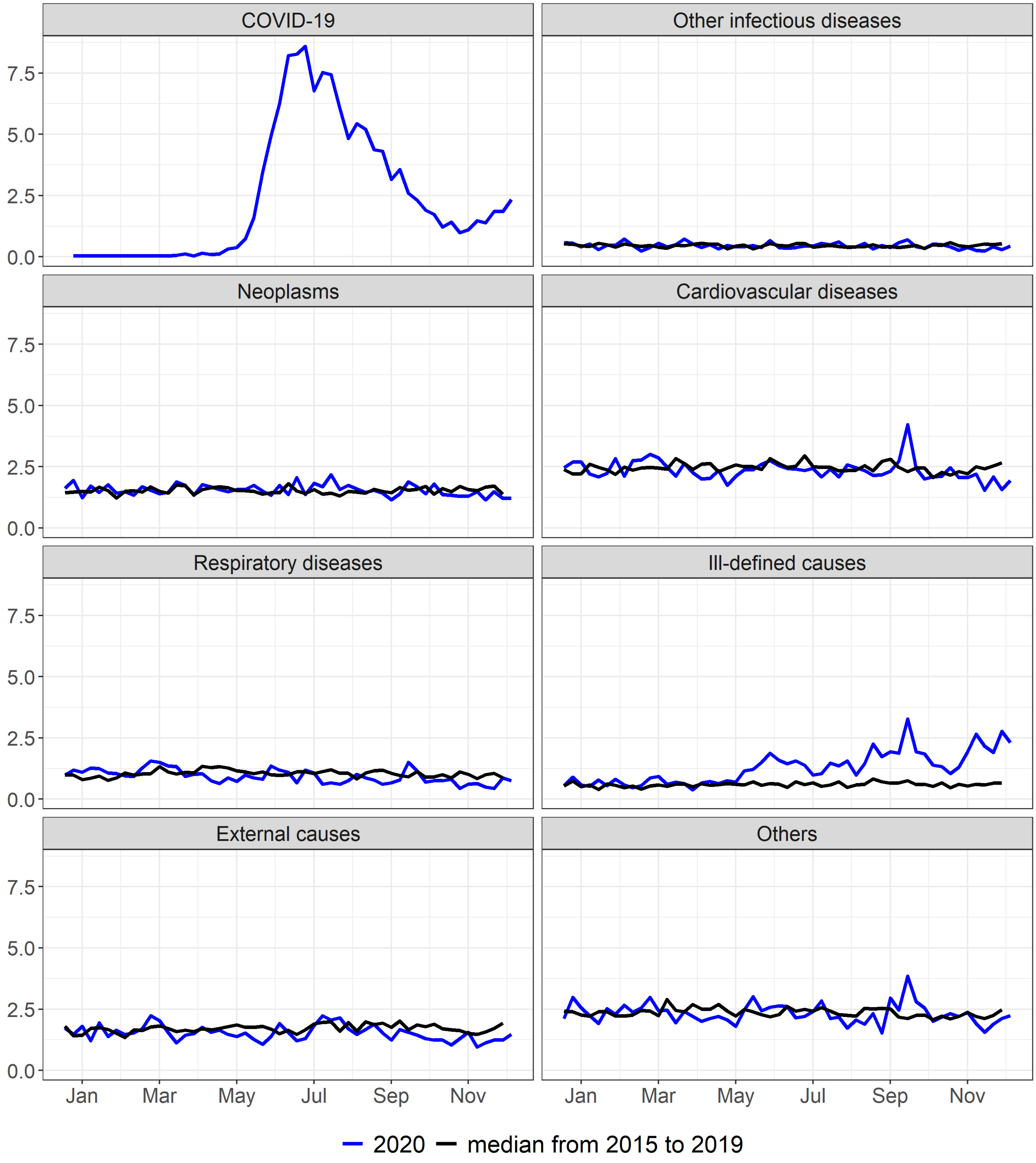


Supplementary figure 32. Mortality rate (per 100.000) by epidemiological week according to selected causes, MT, 2015 to 2020

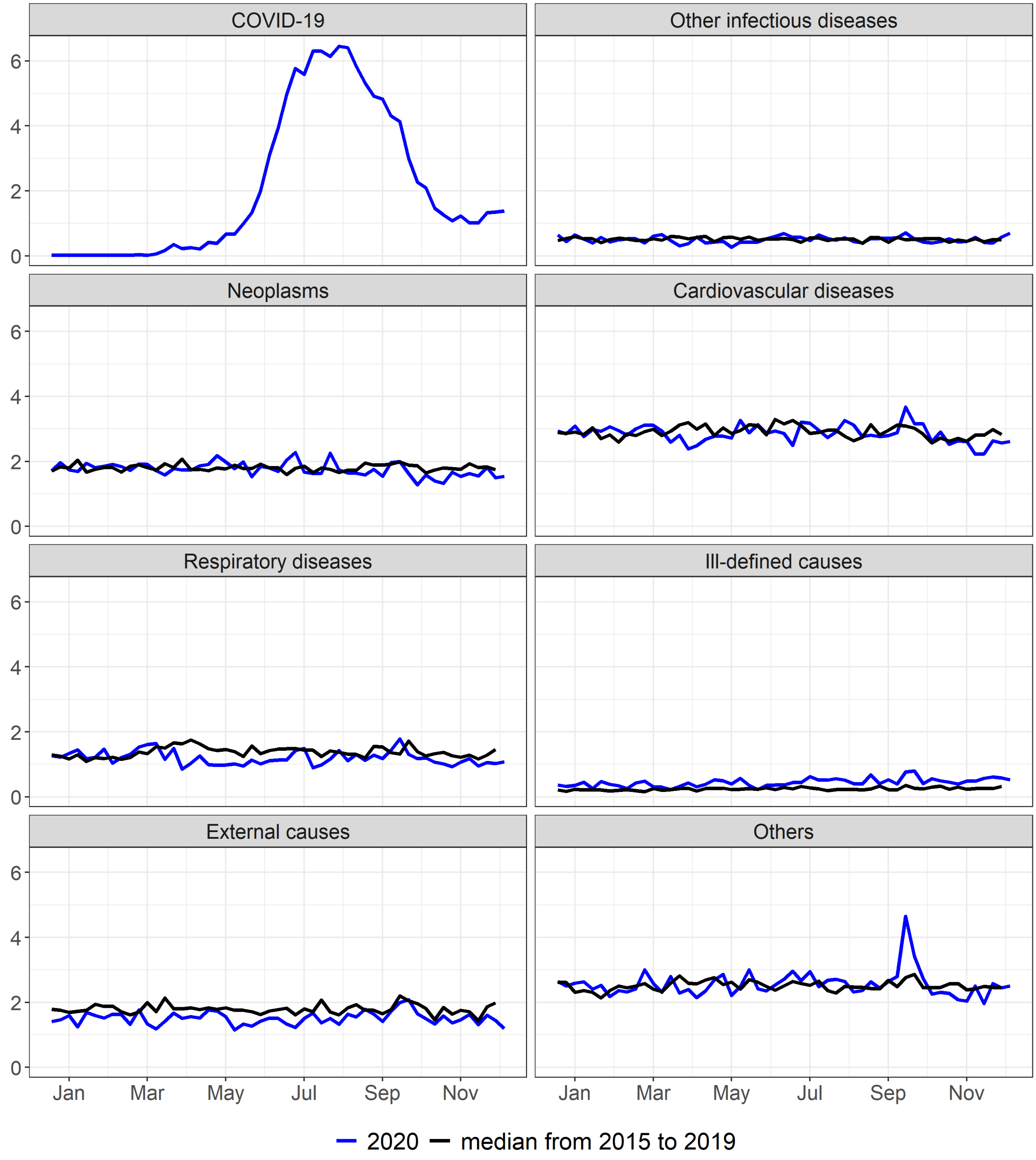


Supplementary figure 33. Mortality rate (per 100.000) by epidemiological week according to selected causes, GO, 2015 to 2020

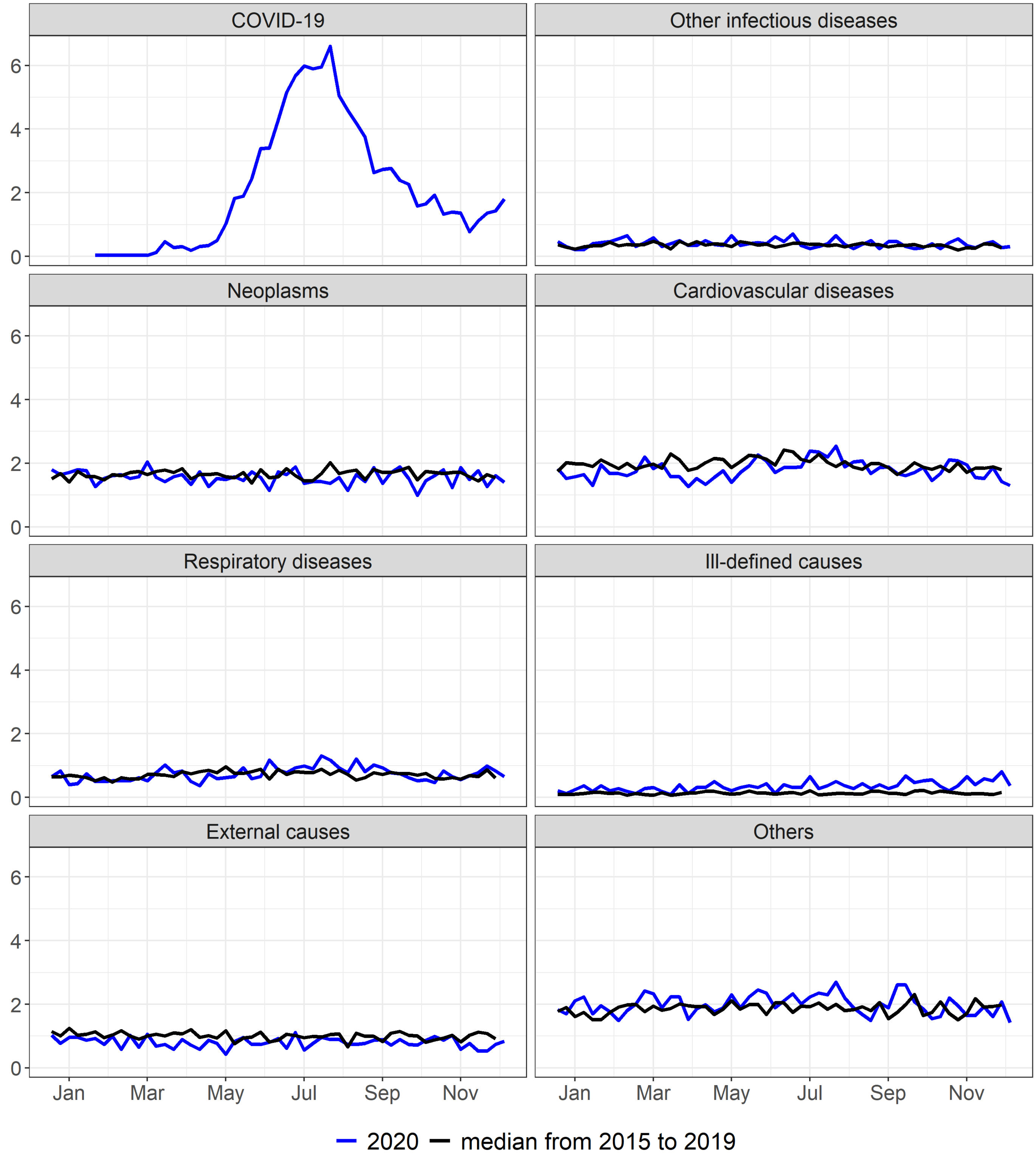


Supplementary figure 34. Mortality rate (per 100.000) by epidemiological week according to selected causes, DF, 2015 to 2020

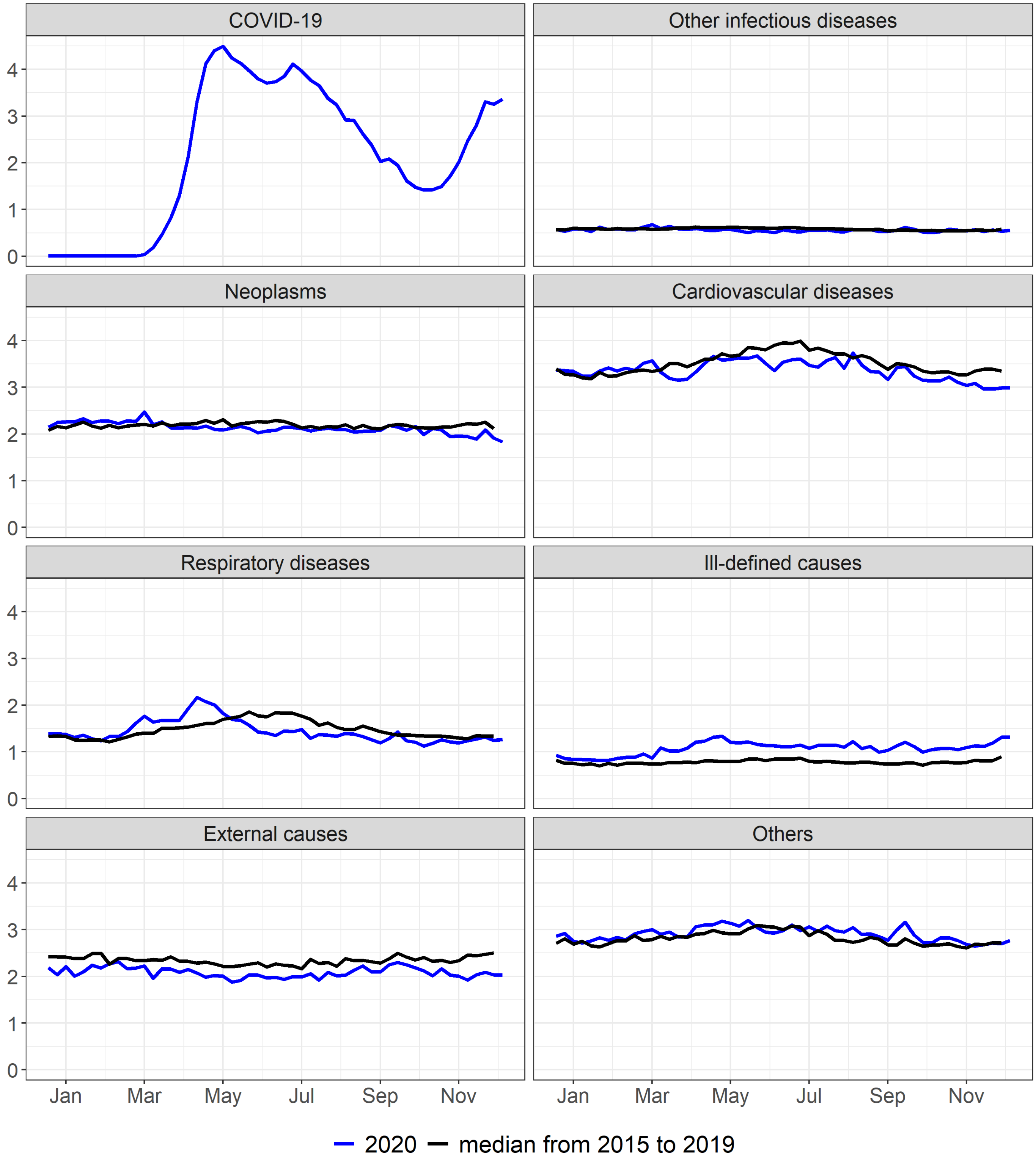


Supplementary figure 35. Mortality rate (per 100.000) by epidemiological week according to selected causes, Male sex, 2015 to 2020

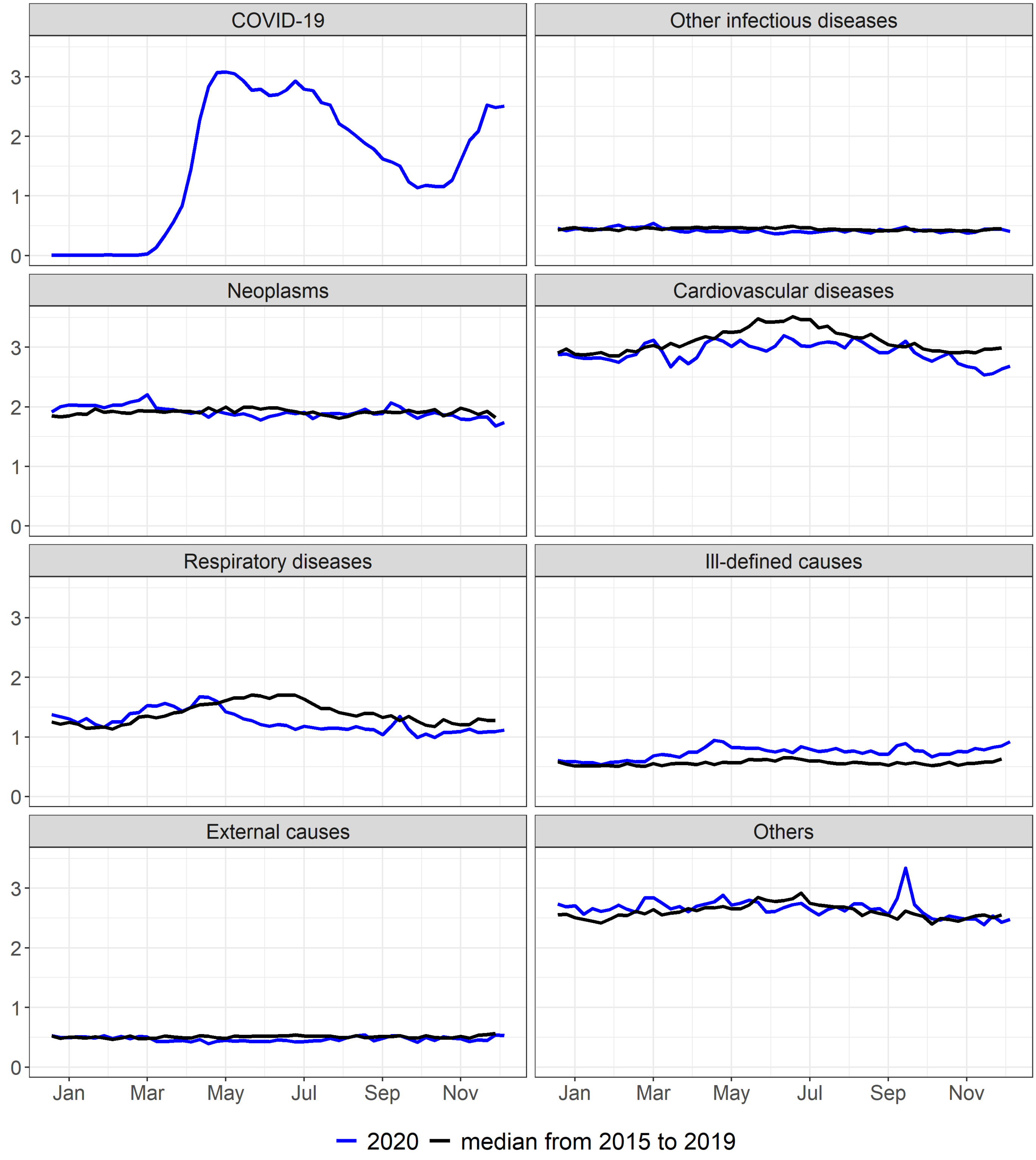


Supplementary figure 36. Mortality rate (per 100.000) by epidemiological week according to selected causes, Female sex, 2015 to 2020

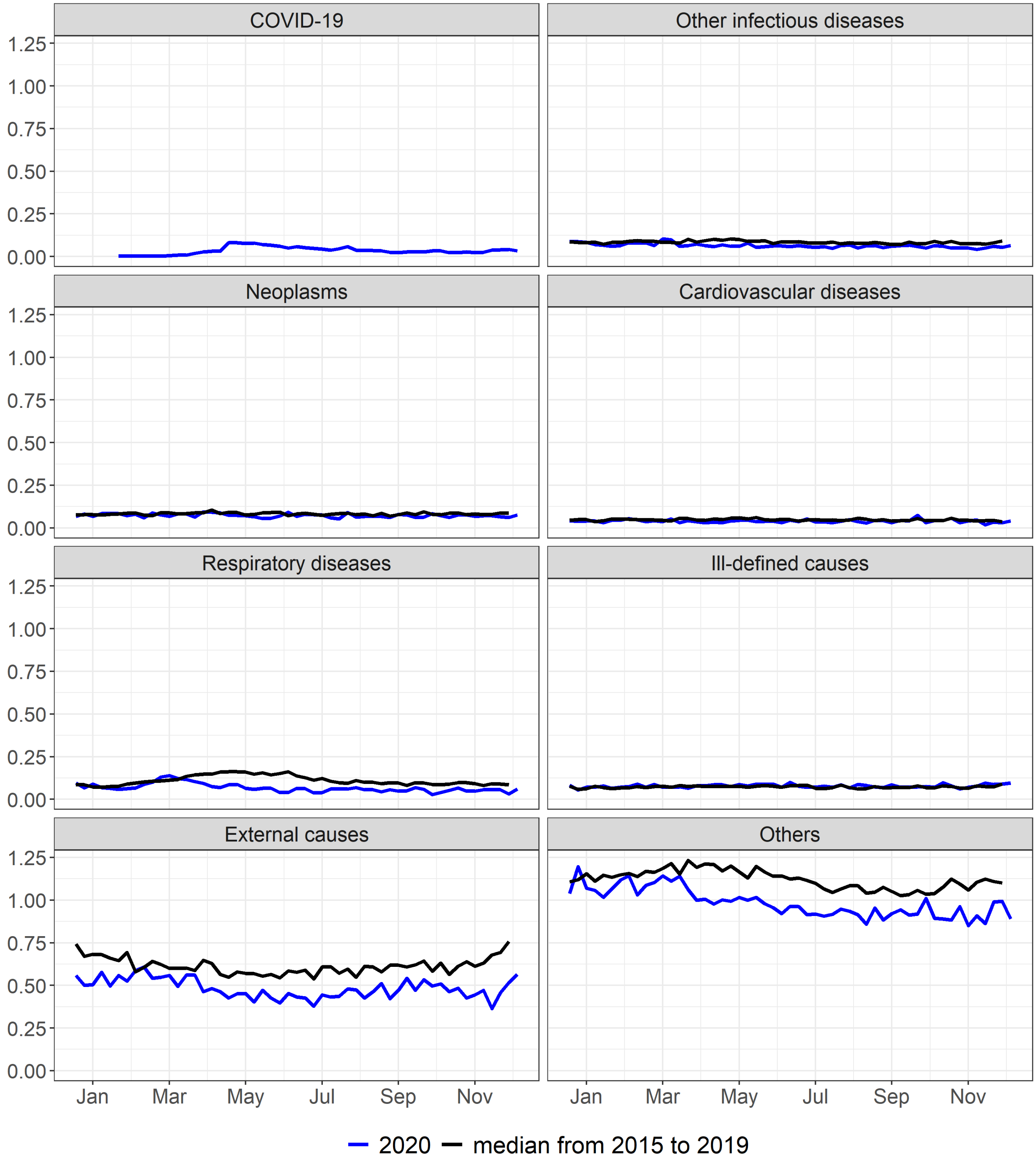


Supplementary figure 37. Mortality rate (per 100.000) by epidemiological week according to selected causes, Age_00_to_19, 2015 to 2020

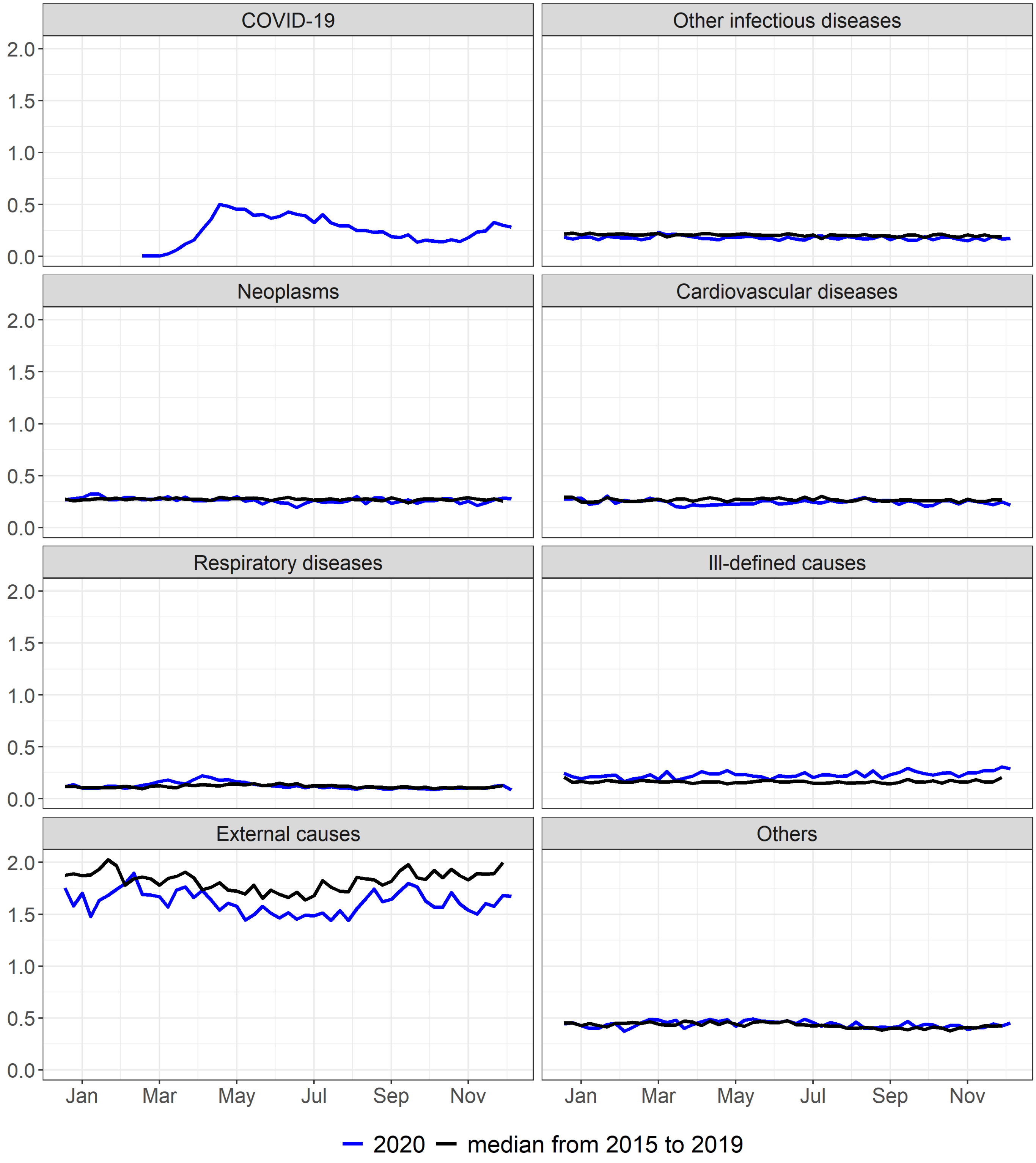


Supplementary figure 38. Mortality rate (per 100.000) by epidemiological week according to selected causes, Age_20_to_39, 2015 to 2020

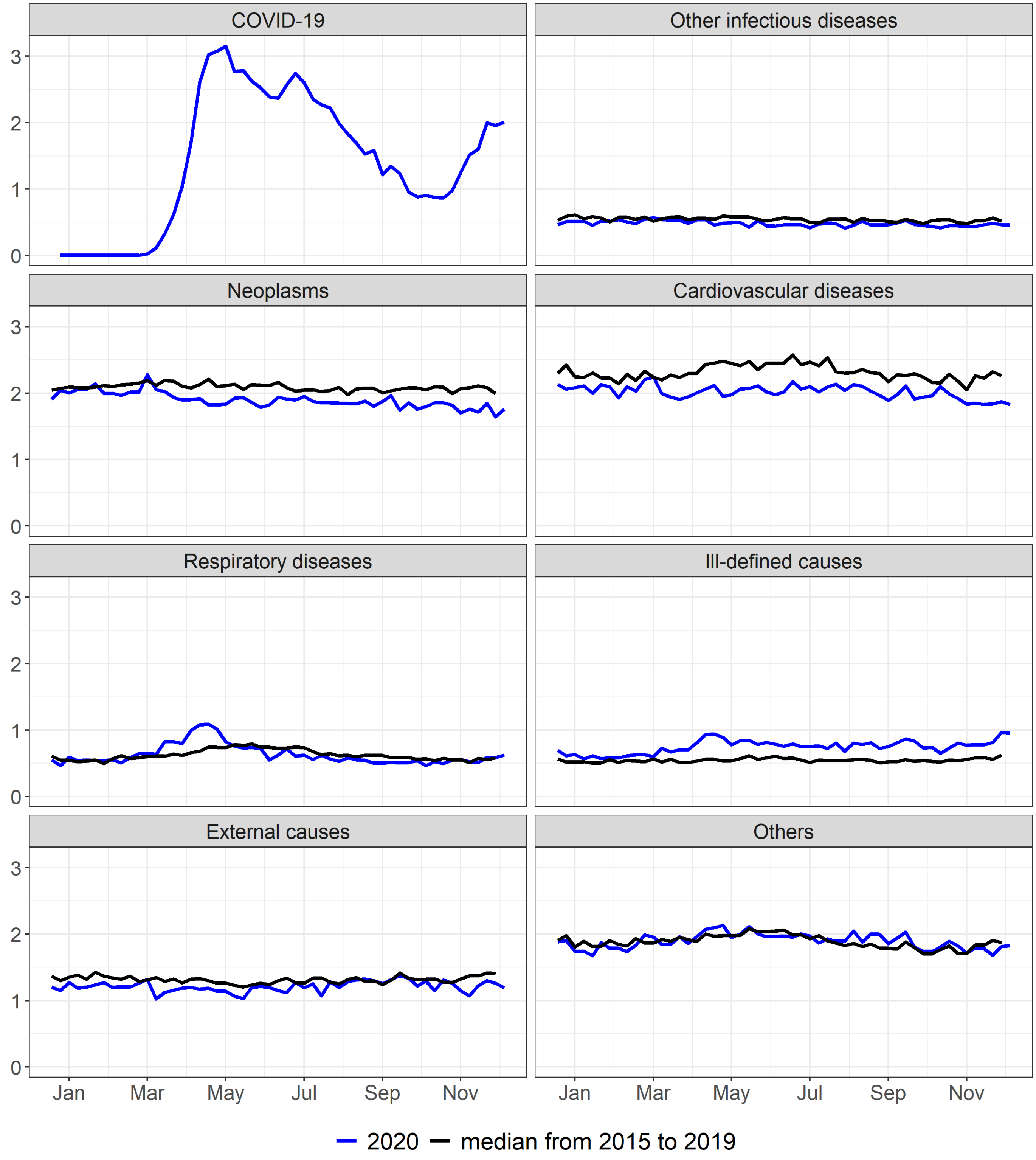


Supplementary figure 39. Mortality rate (per 100.000) by epidemiological week according to selected causes, Age_40_to_59, 2015 to 2020

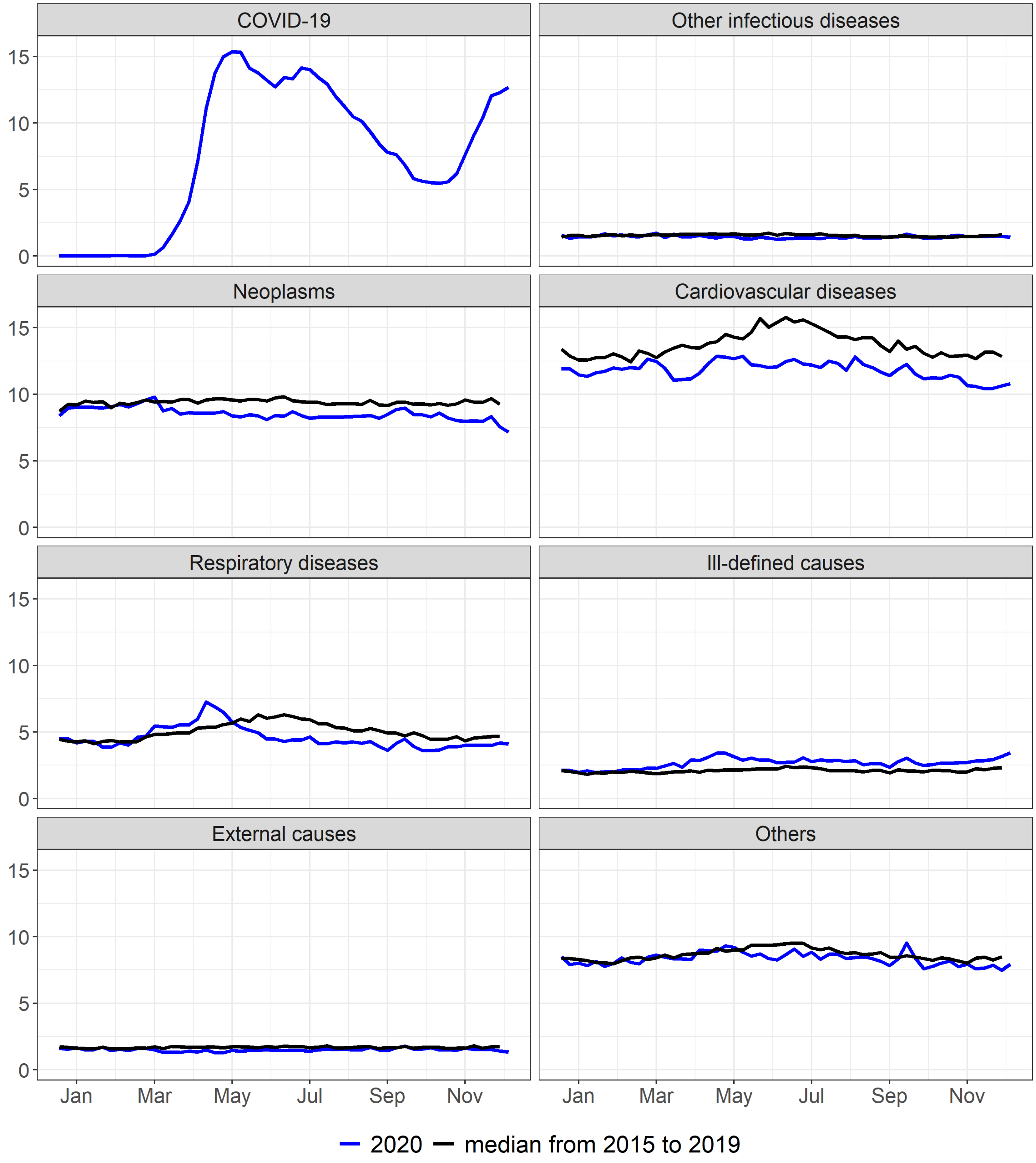


Supplementary figure 40. Mortality rate (per 100.000) by epidemiological week according to selected causes, Age_60_to_79, 2015 to 2020

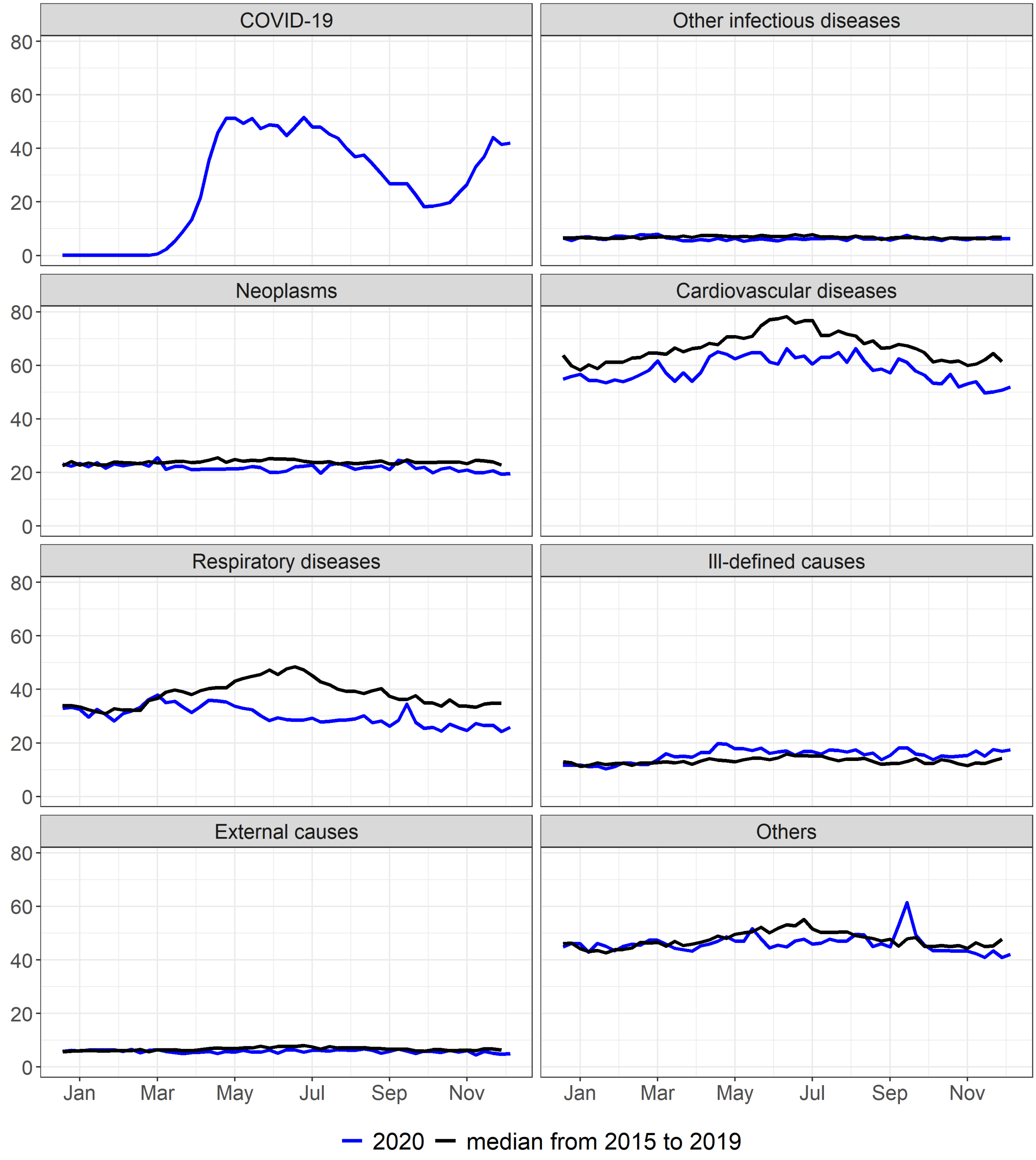


Supplementary figure 41. Mortality rate (per 100.000) by epidemiological week according to selected causes, Age_80_or_older, 2015 to 2020

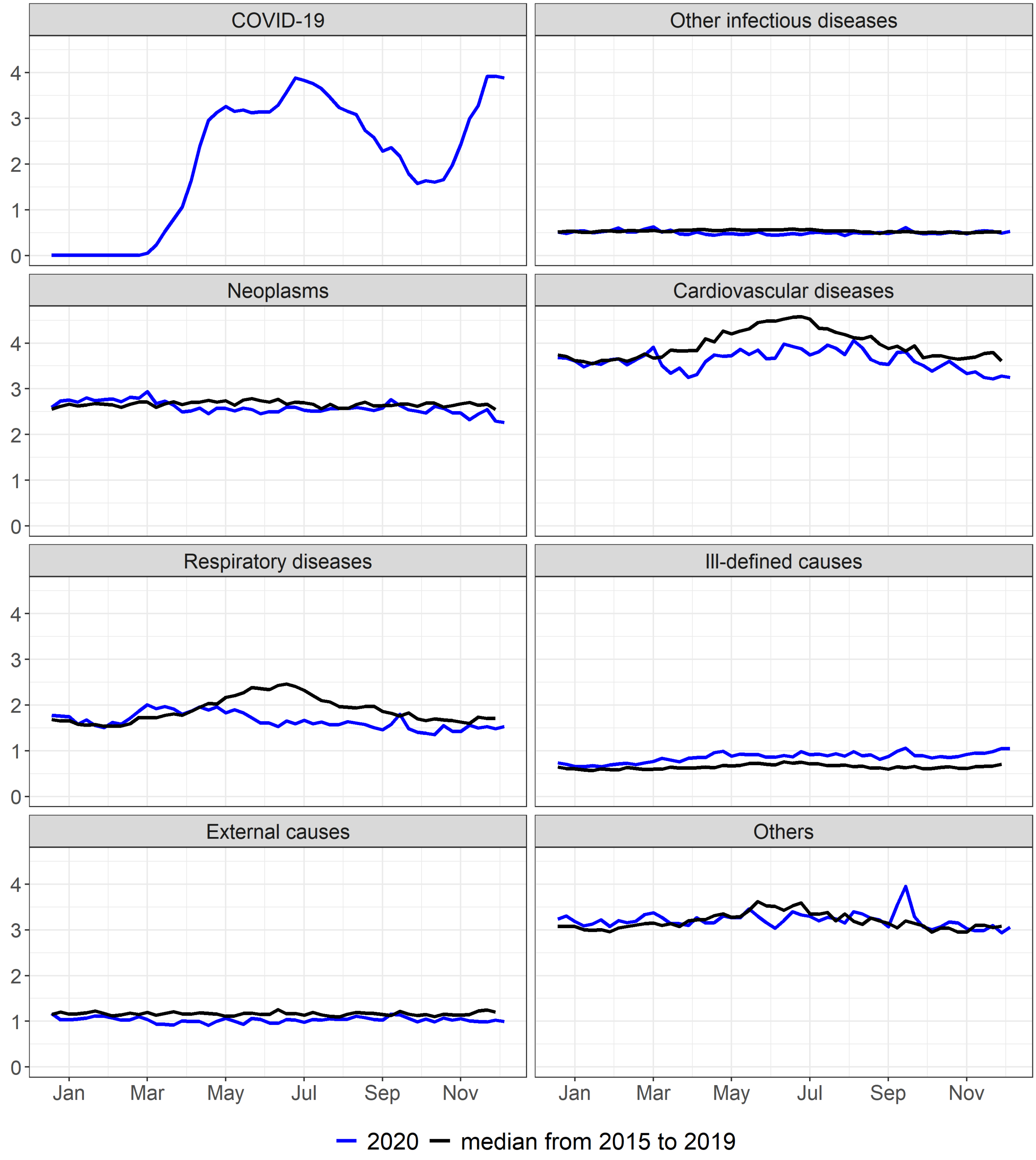


Supplementary figure 42. Mortality rate (per 100.000) by epidemiological week according to selected causes, White race, 2015 to 2020

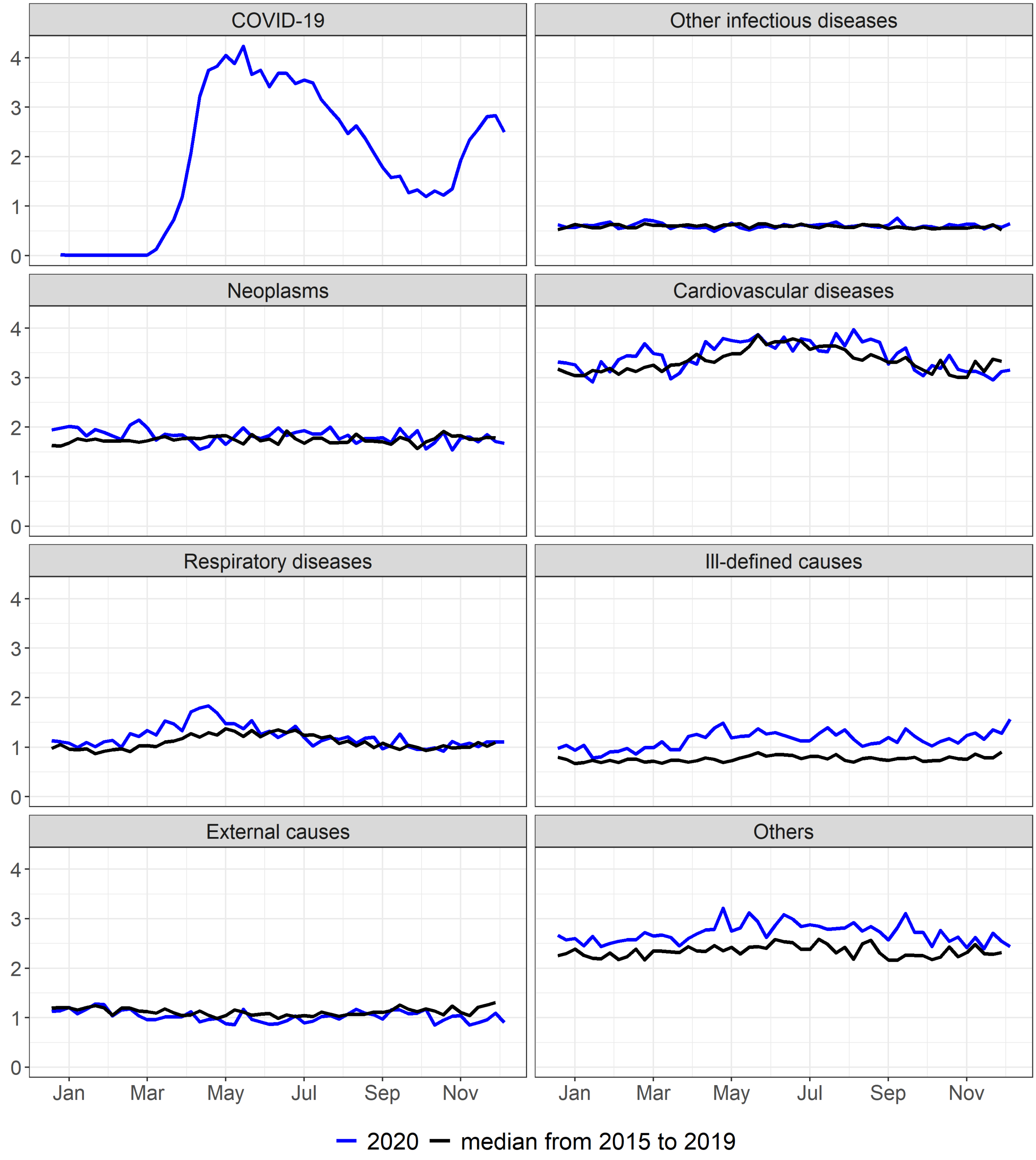


Supplementary figure 43. Mortality rate (per 100.000) by epidemiological week according to selected causes, Black race, 2015 to 2020

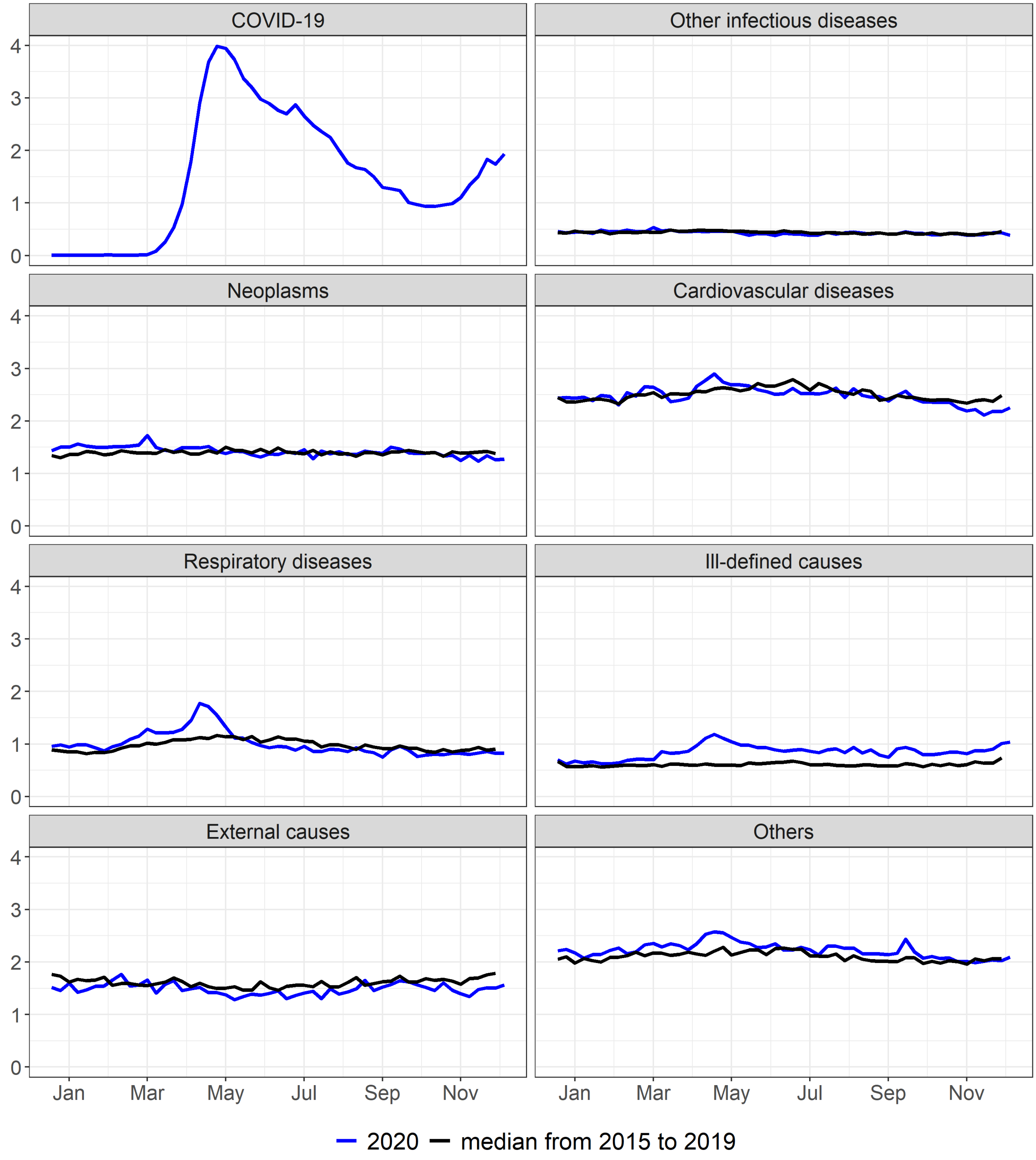


Supplementary figure 44. Mortality rate (per 100.000) by epidemiological week according to selected causes, Brown race, 2015 to 2020

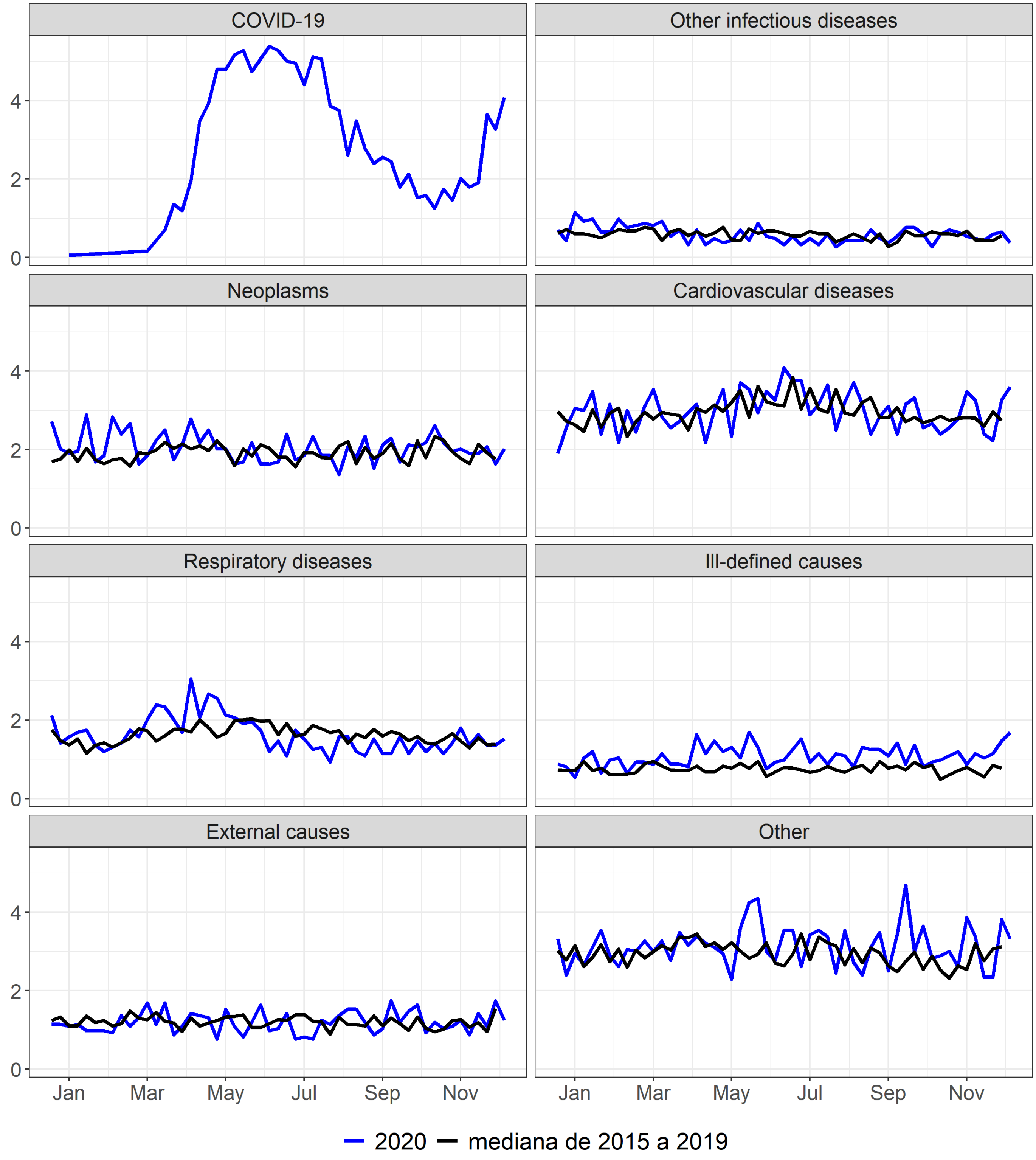


Supplementary figure 45. Mortality rate (per 100.000) by epidemiological week according to selected causes, Other race, 2015 to 2020

#### Supplementary figure 46. Alarms identified by the Farrington algorithm, Northeast region, Brazil, 2015 to 2020

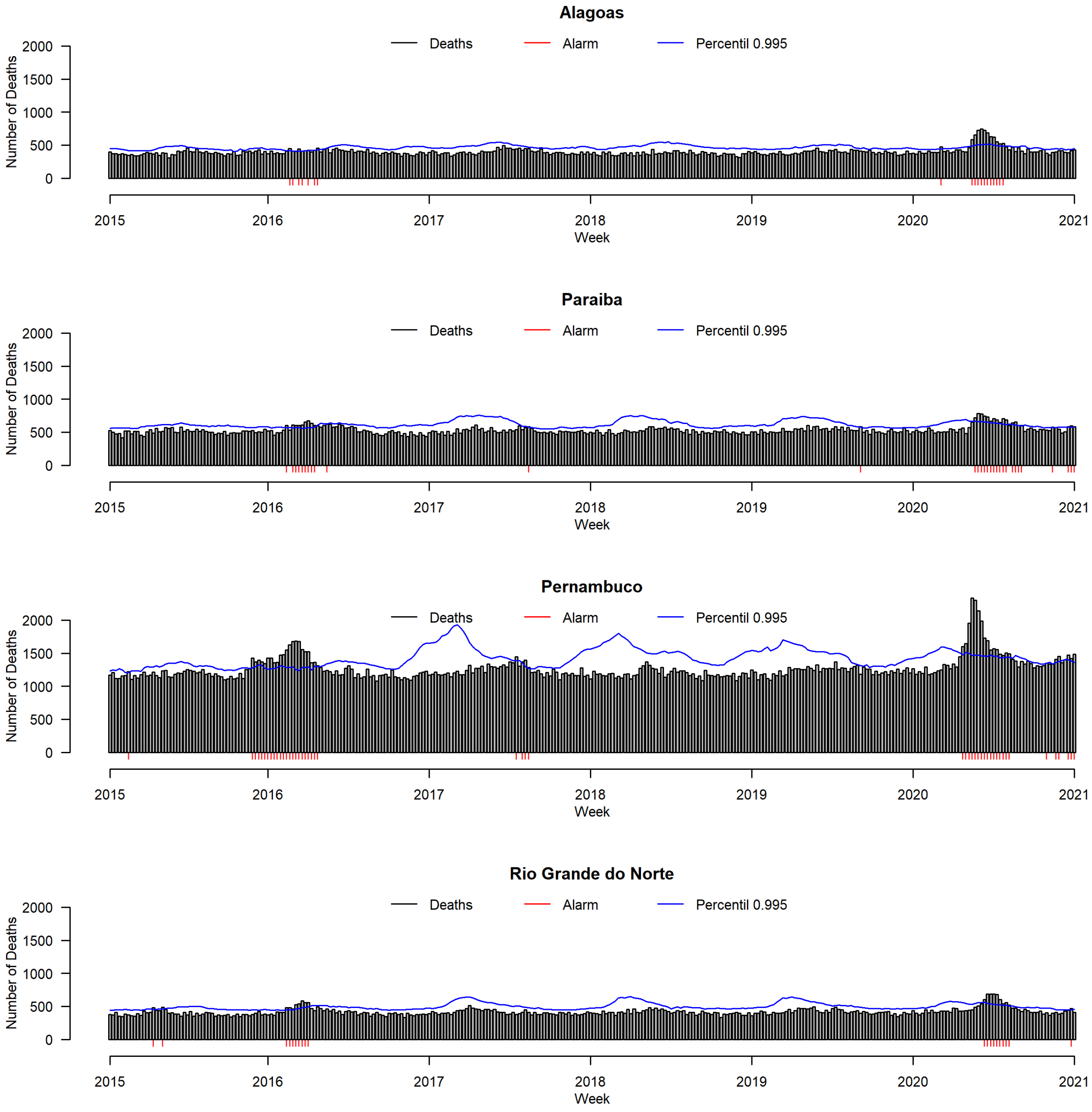

**Supplementary figure 47. Alarms identified by the Farrington algorithm, Southeast region, Brazil, 2015 to 2020**

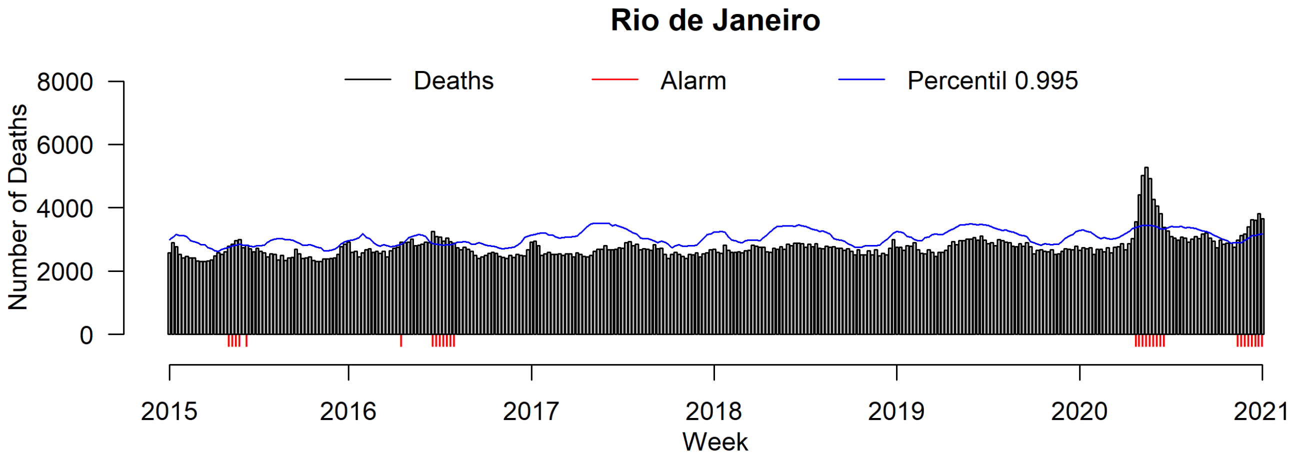


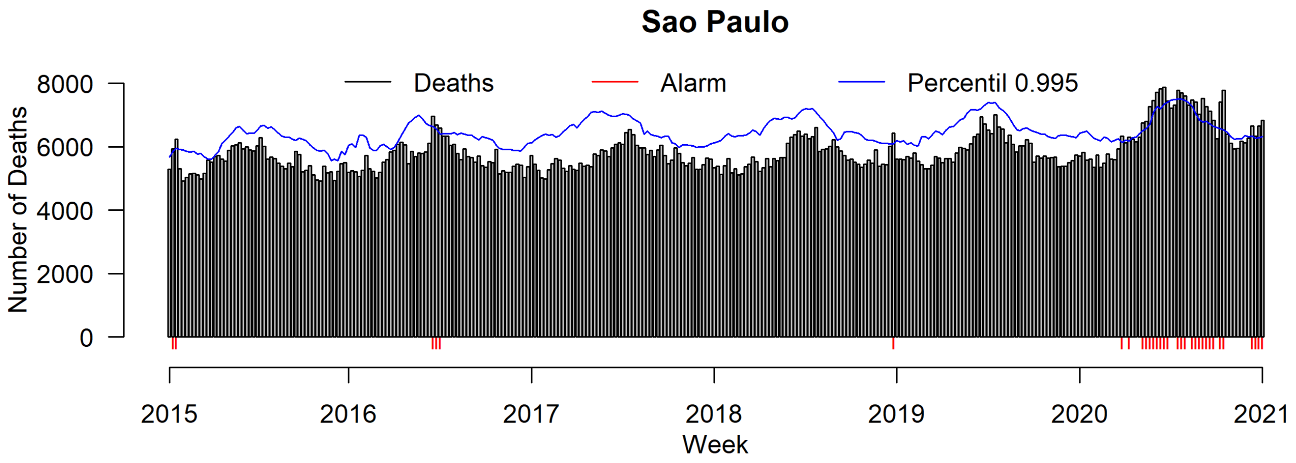

**Supplementary figure 48. Alarms identified by the Farrington algorithm, South region, Brazil, 2015 to 2020**

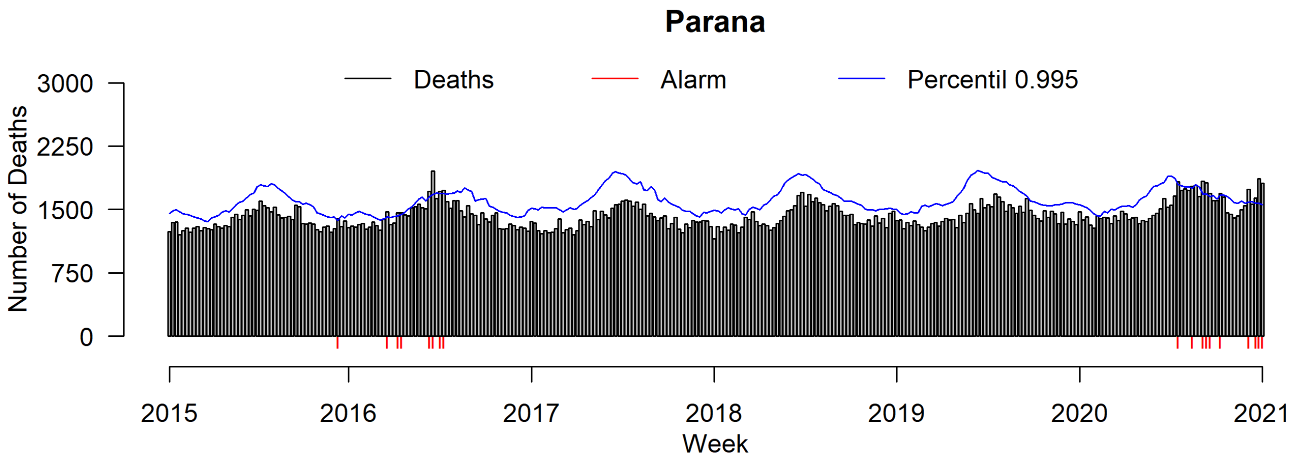


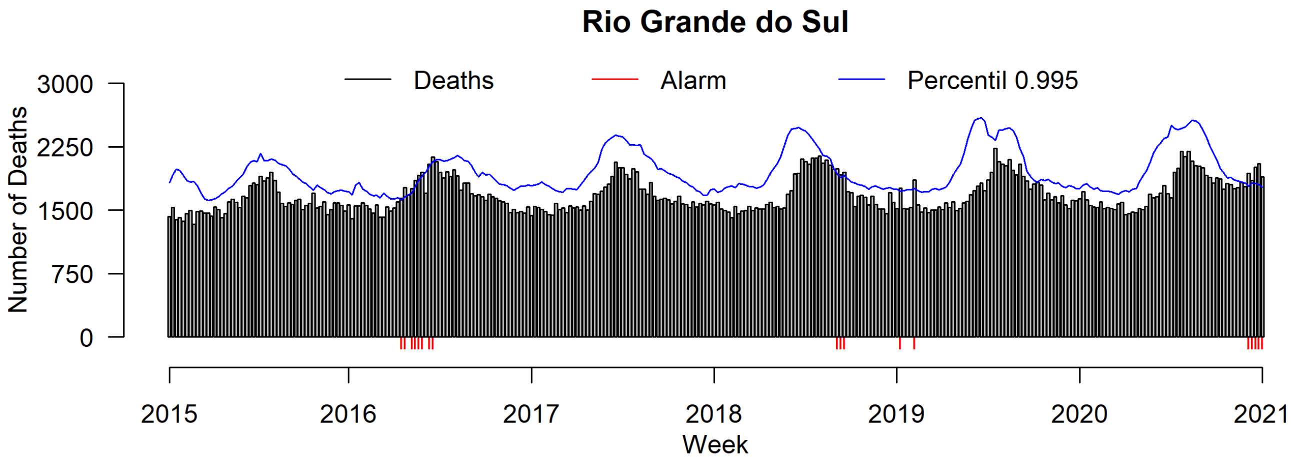
